# Common genetic associations between age-related diseases

**DOI:** 10.1101/2020.06.16.20132621

**Authors:** Handan Melike Dönertaş, Daniel K. Fabian, Matías Fuentealba Valenzuela, Linda Partridge, Janet M. Thornton

## Abstract

Age is a common risk factor in many diseases, but the molecular basis for this relationship is elusive. In this study we identified 4 disease clusters from 116 diseases in the UK Biobank data, defined by their age-of-onset profiles, and found that diseases with the same onset profile are genetically more similar, suggesting a common etiology. This similarity was not explained by disease categories, co-occurrences or disease cause-effect relationships. Two of the four disease clusters had an increased risk of occurrence from age 20 and 40 years respectively. They both showed an association with known aging-related genes, yet differed in functional enrichment and evolutionary profiles. Moreover, they both had age-related expression and methylation changes. We also tested mutation accumulation and antagonistic pleiotropy theories of aging and found support for both.

## Introduction

Aging is associated with a time-dependent decrease in the functional integrity of organisms and an increase in susceptibility to pathologies^1^. The worldwide increase in lifespan has not been matched by an increase in healthspan, and there is a growing period of loss of function, and disease at the end of life^2^. Aging thus poses a significant global challenge, because it is the major risk factor for chronic conditions, including cardiovascular disease, cancer, and dementia^3^. Although these diseases involve different organs and pathologies, they all show a strong dependence on age^4^ and could, therefore share common etiologies based upon the underlying mechanisms of aging. It is therefore important to understand if the aging process itself leads to different age-related conditions through common pathways, or if the age-dependency of different diseases has independent, time-dependent causes.

Despite the negative impact of aging on organismal fitness and functionality, it is widespread in the animal world as well as in humans^5^ and has therefore been described as an evolutionary paradox^6^. Aging can nonetheless evolve as the force of natural selection weakens with age due to extrinsic hazard. Mutations that are deleterious only in later life can accumulate in the population through mutation pressure, because the force of natural selection eliminating them from the population declines with the age of onset of their effects (mutation accumulation theory of aging)^7^. Pleiotropic variants that are beneficial during early life but detrimental later in life can also become prevalent in populations through natural selection (antagonistic pleiotropy theory of aging)^8^. Thus, genome-wide germline genetic variants that increase the risk of diseases at old age may not be pruned by natural selection or may be associated with beneficial phenotypes earlier in life.

The risk of many **<u>a</u>**ge-**<u>r</u>**elated **<u>d</u>**iseases (ARDs) is influenced by genetic variation. **<u>G</u>**enome-**<u>w</u>**ide **<u>a</u>**ssociation **<u>s</u>**tudies (GWAS) have identified genetic variants that alter complex traits. Pleiotropy, where variants or genes influence multiple traits, is more prevalent than previously thought^9–12^, indicating that different traits share common causal pathways^13^. Pleiotropy within the disease classification system^12^ and in certain disease classes, such as immune-related diseases^14,15^ and cancer^16^, has been studied, but the understanding of pleiotropy in ARDs more broadly is limited. Some studies have investigated the common pathways between manually curated age-related traits^17–19^. Despite the challenges of combining results from different published datasets, these studies provided the first clues that at least some ARDs share common pathways, which are also related to a significant but limited number of longevity-regulating genes in model organisms. In this study, using disease age-of-onset profiles, we extend the previous efforts by providing the first data-driven classification of a large number of diseases according to their age-profile, followed by a genetic analysis in one of the largest and most comprehensive cohorts available. In this way, we provide a comparative genetic analysis between ARDs and non-ARDs and also between ARDs with different age profiles.

The **<u>UK B</u>**io**<u>b</u>**ank (UKBB)^20^ includes genetic and health-related data for almost half a million participants. We extracted age-of-onset profiles for 116 diseases and identified unbiased clusters to define the relationship between disease incidence and age. We identified variants associated with each disease and compared the genetic associations between diseases based on these clusters. We first found that diseases with the same age profile share genetic associations, which cannot be explained by disease categories, co-occurrences, or mediated pleiotropy, and thus reflects a common etiology (Figure S1). We further characterized these shared associations compared to previously known longevity-associated genes, genes with age-dependent expression or methylation changes, and biological functions. Finally, we compared the variants associated with diseases that start to occur at different ages and identified different evolutionary characteristics, supporting the mutation accumulation and antagonistic pleiotropy theories of aging.

## Results

### Data

We used self-reported diseases and age-at-diagnosis covering 484,598 participants, and their genotypes in the UKBB^20^. Age-at-diagnosis for self-reported diseases ranges from the age of 0 to 70, and is less biased than age-at-diagnosis for ICD10 codes, which includes data for only after 1992^21^. Details of the UKBB data, quality control steps, and exploratory analyses are given in the Supplementary Information and Supplementary Figures S2-9. Self-reported diseases in the UKBB are hierarchically structured and the top nodes; such as cardiovascular or endocrine diseases, were considered as *disease categories* (Figure S7). We only analyzed common diseases (*i*.*e*. with at least 2,000 cases) that were not sex-limited (n=116 in 472 self-reported diseases). Importantly, we did not include cancer in our analyses as the interaction between genetic and environmental contributors is likely different from non-cancer illnesses, even though they may have a similar age-of-onset profile (for details, see Methods).

### Age-of-onset clusters

Age is associated with increased risk of many diseases. In order to characterize the association between age and different diseases we first used age-at-diagnosis as a proxy to disease onset and derived disease age-of-onset profiles (Figure S10-19). On average, cardiovascular and endocrine diseases had a high median age-of-onset, while infections had the lowest age-of-onset (Figure S20). We then clustered diseases into 4 clusters (the optimum number determined by the gap statistic) using the PAM algorithm and disease dissimilarities calculated using CORT distance^22^ (Figure 1, Table S1). Cluster 1 diseases (n=25) showed a rapid increase with age after the age of 40; 11 were cardiovascular diseases, but the cluster also included other diseases such as diabetes, osteoporosis, and cataract. Cluster 2 (n=51) diseases started to increase in the population at an earlier age of 20, but had a slower rate of increase with age; the diseases in this cluster were the most diverse, including 17 musculoskeletal, 13 gastrointestinal diseases, as well as others such as anemia, deep venous thrombosis, thyroid problems, depression. Cluster 3 diseases (n=30) showed a low age dependency with a mostly uniform distribution across ages, but with slight increases around the ages of 10 and 60 years. This category included similar numbers of immunological, neurological, musculoskeletal, gastrointestinal and respiratory diseases but all have an ‘immune’ component even if not classified in this way by the UKBB (e.g., inflammatory bowel disease (gastrointestinal), asthma (respiratory), psoriasis (dermatology)). Cluster 4 (n=10) had a peak at around 0-10 years of age and included respiratory diseases (n=5) and infections (n=4). Notably, all infectious diseases were in this cluster.

**Figure 1:**
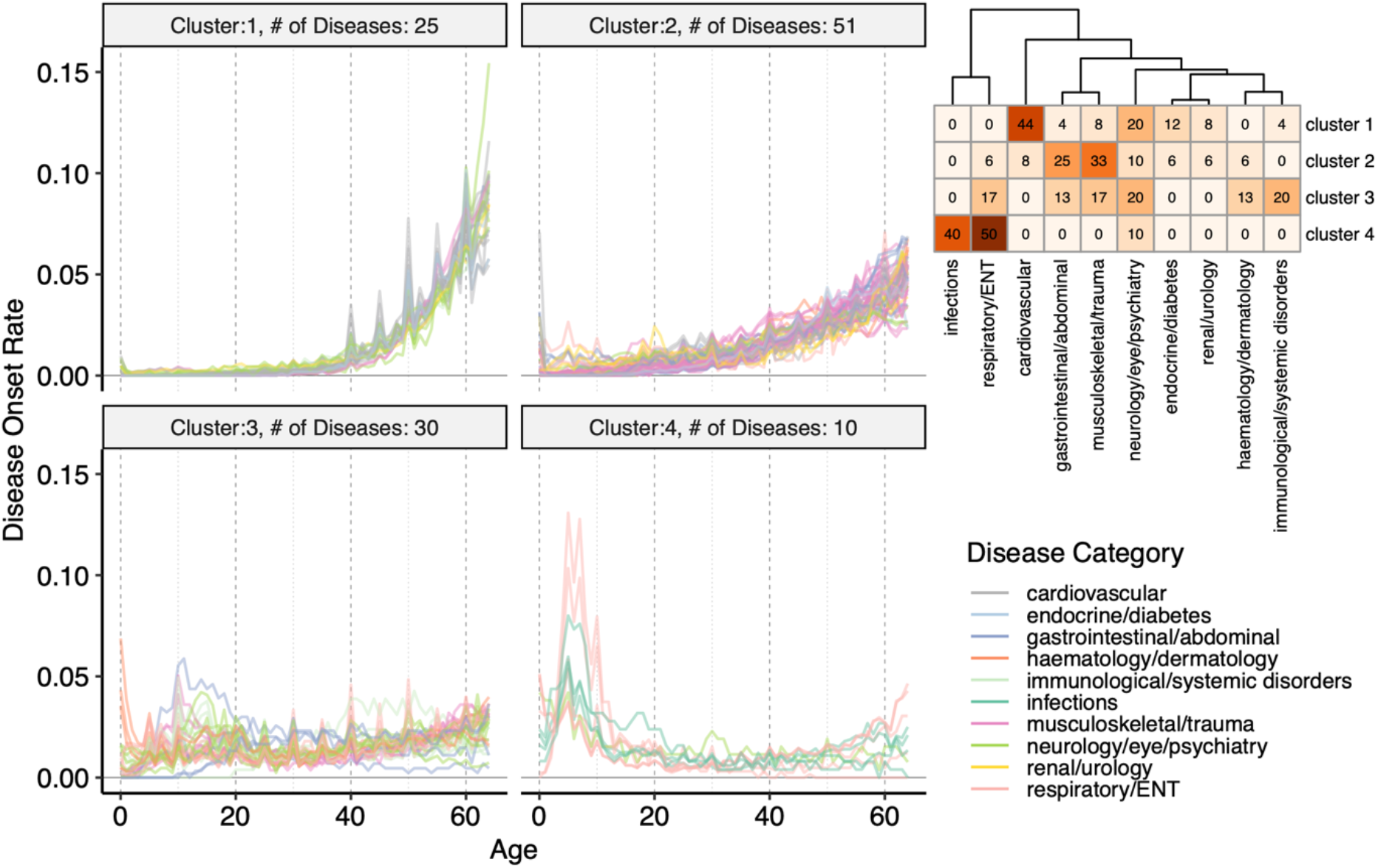
Age-of-onset profiles clustered by the PAM algorithm, using dissimilarities calculated with **<u>t</u>**emporal **<u>cor</u>**relation measure (CORT). The y-axis shows the number of individuals who were diagnosed with the disease at a certain age, divided by the total number of people having that disease. Values were calculated by taking the median value of 100 permutations of 10,000 people in the UKBB (see Methods). The x-axis shows the age-of-onset in years. Each line denotes one disease and is colored by disease categories. The heatmap in the right upper corner shows the percent overlap between categories and clusters. Numbers give the % of an age-of-onset cluster belonging to each category. Supplementary Figures S10-19 shows the distributions for each disease separately.

### Diseases in the same age-of-onset cluster show higher genetic similarity

Using linear mixed models implemented in BOLT-LMM^23^, we performed GWAS for case versus control on each disease separately and included approximately 10 million common variants that pass quality control and have **<u>m</u>**inor **<u>a</u>**llele **<u>f</u>**requency (MAF) > 0.01 (see Methods)^24,25^. Considering associations with the literature-standard p-value lower than 5 x 10^−8^ as significant^26,27^, we next quantified the associations for each disease, category, and age-of-onset cluster (Figure S21). The **<u>m</u>**ajor **<u>h</u>**istocompatibility **<u>c</u>**omplex (MHC) region is excluded from all analyses, as in the literature, because of its unusually high effect sizes and **<u>l</u>**inkage **<u>d</u>**isequilibrium (LD) patterns (chr6: 28,477,797 - 33,448,354)^28,29^. Out of 116 diseases, 36 had no significant association and the total number of polymorphic sites with at least one significant association was 93,817. The maximum number of significant associations per disease was 35,001 (hypertension) and the median and mean were 13.5 and 1389.3, respectively. We also checked if diseases from different age-of-onset clusters vary in the number of associations. Cluster 4 had hardly any significant associations (the disease with the maximum number of associations had only 3 significant variants). Although cluster 1 had the highest number of significant associations on average, the values across clusters 1, 2, and 3 were not significantly different (Figure S21b). Moreover, endocrine, immunological, cardiovascular diseases had the highest number of associations and infections had the lowest (Figure S21c). Only 1% of the significant polymorphisms (n=932) were in coding regions, and of these 49% (n=452) were missense and only 1% (n=10) were nonsense. We further found that 47% of significant variants (n=43,810) were associated with multiple diseases, but only ∼9% were associated with multiple diseases from different categories (n=8,048) and again ∼9% with different age-of-onset clusters (n=8,801) (Figure S22).

We next sought to characterize the genetic similarities between diseases using a score that shows the excess of overlapping associations between diseases, given the number of significant associations for each disease (see Methods). Importantly, we calculated genetic similarities between 80 diseases that have at least one significant association, excluding the pairs that are vertically connected (*i*.*e*. ancestors to child) in the disease hierarchy (*e*.*g*. essential hypertension and hypertension). We found 47 significant overlaps and diseases with similar age-of-onset profiles showed a higher genetic similarity, even when controlled for disease categories and co-occurrences (F-test p=1.19 x 10^−8^, Figure 2a-b). Moreover, this trend was reproducible when each cluster was analyzed separately (Figure S24). While correcting for the disease categories and co-occurrences, some true positive signals may be removed from the analysis. However, this correction is necessary, as we used the same cohort for multiple diseases and, thus, diseases that co-occur use the same set of samples. Nevertheless, we retained a significant signal even after this correction, demonstrating that diseases with a similar age-of-onset profile show increased genetic similarity compared to those with different profiles, suggesting shared genetic associations (Figure S1).

**Figure 2:**
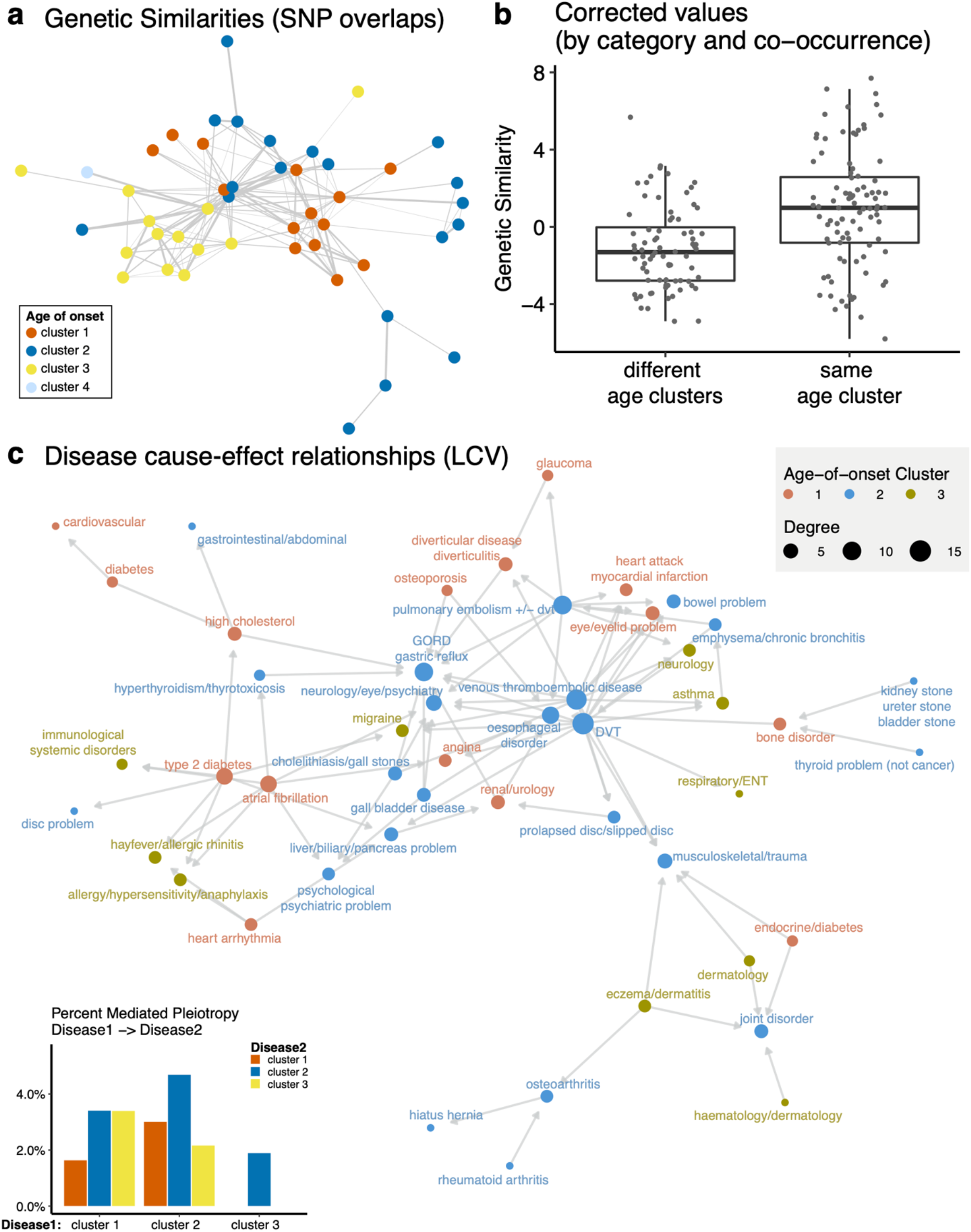
(a) Network representation of the genetic similarities calculated by the overlaps between significantly associated SNPs between diseases. Nodes (n=47) show diseases with a significant genetic similarity to at least one disease and are colored by the age-of-onset cluster. Edges (n=167) are weighted by the genetic similarity corrected by disease categories and co-occurrences. (b) The difference between genetic similarity within and across the age-of-onset clusters. The y-axis shows genetic similarity corrected by category and co-occurrence (raw values are available in Figure S23). The x-axis groups similarities into different or same age-of-onset clusters. c) Network representation of the causal relationships between diseases calculated using LCV. Each node (n=48) shows a disease, colored by the age-of-onset cluster. Size of the nodes represent the number of significant causal relationships between diseases, including both in and out degrees. Arrows show the causal relationship between pairs with FDR corrected p≤0.01 and GCP>0.6. The inset bar plot shows the percent significant causal relationships among all possible pairs (y-axis) between disease 1 (x-axis) and disease 2 (bars colored by the age-of-onset).

We repeated the analysis using 1,703 previously defined LD blocks^30^ instead of considering all **<u>s</u>**ingle **<u>n</u>**ucleotide **<u>p</u>**olymorphisms (SNP) as independent. There was no significant genetic similarity between diseases from different age-of-onset clusters (Figure S25) and the similarities within the same age-of-onset cluster were not explained by the disease categories (p=0.89) and co-occurrences (p=0.15). We further confirmed the results using **<u>h</u>**igh-**<u>d</u>**efinition **<u>l</u>**ikelihood (HDL) inference of genetic correlations, which makes use of the genome-wide data instead of the significant associations and also considers LD^31^. Similarity scores calculated by SNP overlaps and HDL regression were significantly correlated (ρ = 0.75, p<2.2e-16), and the similarities between diseases in the same age-of-onset cluster was significantly higher than that of in different age-of-onset clusters (Figure S26), confirming the conclusion that diseases with similar age-of-onset profiles show higher genetic similarity using another methodology.

Lastly, we checked whether economic status or diet is associated with our results and whether that might drive the commonality between diseases. We found no association between the list of shared SNPs involved in multiple diseases or multiple disease categories and SNPs associated with the “Townsend deprivation index” or any type of special diet, except for gluten-free diet^32^. There was a significant overlap between SNPs associated with gluten-free diet in UKBB and SNPs associated with diseases spanning multiple categories in the age of onset clusters 1 & 2, and 1 & 2 & 3. If two traits are associated, we would see an increased overlap as the p-value cutoff is more stringent; however, this was not the case for any of the traits, including the gluten-free diet (Figure S27).

### Mediated pleiotropy does not explain higher genetic similarities within age-of-onset clusters

We next asked if mediated pleiotropy, rather than a common etiology (Figure S1), may explain higher similarity within age-of-onset clusters. Using a recent methodology developed by O’Connor & Price, we tested for partial or fully causal relationship between diseases^33^. In particular, the method identifies if a **<u>l</u>**atent **<u>c</u>**ausal **<u>v</u>**ariable (LCV) mediates the genetic correlation between diseases. Using a genetic causality proportion, it assigns a causal relationship if one of the diseases is more strongly correlated with the LCV. The authors report that, unlike mendelian randomization, this method can distinguish between the correlation due to common etiology and causation. We tested for potential causation between 60 diseases, excluding the ones with less than 10 significant genetic variants and low heritability estimates (Z_h_<7)^33^. Also, similar to genetic similarities, we did not calculate the causation between diseases that are vertically connected in the disease hierarchy. Following the same significance criteria proposed in the methods article (FDR corrected p≤0.01 and mean **<u>G</u>**enetic **<u>C</u>**ausality **<u>P</u>**roportion (GCP)>0.6), we found significant evidence for full or partial genetic causality in 91 disease pairs between 48 out of 116 diseases in our analysis (Figure 2c, Table S2). Using Fisher’s exact test, we tested if mediated pleiotropy was more common between diseases in certain age-of-onset clusters but did not find any significant difference (FDR corrected p>0.1 for all comparisons, inset bar plot in Figure 2c). Thus, although we detected mediated pleiotropy between some diseases, higher genetic similarities within the same age-of-onset clusters (Figure 2a-b) were not explained in this way and were more likely to be driven by common etiologies.

We also investigated the diseases with the highest involvement in mediated pleiotropy. **<u>D</u>**eep **<u>v</u>**ein **<u>t</u>**hrombosis (DVT) (n=14), venous thromboembolic disease (n=13), and pulmonary embolism (n=9) had the highest number of *out* degrees, meaning they were found as causal for multiple diseases, including all 3 age-of-onset clusters and 5 different disease categories. **<u>G</u>**astro-**<u>o</u>**esophageal **<u>r</u>**eflux (GORD)/gastric reflux (n=10) and esophageal disorder (n=8), on the other hand, had the highest number of *in* degrees, meaning there are multiple diseases detected as causal. These causal diseases spanned 5 disease categories and age-of-onset clusters 1 and 2.

### Genes associated with the age-dependent disease clusters overlap with known longevity modulators and independent aging-related traits

We next mapped all variants to genes based on proximity or known **<u>e</u>**xpression **<u>q</u>**uantitative **<u>t</u>**rait **l**oci (eQTLs) using the GTEx (v8) eQTL associations^34^ (see Methods). We first analyzed the overlap between diseases and tissue-specific eQTLs (Figure S28). The results included tissue-disease pairs that were plausible, such as liver and high cholesterol. However, overall, the number of tissue-specific eQTLs were biased due to the varying number of samples in each tissue and thus, we used eQTLs in all tissues to map SNPs to genes. We confirmed that the SNPs associated with diseases spanning multiple disease categories have eQTLs in multiple tissues (Figure S29). To assess the reproducibility of the genes identified, we compared the significant hits with all those reported in the GWAS Catalog. We verified that most of the diseases had significant overlaps with the same or associated traits in the GWAS-Catalog (*e*.*g*. hypertension and blood pressure or osteoporosis and bone density), confirming that our results were reproducible with independent data (Table S3).

We next compiled the genes associated with multiple diseases and multiple categories and grouped them based on the age-of-onset cluster of the associated diseases (Table S4). In particular we created two sets of genes, ‘*multidisease’* and ‘*multicategory’*, for clusters 1, 2, and 3. We excluded cluster 4 because the number of variants significantly associated with this cluster was low (n=7 associated with 5 diseases), mapping to only 3 genes (*ZPBP2, NPC1L1, TARID*). We also compiled genes associated with multiple diseases or categories in combinations of different age-of-onset clusters. Importantly, genes associated with multiple clusters are not in the gene sets for individual clusters as the latter only include the genes specific to individual clusters, *i*.*e*. cluster 1, cluster 2 and cluster 1 & 2 genes included all mutually exclusive sets.

Using these lists, we sought to understand if the common genes between diseases with the same age-of-onset profile had previously been associated with aging. We compared the *multidisease* and *multicluster* gene lists with the literature-based aging databases: GenAge human (genes associated lifespan in humans or closely related species), human orthologs in GenAge model organism (genes modulating lifespan in model organisms), CellAge (genes regulating cellular senescence), DrugAge targets (drugs modulating lifespan in model organisms), and all databases combined^35–37^ (Figure 3a). In general, genes associated with clusters 1 and 2, but not Cluster 3, showed significant enrichment with known aging-related genes. The list of overlapping genes is given in Table S5. The CellAge database showed the significant overlaps with genes associated with clusters 1, 2, and ‘1 & 3’. DrugAge targets had a significant overlap with clusters 1, 2, and ‘1 & 2’. GenAge Human only had significant association with genes associated with cluster ‘1 & 2’. GenAge model organism data significantly overlapped with genes associated with all clusters (1 & 2 & 3). And the combination of all aging-related gene sets showed significant overlaps with clusters 1, 2, 1 & 2, and 1 & 2 & 3. In conclusion, although the association is established through a small subset of genes as also reported in the literature^17,18^, the clusters 1 and 2, constituting age-dependent profiles, shared a significant genetic component with known longevity- and senescence-modulators, while cluster 3 did not. We further confirmed the aging relevance of our cluster 1 and cluster 2 genes using independent GWAS results for ‘aging’ and ‘longevity’ traits as well as Alzheimer’s, Parkinson’s, and age-related macular degeneration diseases which are not represented in our data (Figure S30). The significant overlaps include longevity associated *APOC1, APOC1P1, TOMM40*, and aging associated *APOE, CHRNA3, DEF8, LINC01239, TRIM59* genes, some of which also overlap with Alzheimer’s, Parkinson’s and age-related macular degeneration.

**Figure 3:**
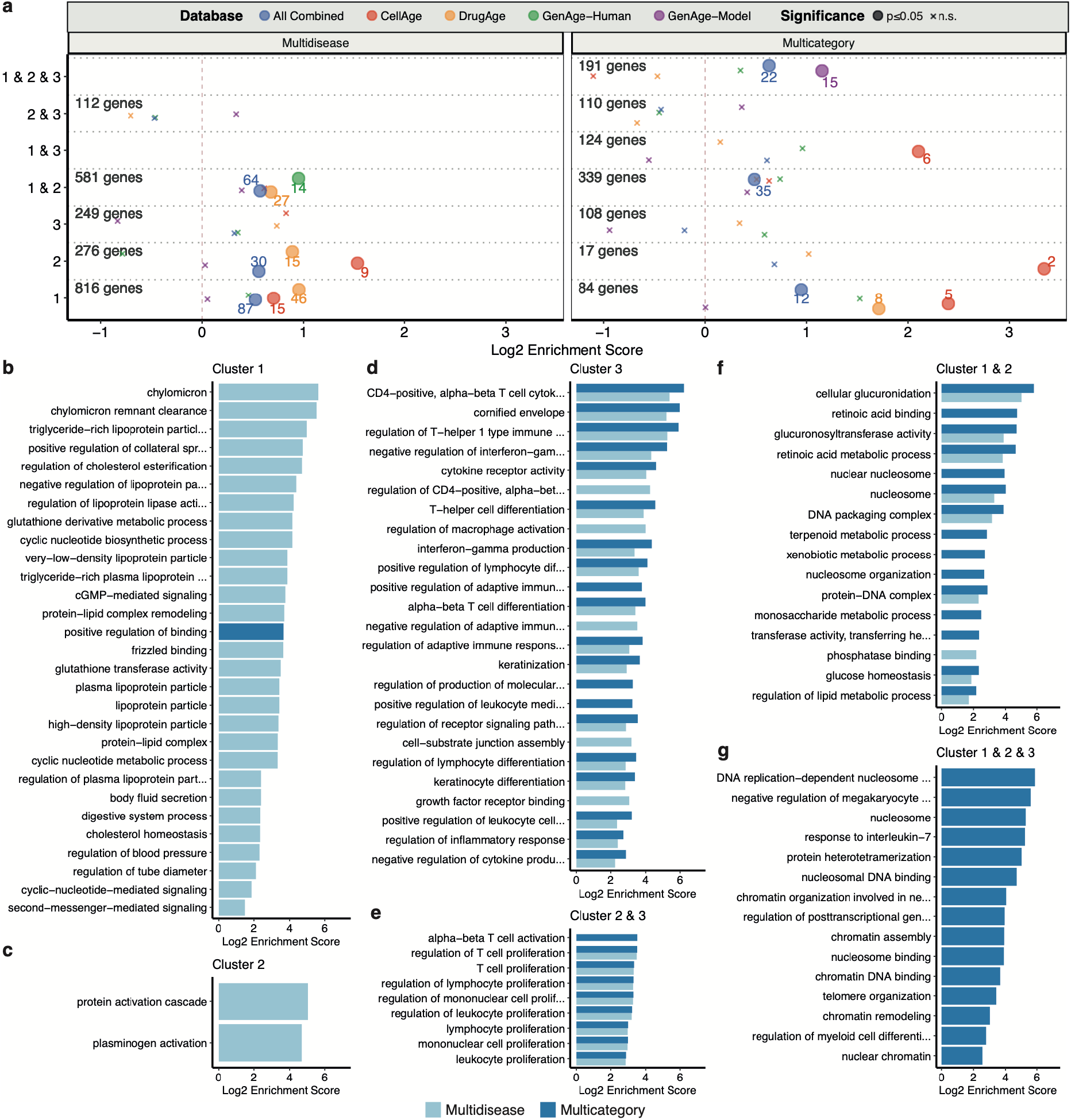
a) Overlap between known aging-related genes in databases and genes associated with diseases in different clusters. The x-axis shows log2 enrichment score, and the y-axis shows the age-of-onset clusters. The numbers of genes in each cluster (for both Multidisease and Multicategory genes) are given. The size of the points shows the statistical significance (large points show marginal p- value≤0.05, small ‘x’ indicate non-significant overlaps) and the color shows different databases. The colored numbers near the points show the numbers of overlapping genes. b-f) Gene Ontology (GO) Enrichment results for genes associated with diseases in b) Cluster 1, c) Cluster 2, d) Cluster 3, e) Cluster 2 & 3, f) Cluster ‘1 & 2’, g) Cluster ‘1 & 2 & 3’. Representative GO categories for significantly enriched categories (BY-adjusted p-value ≤ 0.05) are listed on the y-axis (see Methods). Log2 enrichment scores are given on the x-axis. The color of the bar shows the result for multidisease and multicategory genes. There was no significant enrichment for cluster 1 & 3.

### Age-related changes in expression and methylation of the genes associated with diseases

To further explore the association between the disease-associated SNPs and ageing, we analyzed the age-related expression and methylation profiles of the genes associated with SNPs identified in our study (Figures S31-33). Using tissue-specific eQTL-gene associations, we first checked whether the disease-associated SNPs might induce gene expression changes that mimic the changes that occur at the age of 40, which might explain the age-of-onset distribution of the first cluster. The multicategory cluster 1 eQTLs that increase the expression of genes indeed mimicked the age-related expression changes (Figure S31, expression difference is positive at the age of 40 and is significantly different from 30 and 50). They were enriched in telomere maintenance and organization, columnar epithelial cell differentiation, glucose metabolic process, and multicellular organismal signaling (p < 0.01 but n.s. after multiple testing correction). Interestingly, multicategory cluster 2 and 3 eQTLs that decrease the expression of genes also mimicked the age-related changes (Figure S32, expression difference is negative at the age of 40 and is significantly different from 30 and 50). These genes were enriched in immune response-related categories (Table S6). Thus, the overall picture is complex and requires further investigation, but it suggests age-related increases in the expression of genes associated with multicategory cluster 1 SNPs might influence the age-of-onset curve. Still, immune-related genes that decrease in expression may also contribute to the occurrence of cluster 2 and 3 diseases in late ages.

We next analyzed the association between age-of-onset clusters and methylation by comparing the multidisease and multicategory gene sets with i) DNA methylation (“GO:0006306”, n = 29) and demethylation (“GO:0080111”, n = 20) GO Terms, and ii) differentially methylated genes reported in previously published studies^38,39^. Although DNA demethylation category included none of the multidisease or multicategory genes, the DNA methylation category had 4 overlapping genes. GNAS and HELLS are multidisease cluster 1 genes, and GATAD2A is a multicategory cluster 1 gene. Disruption in the ortholog of HELLS gene in mice (Hells) was found associated with premature aging^35,40^. The fourth gene, CTCF, is a multidisease cluster 2 gene with no known association with aging. Although the overlap is only specific to clusters with age-dependent profiles, these GO terms were not significantly enriched in the disease-associated genes (Figure 3). Next, using the published results of two external methylation data analyses^38,39^, we found a significant overlap between the genes that show differential methylation with age and the clusters with age-dependent profiles, clusters 1, 2 and 1 & 2 (Figure S33). Overall, although there was no significant association with the regulators of DNA methylation, we found age-related changes in gene expression and methylation for the late-onset clusters.

### Genes associated with different age-of-onset clusters have different functions

**<u>G</u>**ene **<u>o</u>**ntology (GO) enrichment analyses were applied to the gene lists, including **<u>b</u>**iological **<u>p</u>**rocess (BP), **<u>m</u>**olecular **<u>f</u>**unction (MF) and **<u>c</u>**ellular **<u>c</u>**omponent (CC) categories. Cluster 1 was associated with many lipoprotein-related categories, cellular signaling, and blood pressure (Figure 3b). Cluster 2 showed association to plasminogen activation and protein activation cascade (Figure 3c). Cluster 3 and clusters ‘2 & 3’ had associations to many immune-related categories and cell adhesion (Figure 3d, e). Genes in clusters ‘1 & 3’ did not have any significant associations. Genes associated with cluster ‘1 & 2’ were related to nucleosome complex, glucose homeostasis, retinoic acid binding (Figure 3f). Genes associated with at least one disease in all clusters (‘1 & 2 & 3’) showed association with interleukin-7 response, differentiation, telomere as well as nucleosome complex (Figure 3f). Since cluster 3 did not have an age-dependent profile, the association with nucleosome complex could represent pleiotropic genes in general. Here we listed the categories that are representative to all other significant functional groups. The full list is given in Table S7, and the procedure of selecting representatives is described in the Methods. Overall, these results suggest that, although cluster 1 and cluster 2 genes were both linked to previously identified aging-related genes, they have distinct functional profiles.

### Evolution of aging and age-related diseases

Lastly, we sought to understand the abundance of disease-associated variants in the population and their relationship with the evolutionary theories of aging. **<u>M</u>**utation **<u>a</u>**ccumulation (MA) theory explains the functional decline at later ages by relatively lower selection pressure on deleterious germline variants that are functional at later ages. Accordingly, we first hypothesized that SNPs associated with later-onset diseases (Cluster 1) would have a higher frequency than the SNPs associated with diseases that occur at earlier ages (Clusters 2 and 3), which are presumably under stronger selection pressure. Not finding any difference in the power to detect SNPs for different clusters by comparing median MAF and number of cases (Figure S34), we compared the allele frequencies associated with different age-of-onset clusters^41^. As SNPs which are close together in the genome are expected to have similar allele frequencies due to linkage, we calculated the median risk allele frequency for SNPs within previously defined LD blocks^30^. Supporting the MA, diseases of cluster 1 had significantly higher risk allele frequencies than cluster 2, both for the SNPs associated with one disease (Figure 4a, Wilcoxon test p=0.00033) or with one cluster (Figure 4b, Wilcoxon test p=0.0068, also confirmed by bootstrapping n=100 loci for B=1,000; Figure S35). We further confirmed that this trend is not specific to the UK population, as we obtained comparable results in all super-populations of the 1000 Genomes Project^42^ (Figure S36-37). Variants associated with cluster 3, which includes immune-related diseases, were not significantly different from those associated with cluster 1, although cluster 3 diseases can occur even at an earlier age. Moreover, although the difference in the median allele frequencies was not significant, the shapes of distributions were different, with a significant shift towards higher risk allele frequencies only in cluster 1 (p_cl1_=0.05, p_cl3_=0.86 calculated using 10,000 permutations). High minor allele frequencies, thus a higher variation we observed in cluster 3 is in line with the previous suggestion that immune-related genes are under long-term balancing selection in humans^43^, although positive selection also influences immunity^44–46^.

**Figure 4:**
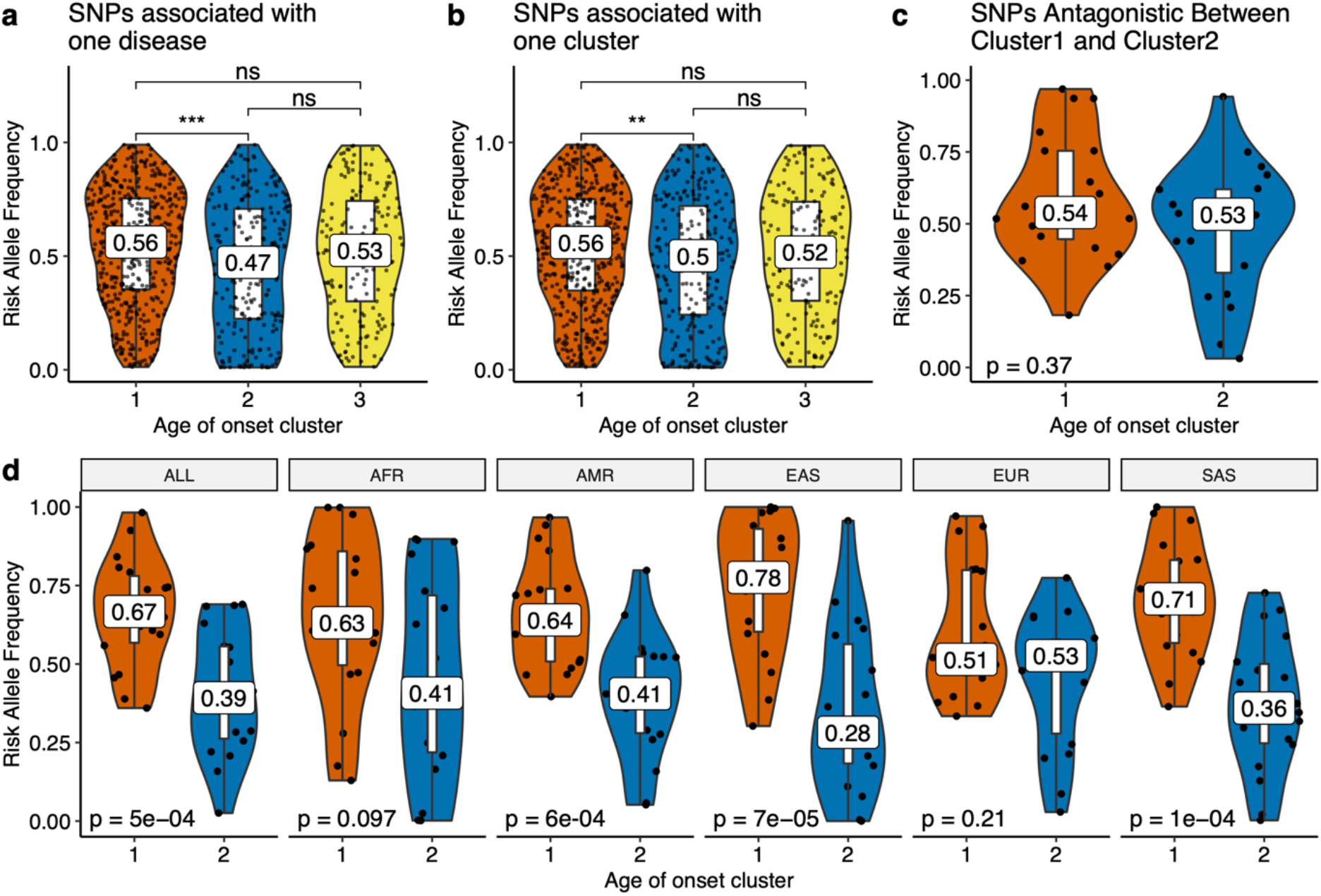
Risk allele frequency distributions (y-axis) for different age-of-onset clusters (x-axis) in the UKBB for a) SNPs associated with one disease (excluding antagonistic associations), b) SNPs specific to one cluster (excluding antagonistic associations) (ns: p>0.05, *:p≤0.05, **: p≤0.01, ***: p≤0.001, ****: p≤0.00001), and c) SNPs that have antagonistic association with cluster 1 and 2 (excluding agonists between cluster 1 and 2). d) The same as panel c but for different 1000 Genomes super-populations (ALL: complete 1000 Genomes cohort, AFR: African, AMR: Ad Mixed American, EAS: East Asian, EUR: European, SAS: South Asian).

To test the **<u>a</u>**ntagonistic **<u>p</u>**leiotropy theory (AP), we first asked if the diseases with different onsets have an excess of antagonistic SNPs. Similar to a previous study^41^, we defined a pleiotropic biallelic SNP as *agonistic* if the risk allele is the same for different diseases, and as *antagonistic* if opposite alleles are associated with increased risk for different diseases. If one of these diseases is under a stronger negative selection, then the risk allele of the other disease could increase over time. Comparing the proportion of agonist and antagonist SNPs within and between the age-of-onset clusters, we found that there is an excess of antagonistic pleiotropy between diseases with different age-of-onset profiles (Fisher’s exact test p<0.001, Table S8). Next, we tested the differences in risk allele frequencies between the clusters, as AP predicts a higher risk allele frequency for late-onset diseases. The risk allele frequencies for antagonistic associations between cluster 1 and 3 or 2 and 3 did not show any significant difference in the UKBB or 1000 Genome super-populations (Figures S38-39). Interestingly, the difference between the risk allele frequencies for cluster 1 and cluster 2 was also not significant for the UKBB population (Figure 4c). However, all 1000 Genome super-populations except for Europeans had higher risk allele frequencies for cluster 1 diseases (Figure 4d). We hypothesized that this is mainly due to increased power when testing the antagonistic associations with frequency closer to 0.5. We thus investigated the allele frequency differences for the significant variants with increased effect sizes. Indeed, associations with a larger effect size showed the expected differences in allele frequencies, although the number of independent loci was limited (Figure S40). We also examined the type of diseases and genes associated with antagonistic pleiotropy. The main driver of the pattern was the loci with *ABCG8* and *ABCG5* genes, showing antagonistic relationship for high cholesterol (cluster 1) and other lipid-related diseases in cluster 2, such as gallbladder disease and cholelithiasis. Another locus included variants that show antagonistic relationship with cardiovascular disease (cluster 1) and the cluster 2 diseases gout (*ADH1B*), osteoarthritis and joint disorder (*SLC39A8*), and osteoarthritis (*BANK1*). Another potential candidate was a locus associated with hypertension (cluster 1) and musculoskeletal diseases (cluster 2), but this locus included multiple candidate genes (Table S9). Nevertheless, our comparison is between common diseases that occur after the age of 20 and 40, which are both after the average age at first reproduction and therefore the start of the decrease in the force of natural selection^47^. Thus, a better comparison would include the mutations causing rare developmental diseases, which are not available in the UKBB.

## Discussion

The number and the incidence of diseases increase with age. In this study, we explored whether this results from a common genetic component among ARDs, which might also be linked to aging. We compared genetic associations and age-of-onset distributions of 116 self-reported diseases in the UKBB and found shared variants, genes and pathways, which were also associated with aging.

Using an unsupervised, data-driven approach to classify diseases based on their age-of-onset profiles, we found 4 main clusters; i) diseases that rapidly increase after 40 years of age, ii) diseases that increase after 20 years of age, iii) diseases with no age-related pattern, and iv) diseases that peak at around 10 years of age. Notably, unlike previous studies^18,48^, by using this unsupervised approach, we detect a distinction between cluster 1 and cluster 2, which both show age-dependency but distinct age-of-onset distributions. These two clusters were associated with genes with different functional and evolutionary characteristics, although they both overlap with known aging-related genes. Moreover, germline mutations may influence late-onset diseases through affecting the function of genes that show differential expression or methylation with age.

Based on genetic associations, diseases with similar age-of-onset profiles showed a higher genetic similarity on average, compared to diseases in other clusters, even when controlled for disease categories and co-occurrences (Figure 2a-b). Moreover, this similarity within age-of-onset clusters was not explained by mediated pleiotropy, in which one of the diseases is causal for the other one, suggesting instead a common etiology. We then studied the genes involved and found that genes associated with clusters 1 and 2 (both constituting ARDs) are enriched with known longevity- and senescence-modulators, while genes associated with cluster 3, which does not show an age-dependent profile, did not show this enrichment. In addition, we found that genes associated with different age-of-onset clusters have different functions. Comparing the risk allele frequencies of variants associated with different age-of-onset profiles, we found support for both mutation accumulation and antagonistic pleiotropy theories of aging, although the number of independent loci supporting the second was limited. Lastly, we provide a list of drugs that target the common genetic component between ARDs, which may in the future be tested for their effects on multimorbidity and polypharmacy associated with late life.

In this study, we had a limited age range, covering individuals up to 65 years old and thus, could not analyze diseases of later ages. Neither did we consider the cancers or changes in regulation of gene expression for the same cohort, which are affected not only by aging, but also various environmental or intrinsic factors. Future cohorts with a broader age range and spanning multi-omics data, somatic mutations, health outcomes, and lifestyle information, will enable a better understanding of the genetic mechanisms of age-of-onset determination and establishing the causal link with candidate genes. Despite these limitations, we present a novel approach to study ARDs using an unbiased, data-driven approach and show that ARDs share common genetic associations linked to aging. Our results suggest that targeting the common pathways between multiple ARDs could offer compression of late life multimorbidity as well as alleviating the effects of polypharmacy.

## Data Availability

The full set of GWAS results from this study can be accessed using BioStudies (S-BSST407) and all results generated in the analysis are provided as Supplementary Datasets and Tables.

https://www.ebi.ac.uk/biostudies/studies/S-BSST407

## Software Availability

All the code used to perform analyses is available in GitHub: https://github.com/mdonertas/ukbb_ageonset

## Ethics Statement

### Author Contributions

H.M.D conceived and designed the study with contributions from L.P. and J.M.T.. H.M.D analyzed the data with the help of D.K.F and M.F.V.. H.M.D. interpreted the results and wrote the manuscript with contributions from all authors. All authors read, revised and approved the final version of this manuscript.

## Acknowledgment

This research has been conducted using the UK Biobank Resource (application no. 30688). The authors thank the GWAS-Catalog team for providing the list of studies using UK Biobank data; James Stephenson and Roman Laskowski for their help in running VarMap tool; and Mehmet Somel, Susan Ozanne, Pedro Beltrao, and Wolfgang Huber for fruitful discussions. H.M.D. is a member of Darwin College, University of Cambridge.

## Methods

### UK Biobank Data

Data was downloaded using bash and following the guidelines provided by the UK Biobank.

#### Sample quality control

After excluding all samples from individuals who have withdrawn their data from UK Biobank, we first filtered out all samples without genotypes (N = 14,248). Then, we used the following criteria for the remaining 488,295 samples.

#### Discordant sex

Data includes two entries for sex: 1) self-reported and 2) genetic sex determined using the call intensities on sex chromosomes. There are multiple reasons why these two entries may not correspond, such as sample mishandling, errors in data input, transgender individuals, and sex chromosome aneuploidies^1,2^. Since we used sex as a covariate in our GWAS model, we preferred to be cautious about this issue and excluded all cases where the genetic sex and self-reported sex did not correspond and all cases where sex chromosome aneuploidy was detected. Specifically, we used the fields ‘31-0.0’ (Sex) and ‘22001-0.0’ (Genetic sex) to compile discordant information. There were 235 self-reported males being identified as female by the genetics, and 143 self-reported females being identified as males by the genetics. We excluded these 378 cases, 0.077% of the data. Moreover, field ‘22019-0.0’ (Sex chromosome aneuploidy) is used to exclude cases with sex chromosome aneuploidy. There were 651 cases of aneuploidy, 0.133% of all data. 181 of these cases (27.80% of aneuploidy cases) were also detected as discordant information in the first step. This corresponds to 47.88% of discordant sex cases. Overall, we identified 848 samples to be excluded based on this criterion.

#### Genotype call rate & Heterozygosity

Genotype missingness and heterozygosity are widely used as a measure of DNA sample quality. For quality filtering based on missingness and heterozygosity we only used the suggested exclusions by UK Biobank. Specifically, we used the field ‘22010-0.0’ (Recommended genomic analysis exclusions) and determined the cases with ‘poor heterozygosity/missingness’ (N = 469). We next used the field ‘22018-0.0’ (Genetic relatedness exclusions) and noted down the cases with ‘Participant self-declared as having a mixed ancestral background’ (N = 692), and the cases with ‘High heterozygosity rate (after correcting for ancestry) or high missing rate’ (N = 840). Lastly, there were 968 cases that are suggested as outliers for heterozygosity or missing rate, field ‘22027-0.0’ (Outliers for heterozygosity or missing rate). We then checked the scatter plots for logit(Missingness) vs. Heterozygosity for each Ethnic Background, in accordance with the identification of samples to exclude by the UK Biobank^2^ (Figure S43). Logit transformation is used to linearize sigmoidal distribution of missingness. Investigation of heterozygosity can detect DNA sample contamination, inbreeding, or mixed ethnicity^1^. This quality check reveals when people with a mixed ethnicity tend to have a higher heterozygosity, even after correcting for PCs. We confirmed these are in accordance with the original article and excluded the samples suggested by the UK Biobank.

Overall, there were 3,697 samples excluded based on these two criteria. Please note that the numbers presented above may not add up to this number, because there were some samples excluded based on multiple criteria. The percent overlap across multiple criteria is given in Figure S44.

#### Preparing the trait data

Using the samples that passed the quality control (N=484,598), we subsetted the data so that it included only the baseline visit. Apart from the data that is already available in UK Biobank, we calculated some other values: *1) BMI:* Using the columns for ‘Weight’ and ‘Standing height’ we calculated BMI as: Weight / (Standing Height / 100)^2^, *2) Parent Age at Death - Minimum:* The youngest age at which either parent died. *3) Parent Age at Death - Maximum:* The age of death for the parent who lived longest. *4) Parent Age at Death - Average:* The average age of death for the two parents. If neither of the parents died, or if the data was unavailable, these values (2-4) were set to be NA. If only one parent died, we use the corresponding age as both the minimum, maximum, and average. *5) The number of self-reported non-cancer diseases:* The number of unique self-reported non-cancer illnesses each participant recorded in the baseline recruitment. *6) The number of self-reported cancers:* The number of unique self-reported cancers each participant recorded in the baseline recruitment. *7) Self-reported diseases after taking the disease hierarchy into consideration (Propagated disease data):* The self-reported diseases in UK Biobank are not independent, but rather are organized in a hierarchical manner. Using the relationship information between diseases, we propagated disease-participant associations, upwards, including terms higher up the tree. For example, if a person reports having “essential hypertension”, we also annotate that person with “hypertension”, and “cardiovascular disease”. *8) Age at diagnosis for the self-reported diseases after taking the disease hierarchy into consideration (Propagated age at diagnosis data):* We re-defined age at diagnosis using the minimum age at diagnosis for all the diseases that were child term for a particular disease in the disease hierarchy. *9) The number of self-reported non-cancer diseases after taking the disease hierarchy into consideration (Propagated number of non-cancer diseases):* The number of unique self-reported diseases each participant records after taking into account the data propagation. *10) Age when the last deceased person died:* We calculated the age of each person when the last death entry in the UKBB happened. This value is used to calculate the proportion of people who died at a certain age interval in Figure S2c.

#### Selecting diseases to analyze

We calculated the disease occurrences for all self-reported diseases in UK Biobank. Specifically, among the cohort we used, we calculated how many participants and what proportion of males and females reported each disease. Since we analyzed the same set of SNPs that have MAF>=0.01 across multiple diseases, to decrease the false positive rate in GWAS, we limited the diseases to a subset with at least 2,000 cases (n = 129 out of 472)^3^. Moreover, we only focused on diseases that were common and not sex-limited, *i*.*e*. we only considered diseases that are seen in 1 in every 1,000 males and females (n = 189 out of 472). The intersection of these two conditions was 116 diseases and we excluded all others.

We only analyzed self-reported non-cancer diseases (field ‘20002’) and did not combine self-reported cancers (field ‘20001’), mainly because i) the number of cases is low (45,224 compared to 384,906 for other diseases), ii) cancer is thought as a result of a complex interaction between germline and somatic mutations^4,5^, whereas the evidence for the effect of somatic mutations in other diseases is limited to rare and neurological disorders^6,7^, iii) the relationship between cancer and aging is complex, *e*.*g*. while telomere attrition and cellular senescence are thought to be evolved as a tumor suppressor mechanisms; aging-related changes in epigenomic landscape and genomic instability contribute to cancer occurrence^8^. Thus, although a similar analysis using cancers would be interesting, we only focused on non-cancer self-reported diseases in this study. Since we did not exclude the individuals with cancer, we also checked if there is a significant overlap in individuals with cancer with the other diseases we analyzed (Figure S45). However, there was no such association.

### Disease co-occurrence calculations

#### Relative risk (RR) score

Relative risk is an estimate of having the disease A, when already affected by disease B. Overall it measures if disease A co-occurs with disease B more frequently than expected if these diseases were independent in the population. It is calculated as a fraction between the number of patients diagnosed with both diseases and a random expectation based on disease prevalence^9^. Mathematically it can be expressed as follows, using the values from Table 1:

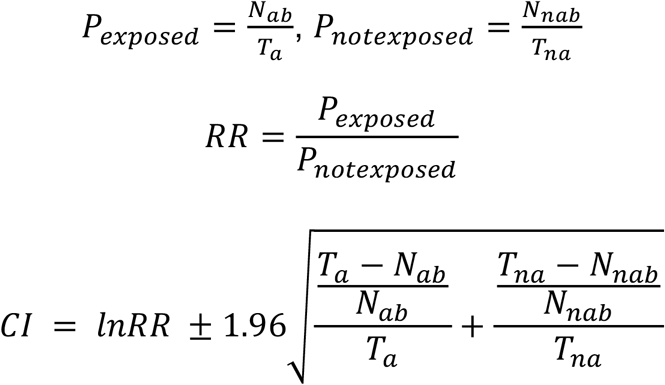

**Table 1:** Contingency table for disease comorbidities.

|  | Disease B | No disease B | Total |
| --- | --- | --- | --- |
| Disease A | $N_{ab}$ | $N_{anb}$ | $T_a$ |
| No disease A | $N_{nab}$ | $N_{nanb}$ | $T_{na}$ |

#### ϕ value (Pearson correlation for binary variables)

The ϕ value measures the robustness of the association between diseases based on co- occurrences^10^. Mathematically, it can be expressed as:

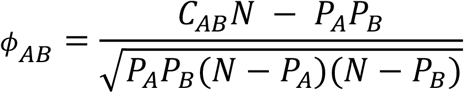

*N*: the total number of individuals

*P*_*A*_: Prevalence of disease A

*C*_*AB*_: Number of patients with both diseases

ϕ ranges between -1 and 1, where the sign indicates the type of association.

### Disease age-of-onset

#### Disease dissimilarity measure

##### Temporal correlation

In order to calculate dissimilarities among diseases, we use CORT^11^ distance as included in R package TSclust^12^. Euclidean distance and dynamic time warping^13^ are the two most widely used proximity measures for time series proximity. However, they are both calculated based on the closeness of the values and disregard the growth behavior. Correlation-based measures are also used to calculate the similarity between time series. However, Pearson correlation overestimates the similarity because of the underlying temporal dependency and Spearman correlation fails to consider the growth rate as it is based on ranks. Chouakria et al., on the other hand, suggested a measure that also considers the proximity-based on growth behavior, CORT^11^. Temporal correlation between two time series objects S_1_=(u_1_,u_2_,…,u_p_) and S_2_=(v_1_,v_2_,…,v_p_) is calculated as follows:

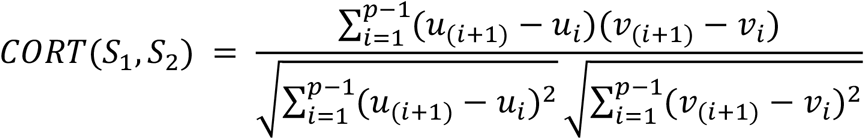

CORT ranges between -1 and 1. A value of CORT = 1 implies that two time series increase or decrease simultaneously with the same growth rate, whereas a value of -1 shows the same growth rate but in opposite direction. If the value is 0, it means there is no temporal correlation between the series.

##### Dissimilarity Index

The dissimilarity index suggested by Chouakria et al.^11^, is calculated based on an automatic adaptive tuning function and considers similarity based on both values and behavior, *i*.*e*. the strength of monotonicity and closeness of the growth rates as calculated by CORT measure introduced in the previous section. They suggest a dissimilarity index D as follows:

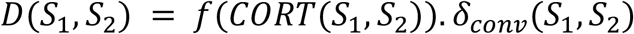

Where *f*(*x*)is an exponential adaptive tuning function:

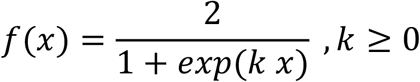

As k increases, the contribution of behavior increases. We use k = 2 and as a result behavior (CORT) contributes 76.2% to D and values (*δ*_*conv*_) contribute 23.8%. For *δ*_*conv*_ we used conventional Euclidean distance.

##### Clustering diseases by age-of-onset

We clustered data using ‘partition around medoids (PAM)’ algorithm^14^ based on the distance measure calculated using the previous step. The aim of this algorithm is to minimize the average distance (based on any dissimilarity measure) between the objects and their closest selected medoid object. It works very similarly to k-means, except instead of defining arbitrary points as the means, it defines medoids among the objects. Thus, it can incorporate any distance measure instead of just using the mean distance between points (*i*.*e*., euclidean distances). The algorithm first searches for k number of objects that represent the structure of the data (Here the number k is assumed to be known a priori but see the next section for the determination of k). After finding a set of k medoids, k clusters are constructed by assigning each observation to the nearest medoid. Overall, the goal is to find k representative objects such that the sum of dissimilarities of the observations to their closest representative is as small as possible. After each assignment, medoid and non-medoid data points are swapped and a cost (sum of distances of points to the new medoid) is calculated. If the total cost of configuration is decreased, then the new configuration is maintained, otherwise, it is reversed. We used ‘pam’ function in the ‘cluster’ package^15^ in R to apply this algorithm.

##### Choosing the optimum number of clusters

The clustering algorithm we used, PAM, clusters data into k clusters, which is determined by the user. So, even if there is no real structure in data, as we increase the number of clusters, we can get more and more clusters. A potential way to decide on the number of clusters is using the gap statistic^16^. This value is calculated by comparing the logarithm of the within-sum-of-squares (WSS) to averages from simulated data without any structure.

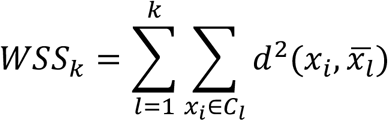

*k*: number of clusters

*C*_*i*_: objects in the l-th cluster

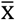: the average point.

Calculating only WSS, however, is not enough as it would be minimized when each point has its own cluster. Thus, we use the gap statistic which suggests calculating *log*(*WSS*_*k*_)for a range of values of k and compare it to that obtained by WSS calculated based on simulated data. So, after WSS is calculated for various values of k, the algorithm involves generating B (we choose B=1,000) reference datasets, using Monte Carlo sampling from a homogeneous distribution and re-calculate WSS for all k values. Using these values gap(k) statistic is calculated as:

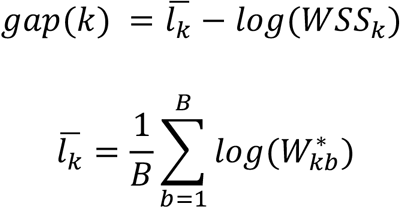

If the clustering is good (i.e. WSS is small) we expect 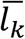 to be higher than log(WSS). Thus, gap statistic is mostly positive and we are interested in the highest value. Tibshirani et al.^16^ suggests using the smallest k such that,

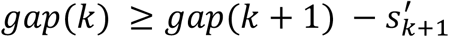

where

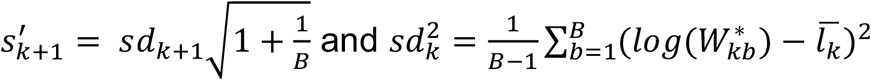

Using this approach, we determined k = 4.

### Genome wide association study

#### Preparing the files required for GWAS

##### Fixing FAM files

In UK Biobank FAM files, the column for ‘phenotype’ includes batch that is coded with characters. In order to use BOLT-LMM^17^, we updated all the entries in this column to numeric values^18^.

##### ‘Remove’ files for BOLT-LMM

BOLT-LMM accepts a list of individuals to be removed from the analysis as an input. These files are called ‘remove’ files and are in the FAM format. We prepared these files for i) withdrawn samples (n = 51), ii) samples that failed the quality control (n = 3,779), iii) samples that have information in PLINK files but lack BGEN files (n = 968).

##### Calculating the SNP statistics

In order to apply a quality filter for SNPs, using PLINK^1^, we calculated i) p-values for each SNP showing whether it deviates from Hardy-Weinberg equilibrium, and ii) Minor allele frequencies (MAF).

##### SNP Quality Control

We excluded SNPs that deviate from Hardy-Weinberg equilibrium (p ≤ 1e-6, n = 202,473) or with a minor allele frequency (MAF) smaller than 0.01 (n = 127,969). In total, we discarded 314,697 SNPs (Note that the numbers do not add up as these SNPs can overlap), resulting in 9,886,868 sites.

#### Phenotype File

We created a phenotype file that can be used as an input for BOLT-LMM, including the following fields: sex, age when attended assessment center, calculated BMI, assessment center, ethnicity, batch, first 20 PCs, and self-reported diseases (one column per disease).

#### GWAS run using BOLT-LMM

For each disease, we run GWAS separately using BOLT-LMM with the following inputs:

- We remove the samples that are in plink files but now in bgen; samples that did not pass our QC; samples from the individuals who have withdrawn their data from the UKBB
- We excluded the SNPs that deviate from Hardy-Weinberg equilibrium, and have minor allele frequency lower than 0.01.
- We used Sex, Age, BMI, assessment center, ethnicity, batch, and the first 20 PCs as covariates.
- To run the mixed-model, a reference LD score table is required. We used LD scores generated using 1000 Genomes European-ancestry samples, which is provided with the BOLT-LMM download.
- Genetic map for hg19 file provided in the BOLT-LMM website.
- We set ‘bgenMinMAF’ argument to 1e-2 and ‘bgenMinINFO’ parameter to 0.5 to only include SNPs that pass these criteria.

#### GWAS Results

We removed MHC region (chr6: 28,477,797 - 33,448,354) from the analysis and considered positions with a p-value lower than 5×10^−8^ as a significant association.

### Coding Variants

We used VarMap^19^ to map variants to proteins and domains. VarMap provides detailed information about coding variants, including annotations for the missense, synonymous, and nonsense variations. In our analysis, if a variant is not annotated as a coding variant in VarMap output, we assumed it is non-coding.

### Genetic similarities between diseases

In order to calculate the overlap between diseases we used the number of SNPs that are significantly associated with both diseases, but corrected by the number that is expected by chance, if two diseases are independent:

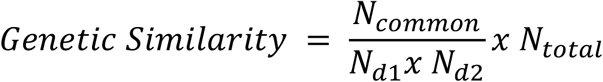

N_common_: Number of SNPs in common.

N_dx_: Number of SNPs associated with disease X.

N_total_: Total number of SNPs analyzed in the study.

The statistical significance of these genetic similarities is calculated using the binomial test, and the similarity is only considered for downstream analysis if p<=0.01. Moreover, the value is only calculated if two diseases do not have any hierarchical relationships in the disease hierarchy.

In order to assess the genetic similarity within age-of-onset clusters, we further used linear regression to correct log2 genetic similarity value by disease co-occurrences (risk ratios) and disease categories (binary data showing whether two diseases are of the same category). The ‘corrected genetic similarity’ is the residuals from this linear model.

#### LD Blocks

In order to assess the similarity between different diseases we use overlaps across significant associations and thus preferred not to do fine mapping. However, a significant challenge is that genomic variations are not independent but instead linked in the genome. To understand the effect of linkage disequilibrium or overcome it, we made use of linkage disequilibrium blocks previously defined for human genome^20^. We repeated the analysis for genetic similarity after collapsing all positions within an LD block and thus creating independent genomic loci (n = 1,703). We use binary information for LD blocks, i.e. blocks with at least one significant association are considered as a hit, and the rest are not.

### High-definition likelihood (HDL) method to calculate genetic similarities

We repeated the genetic similarity analysis with HDL^21^, which is a recent alternative to LDSC to calculate genetic correlations. Using the code available in github (https://github.com/zhenin/HDL/) we calculated the genetic similarities between diseases, using the leading eigenvalues explaining 99 and their correspondent eigenvectors in LD score matrix.

### Comparison between SNPs associated with multiple diseases and economic status or specific diet

We used the UK Biobank GWAS summary statistics provided by Neale Lab^22^, and calculated the overlap between SNPs associated with Townsend Deprivation Index (inverse-rank normalized), or specific diet regimes (i.e. gluten-free, lactose-free, low calorie, vegan, vegetarian, or other as specified in the UK Biobank) and the SNPs associated with multiple diseases or multiple disease categories in each age-of-onset cluster.

### Analysis of mediated pleiotropy between diseases

Using the LCV method developed by O’Connor & Price, we tested the causal relationships between diseases^23^. We used the R functions developed by the authors and provided on GitHub (github.com/lukejoconnor/LCV/). We calculated the genetic causality proportion (GCP) between each disease pair, if the diseases have at least 10 significant variants and a significant heritability estimate as suggested by the developers (Z_h_≥7). We only calculated GCP if the diseases are not vertically connected on the disease hierarchy. Following the criteria applied by the developers, we considered pairs with FDR corrected p≤0.01 and mean GCP>0.6 as significant.

### SNP to gene mapping

We map all SNPs analyzed in GWAS to genes based on proximity and eQTL results.

#### Using proximity

Using VariantAnnotation^24^, TxDb.Hsapiens.UCSC.hg19.knownGene^25^, and GenomicRanges^26^ packages in R, we mapped the genomic coordinates for each SNP to genes. Specifically, if a gene is within the coding region, intron, 5’ or 3’ UTR, or 1kb down- or up-stream of the transcription start site, we annotated that SNP to the gene. As a result, we had 4,443,872 SNP-gene associations for 4,236,176 SNPs and 22,228 Entrez gene IDs. We used the Ensembl biomaRt^27^ package in R to retrieve HGNC symbols (17,994), Ensembl Gene IDs (20,507), and gene descriptions for the Entrez gene IDs obtained from TxDb.Hsapiens.UCSC.hg19.knownGene database.

#### Using GTEx eQTL data

Using SNP-gene associations based on GTEx v8 eQTL data (accessed on 20.10.2020)^28^, we associated SNPs with the genes they could potentially regulate. We first mapped the hg38 variants in GTEx v8 to hg19 using ‘rtracklayer’, ‘liftOver’, and ‘hg38ToHg19.over.chain’ packages in R. We generated a combined tissue list, which associates SNP to the gene if there is at least one tissue in which there is a significant (p <= 5e-8) association. We chose to create a combined tissue list due to following reasons: 1) disease-tissue matching is not straightforward, especially for complex system-wide diseases^29^ and 2) the power to detect eQTLs depend heavily on the number of samples from each tissue. Thus relying on incomparable set of eQTL sets from different tissues may bias our analysis.

As a result, there were 3,261,028 unique SNPs associated with 20,043 Ensembl Gene IDs. We used the biomaRt^27^ package in R to retrieve HGNC Symbols (16,068), Entrez IDs (14,574), and gene descriptions.

#### Comparison of proximity and eQTL based mapping

Instead of only focusing on disease-associated SNPs, we first mapped all SNPs that we analyzed to discover if there is a bias for certain genes (e.g. some genes could have many more SNPs because they are longer, or because they are already associated with certain traits and the chip is designed in that way). There were as much as 19,195 SNPs mapped to one gene (CSMD1) by proximity, whereas there were 68 SNPs per gene on average (median). The number of SNPs per gene was on average, higher for the mappings by eQTL (Figure S46a). The maximum was 10720 SNPs for C4A gene and the median number of SNPs per gene was 324. However, we did not consider MHC region in our downstream analysis and thus this region is also excluded. The correlation between the number of SNPs per gene was low (rho = 0.18, Figure S46b). Since the proximity-based mapping is by definition dependent on the gene length, we also tested if there is a significant correlation between the number of SNPs per gene and gene length. While the correlation is low for gene mappings by eQTL (Spearman’s correlation rho = 0.08, p = 2.2e-16), mappings by proximity show a high correlation as expected (Spearman’s correlation rho = 0.87, p < 2.2e-16). This also explains the low correlation between eQTL and proximity-based mappings. We next checked the correlation between the number of SNPs per gene mapped by proximity but only to promoter region. The correlation between the number of SNPs and gene length decreased (rho = 0.21), and the correlation with the number of SNPs by eQTL slightly increased but was still low (rho = 0.14). Overall, we concluded that both eQTL data and proximity-based mapping could capture different information and decided to use both for the downstream analyses.

### GWAS Catalog analysis

We accessed the GWAS Catalog on 30-07-2019 and used v1.0.2 e96 dataset^30^. We excluded all studies which used UK Biobank dataset (n = 190, data courtesy of GWAS Catalog team). Using the associations with a p-value lower than 5×10^−8^, we compiled significant associations between MAPPED_GENEs and MAPPED_TRAITs. We use GWAS catalog analysis to check if our GWAS hits are supported by previous studies and applied a Fisher test between all traits in GWAS catalog and the diseases in our study. P-values are corrected for multiple testing using FDR correction.

### Analysis of the association with aging

We downloaded GenAge human, GenAge model organism^31^ and DrugAge^32^ data on Aug 13, 2019 and CellAge^33^ data on Oct 02, 2019 (CellAge data is kindly provided by Avelar et al.). We used HGNC Symbols for GenAge and CellAge genes. In order to compile genes that are targeted by the drugs in DrugAge database, using the drug names in DrugAge data, we first compiled PubChem IDs using PubChem REST API^34^. Using UniChem^35^, we mapped PubChem IDs to ChEMBL IDs^36^. Next, using DGIdb^37^, we compiled the genes targeted by these ChEMBL IDs. As a result, we had 307 genes from GenAge human database, 902 genes from GenAge model organism database, 279 genes from CellAge database, and 714 genes targeted by DrugAge drugs. We next calculated the overlaps between these databases and the genes associated with multiple diseases or multiple categories in different age-of-onset clusters. To calculate the expected values and statistical significance, we used 10,000 permutations calculating the overlap for the same number of random genes among genes that can be detected by GWAS. Then, an odds ratio is calculated by dividing the observed value to the mean of expected values.

### Functional Enrichment Test

Using the goseq package in R^38^, which takes the gene length bias into account, we performed a functional analysis of the genes associated with different age-of-onset clusters. Using GO categories with more than 10 and less than 500 annotated genes, we applied an enrichment test for the Gene Ontology (GO)^39,40^ Biological Process (BP), Molecular Function (MF), and Cellular Compartment (CC) categories. BY correction^41^ is applied to the p-values for all tests for all clusters and 3 GO Categories (BP, MF, and CC) combined. We considered associations with a BY-corrected p-value lower than 0.05 as significant. For the ease of visualization and comprehension we selected representative categories for significant associations as follows: For each cluster and GO Ontology (*i*.*e*. BP, MF, CC) separately; i) Jaccard similarity index (*i*.*e*. number of genes in common divided by the number of unique genes in each category combined) is calculated between all significantly associated GO Categories; ii) Jaccard indices are hierarchically clustered and cut to k number of groups, where k is the minimum number of clusters which ensures median Jaccard similarity within a cluster is above 0.5; iii) The category with the highest average similarity to other categories in the same cluster is assigned as the representative.

### Gene Expression Analysis

Using GTEx v8 data^42^, we analyzed age-related changes in gene expression for the tissues with at least 10 samples (n = 48). We filtered out the genes with median TPM lower than 1. We next log2 transformed expression matrix and used linear model to correct for the effect of sex and death. The normalized expression matrix is quantile normalized using normalize.quantiles function from preprocessCore package^43^ in R. We excluded any sample that deviates 3 standard deviation from the first 4 principal components calculated for this quantile normalized matrix using ‘prcomp’ function with scale = T argument and repeated the procedure until there is no outlier in the data (32 tissues did not have any outliers, 14 had all outliers removed in one round, 1 tissue required 2 rounds and 1 tissue - Cells-Cultured fibroblasts – required 5 rounds of outlier removal). The whole analysis pipeline and quality control steps and results are available in https://github.com/mdonertas/aging_in_GTEx_v8. In order to match age-related expression changes with the variants, we used tissue-specific eQTL data. More specifically, we identified the gene-tissue pairs that have significant eQTL associations and matched their gene expression levels. We only considered genes with eQTLs in the same direction. If disease associated variants are associated with different direction of change for a gene, we excluded these cases from the analysis given in Figures S31-32.

### Methylation Analysis

Using the reported gene sets as differentially methylated during ageing from two studies^44,45^, we checked the overlap with our gene sets. We repeated the same analysis applied to calculate the overlap with aging-related genes.

### Drug Repurposing

We searched for the drugs that specifically target multicategory genes in cluster 1, cluster 2, or cluster 1 and 2. Using the Fisher’s exact test, we compiled the drugs in DGIdb^37^ that specifically target these genes (p≤0.01) and drugs that target only one gene in one of these clusters. Importantly, we excluded all non-specific drugs (*i*.*e*. targeting more than 10 genes) from the analyses. The interaction data is compiled from DGIdb, and the names, indications and phases of the drugs are obtained from ChEMBL REST API^36^.

### Evolutionary Analysis

In order to test the mutation accumulation and antagonistic pleiotropy theories of aging we used the risk allele frequencies in UK Biobank and 1000 Genomes super-populations^46^. A risk allele is an allele that shows positive association with a disease. Since the SNPs are not independent and have similar allele frequencies in a given LD block, we analyzed LD blocks instead of individual SNPs and used the median risk allele frequency for a given LD block. We used only the biallelic SNPs for this analysis. Allele frequencies for UK Biobank are calculated using BOLT-LMM and the allele frequencies for 1000 Genome super-populations are obtained from the vcf file provided on the 1000 Genomes project website. To test the antagonistic pleiotropy excess, we calculated the proportion of antagonistic vs. agonist SNPs within the same vs. different age-of-onset clusters using Fisher’s exact test. We considered pleiotropic SNPs as agonist if the risk allele for two or more diseases are the same, and antagonist if the risk alleles are opposite. We only tested the risk allele frequency differences between cluster 1 and cluster 2. Also, we excluded any SNPs that are antagonistic within an age-of-onset cluster and agonist between clusters.

## References

1. López-Otín, C., Blasco, M. A., Partridge, L., Serrano, M. & Kroemer, G. The hallmarks of aging. Cell 153, 1194–1217 (2013).

2. Crimmins, E. M. Lifespan and Healthspan: Past, Present, and Promise. Gerontologist 55, 901–911 (2015).

3. Partridge, L., Deelen, J. & Slagboom, P. E. Facing up to the global challenges of ageing. Nature 561, 45–56 (2018).

4. Niccoli, T. & Partridge, L. Ageing as a risk factor for disease. Curr. Biol. 22, R741–52 (2012).

5. Flatt, T. & Partridge, L. Horizons in the evolution of aging. BMC Biol. 16, 93 (2018).

6. Medvedev, Z. A. An attempt at a rational classification of theories of ageing. Biol. Rev. Camb. Philos. Soc. 65, 375–398 (1990).

7. Medawar, P. B. Unsolved problem of biology. Med. J. Aust. 1, 854–855 (1953).

8. Williams, G. C. Pleiotropy, Natural Selection, and the Evolution of Senescence. Evolution 11, 398–411 (1957).

9. Bulik-Sullivan, B. et al.. An atlas of genetic correlations across human diseases and traits. Nat. Genet. 47, 1236–1241 (2015).

10. Pickrell, J. K. et al.. Detection and interpretation of shared genetic influences on 42 human traits. Nat. Genet. 48, 709–717 (2016).

11. Cross-Disorder Group of the Psychiatric Genomics Consortium et al. Genetic relationship between five psychiatric disorders estimated from genome-wide SNPs. Nat. Genet. 45, 984–994 (2013).

12. Cortes, A., Albers, P. K., Dendrou, C. A., Fugger, L. & McVean, G. Identifying cross-disease components of genetic risk across hospital data in the UK Biobank. Nat. Genet. 52, 126–134 (2020).

13. Solovieff, N., Cotsapas, C., Lee, P. H., Purcell, S. M. & Smoller, J. W. Pleiotropy in complex traits: challenges and strategies. Nat. Rev. Genet. 14, 483–495 (2013).

14. Ellinghaus, D. et al.. Analysis of five chronic inflammatory diseases identifies 27 new associations and highlights disease-specific patterns at shared loci. Nat. Genet. 48, 510–518 (2016).

15. Parkes, M., Cortes, A., van Heel, D. A. & Brown, M. A. Genetic insights into common pathways and complex relationships among immune-mediated diseases. Nat. Rev. Genet. 14, 661–673 (2013).

16. Bien, S. A. & Peters, U. Moving from one to many: insights from the growing list of pleiotropic cancer risk genes. Br. J. Cancer 120, 1087–1089 (2019).

17. Johnson, S. C., Dong, X., Vijg, J. & Suh, Y. Genetic evidence for common pathways in human age-related diseases. Aging Cell 14, 809–817 (2015).

18. Fernandes, M. et al.. Systematic analysis of the gerontome reveals links between aging and age-related diseases. Hum. Mol. Genet. 25, 4804–4818 (2016).

19. Wang, J., Zhang, S., Wang, Y., Chen, L. & Zhang, X.-S. Disease-aging network reveals significant roles of aging genes in connecting genetic diseases. PLoS Comput. Biol. 5, e1000521 (2009).

20. Bycroft, C. et al.. The UK Biobank resource with deep phenotyping and genomic data. Nature 562, 203–209 (2018).

21. UKB : Data-Field 41262. https://biobank.ndph.ox.ac.uk/showcase/field.cgi?id=41262.

22. Chouakria, A. D. & Nagabhushan, P. N. Adaptive dissimilarity index for measuring time series proximity. Adv. Data Anal. Classif. 1, 5–21 (2007).

23. Loh, P.-R. et al.. Efficient Bayesian mixed-model analysis increases association power in large cohorts. Nat. Genet. 47, 284–290 (2015).

24. Anderson, C. A. et al.. Data quality control in genetic case-control association studies. Nat. Protoc. 5, 1564–1573 (2010).

25. Zhou, W. et al.. Efficiently controlling for case-control imbalance and sample relatedness in large-scale genetic association studies. Nat. Genet. 50, 1335–1341 (2018).

26. Pe’er, I., Yelensky, R., Altshuler, D. & Daly, M. Estimation of the Multiple Testing Burden for Genomewide Association Studies of Common Variants. Nature Precedings (2007) doi:10.1038/npre.2007.359.1.

27. Panagiotou, O. A., Ioannidis, J. P. A. & Genome-Wide Significance Project. What should the genome-wide significance threshold be? Empirical replication of borderline genetic associations. Int. J. Epidemiol. 41, 273–286 (2012).

28. MHC region of the human genome - Genome Reference Consortium. https://www.ncbi.nlm.nih.gov/grc/human/regions/MHC?asm=GRCh37.

29. Bulik-Sullivan, B. K. et al.. LD Score regression distinguishes confounding from polygenicity in genome-wide association studies. Nat. Genet. 47, 291–295 (2015).

30. Berisa, T. & Pickrell, J. K. Approximately independent linkage disequilibrium blocks in human populations. Bioinformatics 32, 283–285 (2016).

31. Ning, Z., Pawitan, Y. & Shen, X. High-definition likelihood inference of genetic correlations across human complex traits. Nat. Genet. (2020) doi:10.1038/s41588-020-0653-y.

32. UK Biobank — Neale lab. http://www.nealelab.is/uk-biobank/.

33. O’Connor, L. J. & Price, A. L. Distinguishing genetic correlation from causation across 52 diseases and complex traits. Nat. Genet. 50, 1728–1734 (2018).

34. Gamazon, E. R. et al.. Using an atlas of gene regulation across 44 human tissues to inform complex disease- and trait-associated variation. Nat. Genet. 50, 956–967 (2018).

35. Tacutu, R. et al.. Human Ageing Genomic Resources: new and updated databases. Nucleic Acids Res. 46, D1083–D1090 (2018).

36. Barardo, D. et al.. The DrugAge database of aging-related drugs. Aging Cell vol. 16 594–597 (2017).

37. Avelar, R. A. et al.. A multidimensional systems biology analysis of cellular senescence in aging and disease. Genome Biol. 21, 91 (2020).

38. Adelman, E. R. et al.. Aging Human Hematopoietic Stem Cells Manifest Profound Epigenetic Reprogramming of Enhancers That May Predispose to Leukemia. Cancer Discov. 9, 1080–1101 (2019).

39. Marttila, S. et al.. Ageing-associated changes in the human DNA methylome: genomic locations and effects on gene expression. BMC Genomics 16, 179 (2015).

40. Sun, L.-Q. et al.. Growth retardation and premature aging phenotypes in mice with disruption of the SNF2-like gene, PASG. Genes Dev. 18, 1035–1046 (2004).

41. Rodríguez, J. A. et al.. Antagonistic pleiotropy and mutation accumulation influence human senescence and disease. Nat Ecol Evol 1, 55 (2017).

42. 1000 Genomes Project Consortium et al. A global reference for human genetic variation. Nature 526, 68–74 (2015).

43. Bitarello, B. D. et al.. Signatures of Long-Term Balancing Selection in Human Genomes. Genome Biol. Evol. 10, 939–955 (2018).

44. Kosiol, C. et al.. Patterns of positive selection in six Mammalian genomes. PLoS Genet. 4, e1000144 (2008).

45. Nielsen, R. et al.. A scan for positively selected genes in the genomes of humans and chimpanzees. PLoS Biol. 3, e170 (2005).

46. Shultz, A. J. & Sackton, T. B. Immune genes are hotspots of shared positive selection across birds and mammals. Elife 8, (2019).

47. Fisher, R. A. The genetical theory of natural selection. 272, (1930).

48. Wolfson, M., Budovsky, A., Tacutu, R. & Fraifeld, V. The signaling hubs at the crossroad of longevity and age-related disease networks. Int. J. Biochem. Cell Biol. 41, 516–520 (2009).

## References

1. Anderson, C. A. et al.. Data quality control in genetic case-control association studies. Nat. Protoc. 5, 1564–1573 (2010).

2. Bycroft, C. et al.. The UK Biobank resource with deep phenotyping and genomic data. Nature 562, 203–209 (2018).

3. Zhou, W. et al.. Efficiently controlling for case-control imbalance and sample relatedness in large-scale genetic association studies. Nat. Genet. 50, 1335–1341 (2018).

4. Kanchi, K. L. et al.. Integrated analysis of germline and somatic variants in ovarian cancer. Nat. Commun. 5, 3156 (2014).

5. Khurana, E. et al.. Role of non-coding sequence variants in cancer. Nat. Rev. Genet. 17, 93–108 (2016).

6. Poduri, A., Evrony, G. D., Cai, X. & Walsh, C. A. Somatic mutation, genomic variation, and neurological disease. Science 341, 1237758 (2013).

7. Zhang, L. & Vijg, J. Somatic Mutagenesis in Mammals and Its Implications for Human Disease and Aging. Annu. Rev. Genet. 52, 397–419 (2018).

8. Finkel, T., Serrano, M. & Blasco, M. A. The common biology of cancer and ageing. Nature 448, 767–774 (2007).

9. Sanchez-Valle, J. et al.. Unveiling the molecular basis of disease co-occurrence: towards personalized comorbidity profiles. bioRxiv 431312 (2018) doi:10.1101/431312.

10. Gutiérrez-Sacristán, A. et al.. comoRbidity: an R package for the systematic analysis of disease comorbidities. Bioinformatics 34, 3228–3230 (2018).

11. Chouakria, A. D. & Nagabhushan, P. N. Adaptive dissimilarity index for measuring time series proximity. Adv. Data Anal. Classif. 1, 5–21 (2007).

12. Montero, P. & Vilar, J. TSclust: An R Package for Time Series Clustering. Journal of Statistical Software, Articles 62, 1–43 (2014).

13. Berndt, D. J. & Clifford, J. Using Dynamic Time Warping to Find Patterns in Time Series. in Proceedings of the 3rd International Conference on Knowledge Discovery and Data Mining 359–370 (AAAI Press, 1994).

14. Partitioning Around Medoids (Program PAM). in Finding Groups in Data (eds. Kaufman, L. & Rousseeuw, P. J.) 68–125 (John Wiley & Sons, Inc., 1990).

15. Maechler, M., Rousseeuw, P., Struyf, A., Hubert, M. & Hornik, K. cluster: Cluster Analysis Basics and Extensions. (2019).

16. Tibshirani, R., Walther, G. & Hastie, T. Estimating the number of clusters in a data set via the gap statistic. Journal of the Royal Statistical Society: Series B (Statistical Methodology) vol. 63 411–423 (2001).

17. Loh, P.-R. et al.. Efficient Bayesian mixed-model analysis increases association power in large cohorts. Nat. Genet. 47, 284–290 (2015).

18. Loh, P.-R. BOLT-LMM v2. 3.1 User Manual. https://data.broadinstitute.org/alkesgroup/BOLT-LMM/ (2017).

19. Stephenson, J. D., Laskowski, R. A., Nightingale, A., Hurles, M. E. & Thornton, J. M. VarMap: a web tool for mapping genomic coordinates to protein sequence and structure and retrieving protein structural annotations. Bioinformatics (2019) doi:10.1093/bioinformatics/btz482.

20. Berisa, T. & Pickrell, J. K. Approximately independent linkage disequilibrium blocks in human populations. Bioinformatics 32, 283–285 (2016).

21. Ning, Z., Pawitan, Y. & Shen, X. High-definition likelihood inference of genetic correlations across human complex traits. Nat. Genet. (2020) doi:10.1038/s41588-020-0653-y.

22. UK Biobank — Neale lab. http://www.nealelab.is/uk-biobank/.

23. O’Connor, L. J. & Price, A. L. Distinguishing genetic correlation from causation across 52 diseases and complex traits. Nat. Genet. 50, 1728–1734 (2018).

24. Obenchain, V. et al.. VariantAnnotation: a Bioconductor package for exploration and annotation of genetic variants. Bioinformatics 30, 2076–2078 (2014).

25. Carlson, M. & Maintainer, B. P. TxDb.Hsapiens.UCSC.hg19.knownGene: Annotation package for TxDb object(s). (2015).

26. Lawrence, M. et al.. Software for computing and annotating genomic ranges. PLoS Comput. Biol. 9, e1003118 (2013).

27. Durinck, S., Spellman, P. T., Birney, E. & Huber, W. Mapping identifiers for the integration of genomic datasets with the R/Bioconductor package biomaRt. Nat. Protoc. 4, 1184–1191 (2009).

28. Gamazon, E. R. et al.. Using an atlas of gene regulation across 44 human tissues to inform complex disease- and trait-associated variation. Nat. Genet. 50, 956–967 (2018).

29. Lage, K. et al.. A large-scale analysis of tissue-specific pathology and gene expression of human disease genes and complexes. Proc. Natl. Acad. Sci. U. S. A. 105, 20870–20875 (2008).

30. Buniello, A. et al.. The NHGRI-EBI GWAS Catalog of published genome-wide association studies, targeted arrays and summary statistics 2019. Nucleic Acids Res. 47, D1005–D1012 (2019).

31. Tacutu, R. et al.. Human Ageing Genomic Resources: new and updated databases. Nucleic Acids Res. 46, D1083–D1090 (2018).

32. Barardo, D. et al.. The DrugAge database of aging-related drugs. Aging Cell 16, 594–597 (2017).

33. Avelar, R. A. et al.. A multidimensional systems biology analysis of cellular senescence in aging and disease. Genome Biol. 21, 91 (2020).

34. Kim, S. et al.. PubChem 2019 update: improved access to chemical data. Nucleic Acids Res. 47, D1102–D1109 (2019).

35. Chambers, J. et al.. UniChem: a unified chemical structure cross-referencing and identifier tracking system. J. Cheminform. 5, 3 (2013).

36. Gaulton, A. et al.. The ChEMBL database in 2017. Nucleic Acids Res. 45, D945–D954 (2017).

37. Cotto, K. C. et al.. DGIdb 3.0: a redesign and expansion of the drug–gene interaction database. Nucleic Acids Res. 46, D1068–D1073 (2018).

38. Young, M. D., Wakefield, M. J., Smyth, G. K. & Oshlack, A. Gene ontology analysis for RNA-seq: accounting for selection bias. Genome Biol. 11, R14 (2010).

39. The Gene Ontology Consortium & The Gene Ontology Consortium. The Gene Ontology Resource: 20 years and still GOing strong. Nucleic Acids Research vol. 47 D330–D338 (2019).

40. Ashburner, M. et al.. Gene ontology: tool for the unification of biology. The Gene Ontology Consortium. Nat. Genet. 25, 25–29 (2000).

41. Benjamini, Y. & Yekutieli, D. The control of the false discovery rate in multiple testing under dependency. Ann. Stat. 29, 1165–1188 (2001).

42. GTEx Consortium. Human genomics. The Genotype-Tissue Expression (GTEx) pilot analysis: multitissue gene regulation in humans. Science 348, 648–660 (2015).

43. Bolstad, B. preprocessCore: A collection of pre-processing functions. (2020).

44. Adelman, E. R. et al.. Aging Human Hematopoietic Stem Cells Manifest Profound Epigenetic Reprogramming of Enhancers That May Predispose to Leukemia. Cancer Discov. 9, 1080–1101 (2019).

45. Marttila, S. et al.. Ageing-associated changes in the human DNA methylome: genomic locations and effects on gene expression. BMC Genomics 16, 179 (2015).

46. 1000 Genomes Project Consortium et al. A global reference for human genetic variation. Nature 526, 68–74 (2015).

