## Supplemental Information and Figures for "Common genetic associations between age-related diseases"

### List of Supplementary Tables

**Table S1:** List of diseases in each age-of-onset cluster.

**Table S2:** Partially genetic causal relationship between diseases for all significant pairs (FDR corrected p≤0.01 and mean GCP>0.6).

**Table S3:** Enrichment results comparing GWAS hits for the UKBB diseases with the GWAS-Catalog traits.

**Table S4:** List of multidisease and multicategory genes associated with each cluster or cluster combinations.

**Table S5:** List of genes overlapping between aging databases and the multidisease or multicategory genes associated with each cluster.

**Table S6:** GO enrichment results for the multicategory cluster 1 genes that show increased gene expression at the age of 40, and multicategory cluster 2 and 3 genes that show decreased gene expression at the age of 40.

**Table S7:** GO enrichment results.

**Table S8:** Summary of Fisher's exact test result, testing the agonist to antagonist ratio within vs. across clusters.

**Table S9:** List of antagonistic associations between cluster 1 and cluster 2 diseases and the corresponding genes. 'Median RAF Difference' corresponds to the median value of the risk allele frequencies (cluster1 minus cluster2) for each antagonistic SNP.

**Table S10:** List of drugs specifically targeting multicategory clusters 1, 2, or '1 & 2' genes.

### Supplementary Information

#### UK Biobank Data

Using the samples that passed quality control (see Methods, n = 484,598), we first did an exploratory analysis using the basic demographics, disease data, and aging-related data fields.

There were more females (n=262,758) than males (n = 221,840) (Figure S2a). The age range of participants during the first visit was between 37 (minimum age of males = 37, females = 39) to 73 (maximum age of males = 73, females = 71) with a median value of 58 (median age of males = 58, females = 57) (Figure S2b). There were 13,697 participants who died after participating in the study and the death rate was higher in males (Figure S2c). As expected, height, weight, and BMI also differed in females and males (Figure S2d-f).

Participants were also asked how they rate their health, how satisfied they are with their health, smoking status, alcohol drinker status, if other people generally say they look i) younger than they are, ii) about their age, or iii) older than they are, and if they had a close relative who had non-accidental sudden death. Overall, more people rated their health high and were happy with their health (Figure S3a-b). Most of the UKBB participants either never smoked or were previous smokers (Figure S3c) and are current alcohol drinkers (Figure S3d). Most of the participants also reported that people generally think they are either younger than their age or about the same age (Figure S3e). Most of the participants did not have any close relatives who died suddenly from non-accidental causes (Figure S3f).

We also checked the distribution of other aging-related fields, namely parents' age at death, age at menarche, and age at menopause. There were 391,842 participants with at least one parent idead. The distribution was wide (10 to 117), but the majority of the data (between the first and third quantiles) lie between 65 and 79.5 (average age at death) (Figure S4a). The age at menarche differed between 5 and 25, with a median of 13 (Figure S4b). The age at menopause differed between 18 and 68, with a median of 50 (Figure S4c).

The number of self-reported operations ranged between 1 to 32, with a median of 1 and the number of self-reported medications ranged between 0 to 48, with a median of 2 (Figure S5a). Among 39,910 participants with cancer, most of them had only one cancer, while there was also a participant with 6 cancers (Figure S5b).

We then checked the correlations between these traits (Figure S6). Age when attended assessment center was very strongly correlated with age at death. It also showed a correlation with parental age at death, the number of non-cancer diseases, and the number of medications taken. "Overall health rating" and "health satisfaction" were also correlated with the number of diseases and medications. Moreover, these values also showed a correlation with BMI and weight. While 'sex' and 'standing height' were correlated with 'weight', they were not correlated with 'BMI' and 'overall health rating' which are both correlated with 'weight'. BMI was also correlated with 'number of medications taken'.

Although we did not use these traits directly in our analysis, we performed an exploratory analysis to decide on the potential covariates to use in the GWAS model (see Methods).

**Disease Categories**

The UKBB includes disease information from two sources: i) disease ICD10/9 codes based on **h**ospital **e**pisode **s**tatistics (HES) and ii) the **s**elf-**r**eported (SR) diseases. In our study, we use SR diseases as these data provide the age at diagnosis. Since the first age at diagnosis information for ICD10 codes starts only in 1992 (http://biobank.ndph.ox.ac.uk/showcase/field.cgi?id=41262) self-reported data is less biased by the age distribution of UK Biobank participants.. Moreover, a previous study using the UKBB suggested that GWAS using self-reported diseases and ICD-10 codes were sufficiently similar[1].

Like ICD10 codes, the UKBB SR Diseases are defined in a hierarchical structure (Figure S7). This tree is constructed by the UKBB nurses and it mostly reflects the system or the tissue in which that disease is most symptomatic in. Participants enter SR disease data with a trained nurse, who guides them. However, we some participants did not consider the disease hierarchy while some did. For example, some patients having 'essential hypertension' also reported having 'hypertension' which is the parent node, while some did not. In order not to bias data, we propagated disease data towards upper levels, so that a participant with a disease at a lower level is always annotated with the connected nodes at upper levels.

Importantly, we only considered 116 non-cancer diseases with at least 2,000 cases and that were not sex-specific. Although we exclude sex-limited diseases, we included the ones that were more prevalent in females (thyroid problem, hypothyroidism, bone disorder, osteoporosis) or in males (abdominal hernia, gout, heart attack) (Figure S8).

**Disease Co-occurrences**

We next calculated disease co-occurrences, using relative risk score to calculate associations and ϕ values as a measure of robustness[2–5] (Methods). There were five major clusters with high relative risk scores and robustness and they seemingly cluster by disease categories: i) musculoskeletal/trauma diseases and early-onset gastrointestinal diseases such as appendicitis, ii) other musculoskeletal/trauma diseases such as sciatica and disc problems, iii) respiratory/ENT diseases such as bronchitis and pneumonia, iv) cardiovascular diseases and diabetes, v) retinal problem, glaucoma, and cataract (Figure S9). While most of these clusters are biologically plausible, some could be explained by reporting bias, e.g. it is plausible that only a fraction of people reported their childhood diseases, resulting in an artificial association between bone fractures and appendicitis. Moreover, we saw a strong negative correlation between osteoarthritis and arthritis (nos). The disease 'arthritis (nos)' does not include osteoarthritis by definition (nos = not osteoarthritis) and seeing this association suggests that we can detect co-occurrences reliably.

**Drug repurposing to improve healthspan**

Identification of drugs that can target the multicategory genes associated with diseases in clusters 1 and 2 could enable the treatment of many diseases simultaneously and improve healthspan in the elderly. Thus, we investigated if there are drugs that target these genes specifically ($p \leq 0.01$ or having only one specific target, Figure S41). We found drugs targeting multicategory cluster 1 genes i) *ABCC8* and *KCNJ11*, which code for parts of K-ATP channels, ii) *CCND1*, iii) *CDKN2A*, iv) *RSPO3, v) OPRL1, vi) IRS1* and multicategory cluster 2 genes i) *KLKB1* and ii) *HLA-DQB1*. There were also several drugs targeting multicategory genes associated with both cluster 1 and 2 diseases, such as *PPARG*, *INSR*, *FGFR4*, *MAPKAPK5*, *ALDH2*, *PTPN11, MTAP*. One of the drugs we identified, prunetin (targeting *ALDH2*), was previously shown to increase the lifespan of male *Drosophila melanogaster*[6]. Importantly, the significant hits included approved drugs for 12 conditions, including diabetes, bone diseases, cancer, and thrombophlebitis (list of all drugs and indications available in Table S10). Although the majority of these conditions are age-related, drugs used to treat these conditions do not necessarily target the multicategory genes we identified (Figure S42), and thus, the drugs identified here offer new possibilities to prevent polypharmacy in the elder population if their use is prioritized to treat multiple diseases. Moreover, some of these drugs are already considered for multiple diseases from different categories. For example, acetohexamide, which targets the K-ATP channel, is in use for diabetes mellitus and is undergoing clinical trials for cataracts[7]. Nevertheless, this analyses only offers candidates to be tested for their effect on multiple diseases and future research is necessary to evaluate their effect on extending healthspan and alleviating polypharmacy.


1. Cortes, A., Dendrou, C., Fugger, L. & McVean, G. Systematic classification of shared components of genetic risk for common human diseases. *bioRxiv* (2018).
2. Gutiérrez-Sacristán, A. *et al.* comoRbidity: an R package for the systematic analysis of disease comorbidities. *Bioinformatics* **34**, 3228–3230 (2018).
3. Jiang, Y., Ma, S., Shia, B.-C. & Lee, T.-S. An Epidemiological Human Disease Network Derived from Disease Co-occurrence in Taiwan. *Sci. Rep.* **8**, 4557 (2018).
4. Park, J., Lee, D.-S., Christakis, N. A. & Barabási, A.-L. The impact of cellular networks on disease comorbidity. *Mol. Syst. Biol.* **5**, 262 (2009).
5. Sanchez-Valle, J. *et al.* Unveiling the molecular basis of disease co-occurrence: towards personalized comorbidity profiles. *bioRxiv* 431312 (2018) doi:10.1101/431312.
6. Piegholdt, S., Rimbach, G. & Wagner, A. E. The phytoestrogen prunetin affects body composition and improves fitness and lifespan in male Drosophila melanogaster. *FASEB J.* **30**, 948–958 (2016).
7. Compound: Acetohexamide. https://www.ebi.ac.uk/chembl/compound_report_card/CHEMBL1589/.

**Supplementary Figures**

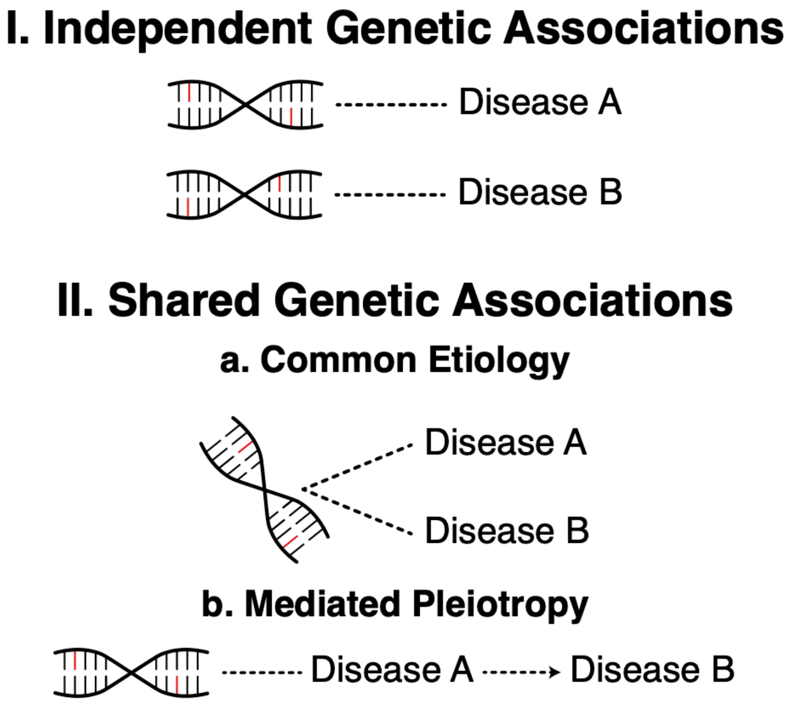


Figure S1: Summary of different models explaining the associations between diseases. Independent genetic associations (I) reflect the case where the number of shared genetic associations between diseases is not more than expected by chance. If the overlap is more than expected (II), it could either reflect (a) common etiology, which reflects shared causes, or (b) mediated pleiotropy, which suggests a common genetic factor influencing only one disease, which in turn increases the risk of a second disease.

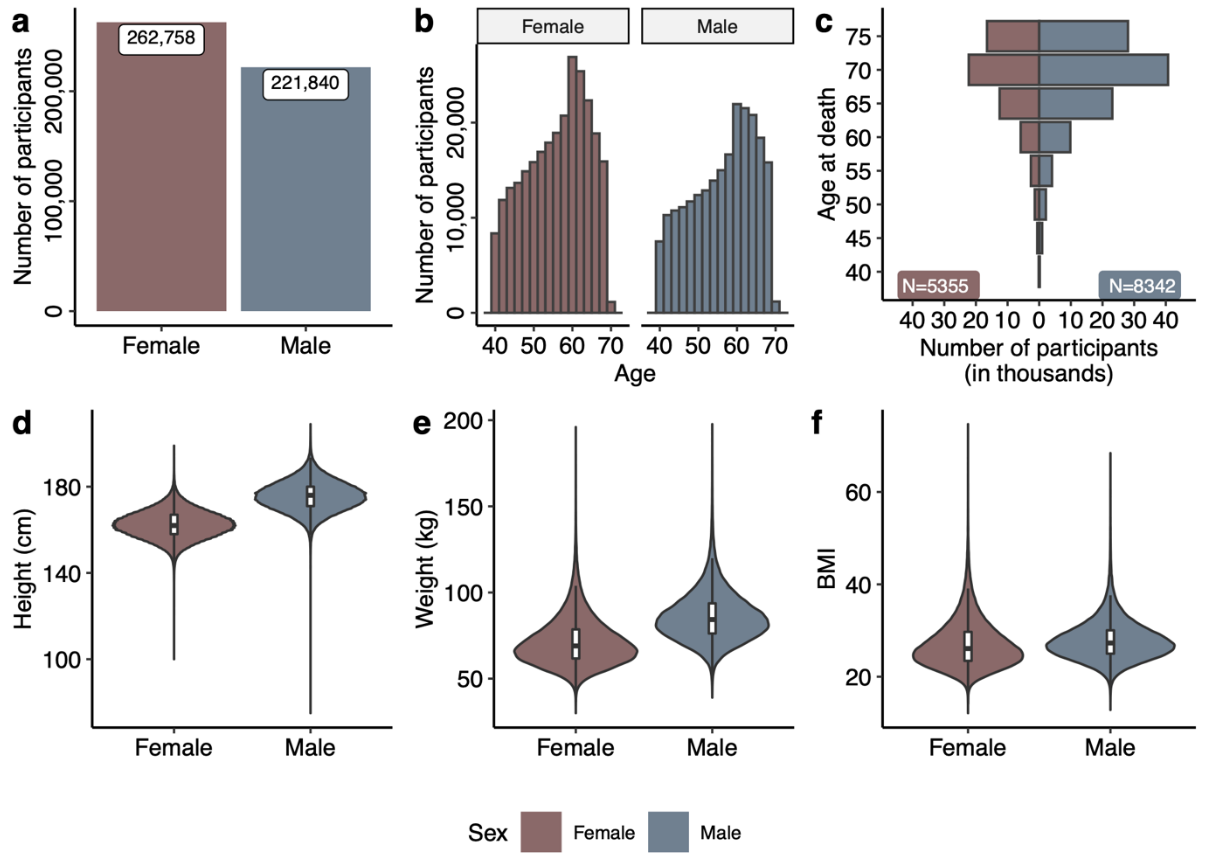


Figure S2: Participant data in UK Biobank after quality control steps. a) The number of female and male participants, b) Age distribution when participants first attended the UKBB assessment center and answered self-reported questions, c) Age at death (every 5 years are binned together) for the participants who died after attending the UKBB assessment center. The values are corrected for the number of female and male participants who passed the ages specified in the y-axis, d) Distributions of 'standing height' field in the UKBB, e) Distributions of 'weight' field in the UKBB, f) Distributions of BMI field calculated using 'standing height' and 'weight' fields in the UKBB (see Methods).

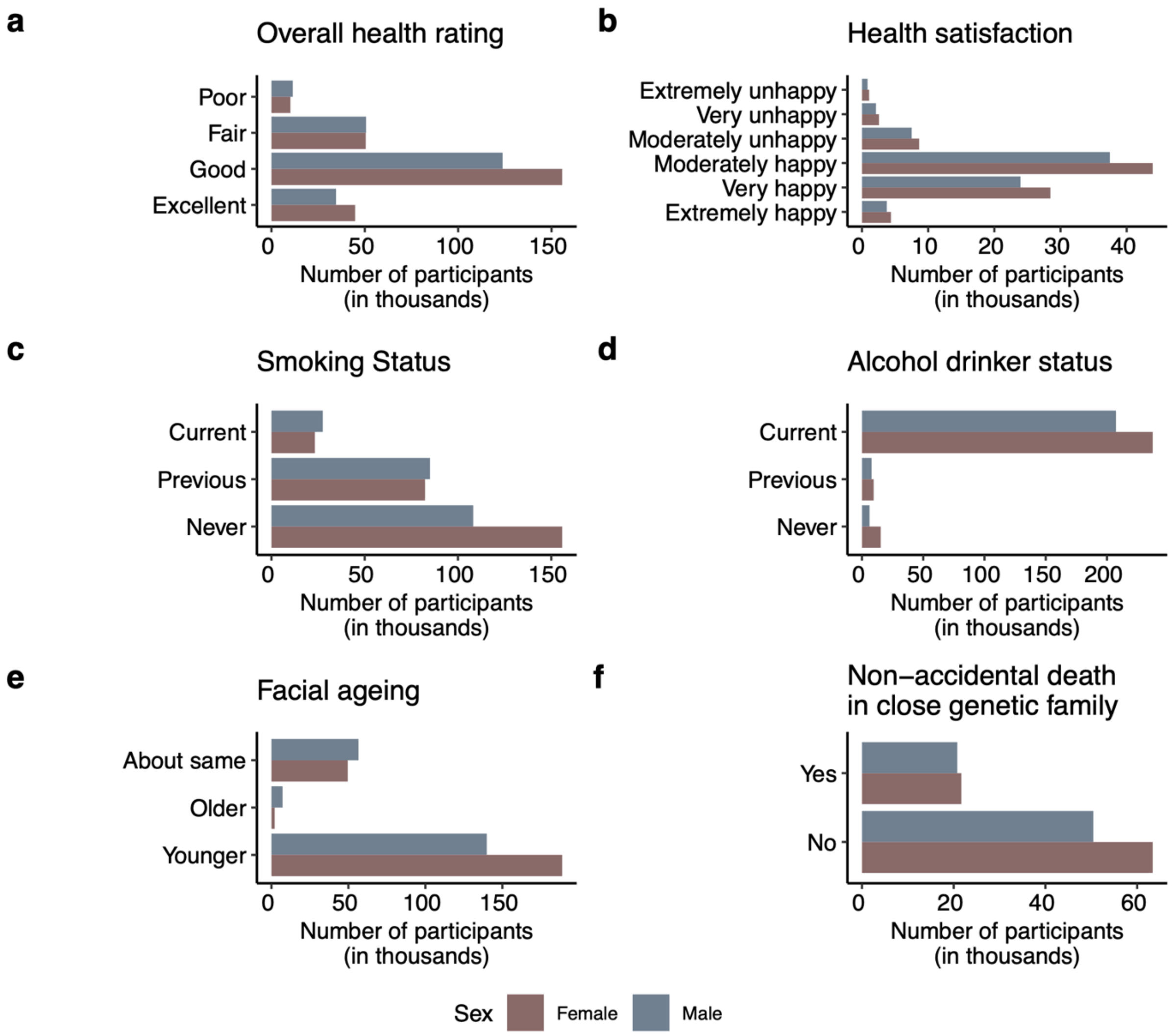


Figure S3: The distribution of a) Overall health rating, b) Health satisfaction, c) Smoking status, d) Alcohol drinker status, e) Facial aging, f) Non-accidental death in close genetic family fields in the UKBB. x-axes show the number of participants, while y-axes are the answers given by the participants.

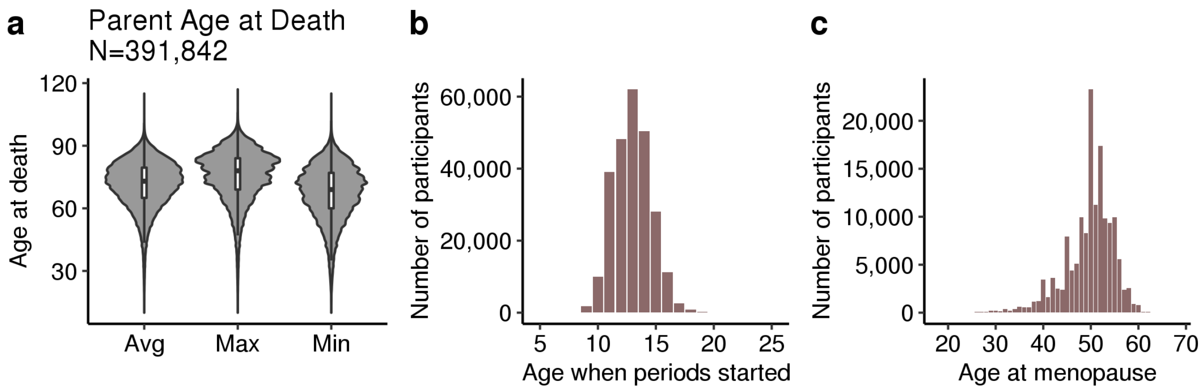


Figure S4: Distributions of a) parents' age at death, b) age when periods started (menarche), and c) Age at menopause (last menstrual period).

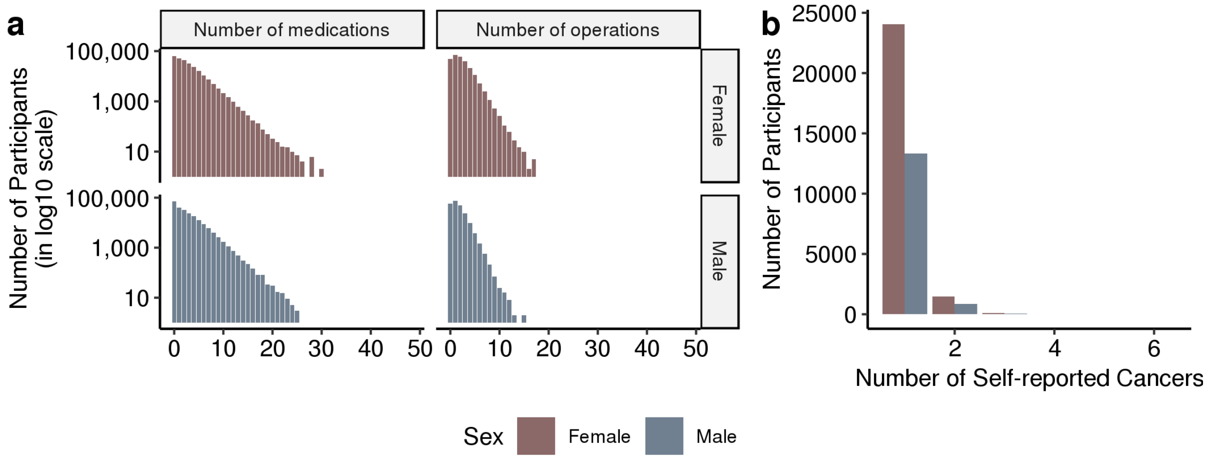


Figure S5: Self-reported health data. a) The number of self-reported medications and operations (x-axes) for the participants in the UK Biobank. The y-axis shows the number of participants on a log10 scale. b) The number of self-reported cancers (x-axis). Y-axis shows the number of participants.

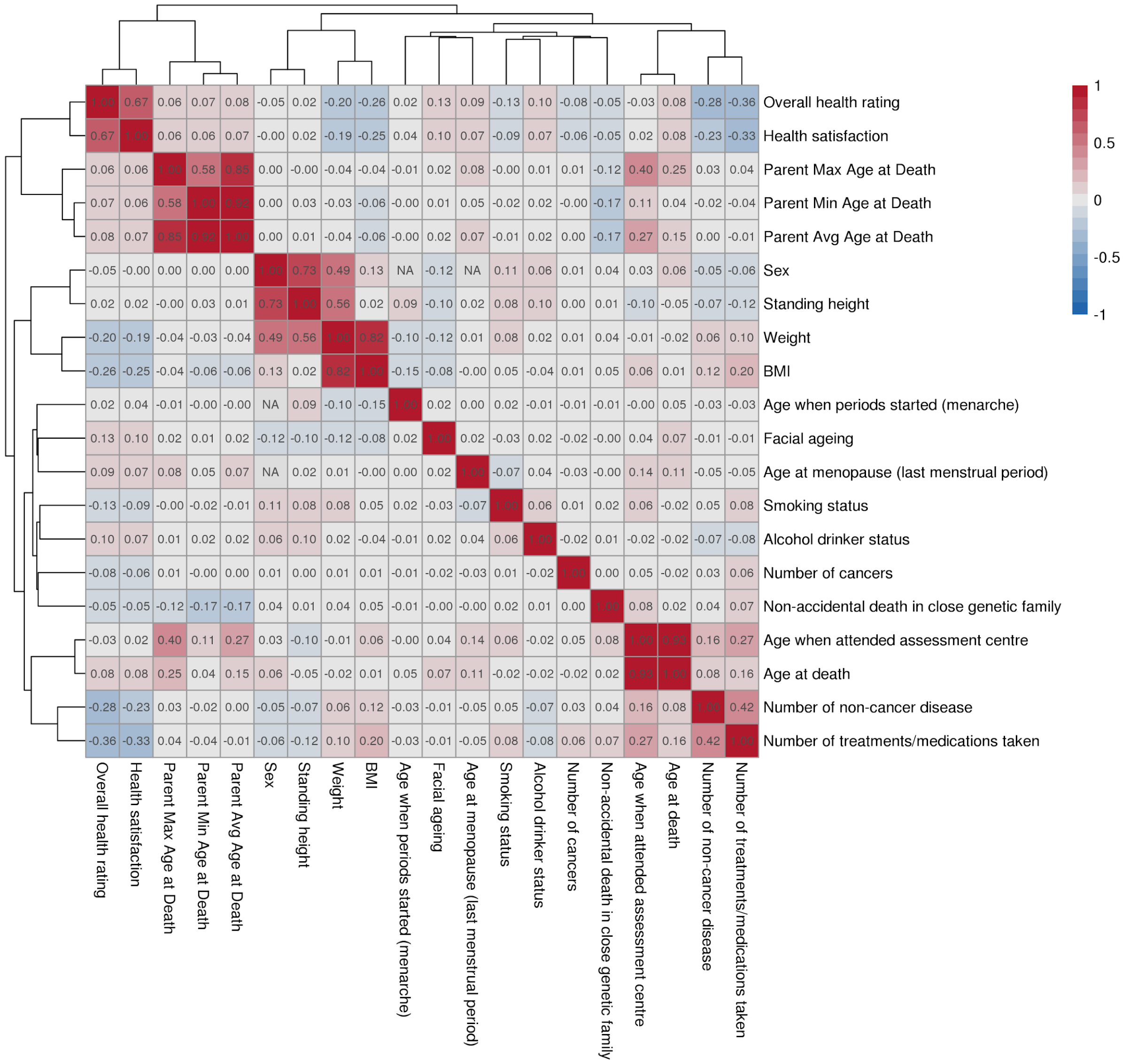


Figure S6: Pairwise correlations between traits. Each row and column shows a trait in the UKBB, and the color shows the pairwise Spearman correlation coefficients between traits. Dark red denotes a strong positive correlation, while dark blue indicates strong negative values. Traits are ordered based on the hierarchical clustering of the correlation coefficients.

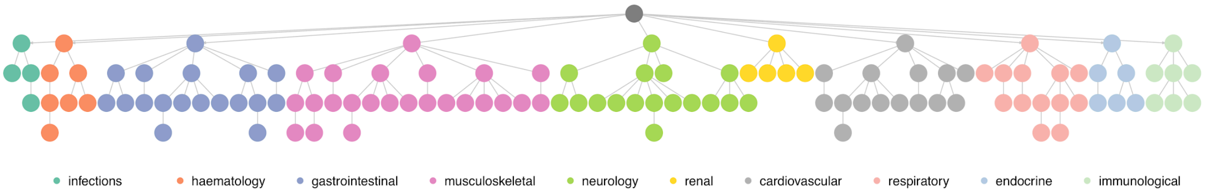


Figure S7: Disease hierarchy for the 116 diseases included in the analysis. The nodes are colored by the disease categories as indicated in the legend.

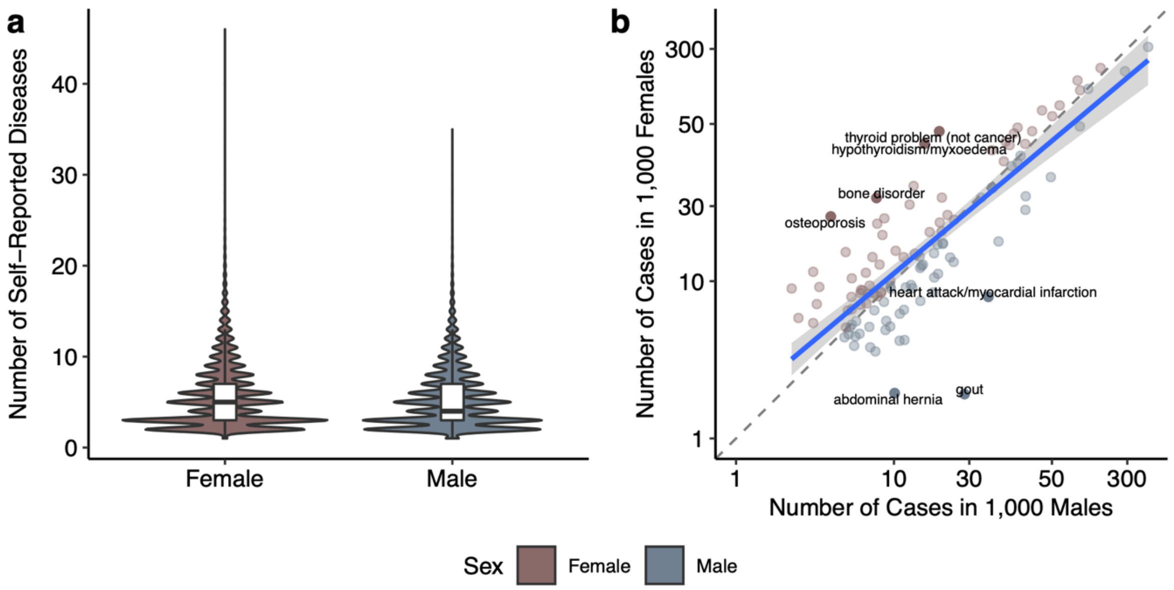


Figure S8: Sex-stratified statistics for 116 selected diseases. a) The distribution of the number of self-reported diseases (y-axis) stratified by sex (x-axis). b) The distribution of disease prevalence in males and females. The x- and y-axes show the number of cases in 1,000 males and females (on a log scale), respectively. The color of each point denotes diseases with a higher prevalence in females (rosy brown, above the dashed line) or males (slate grey, below the dashed line). The linear regression line is depicted as blue. Diseases having a residual value bigger than 3 standard deviations are labeled but not excluded as they are also common in the other sex.

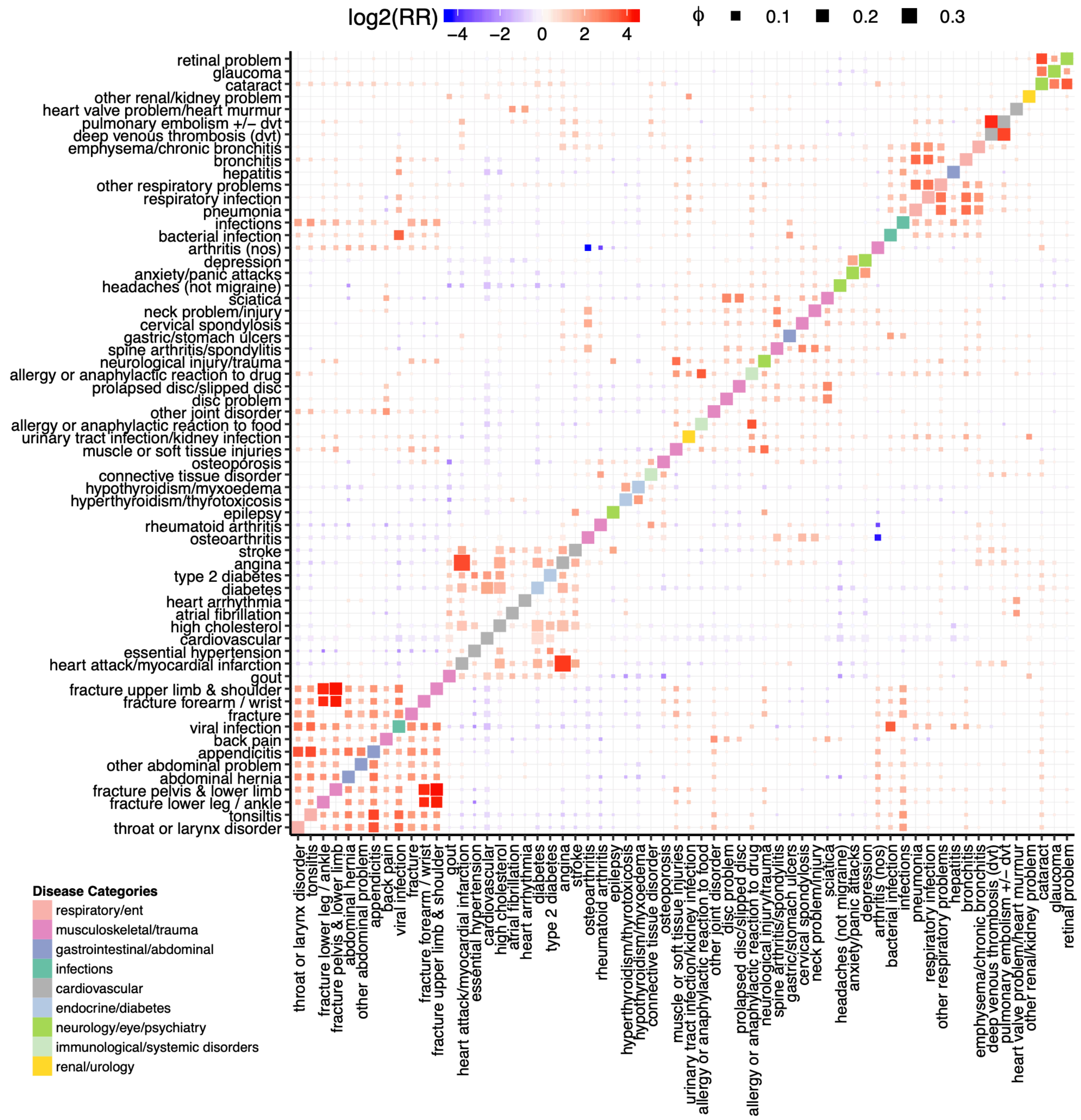


Figure S9: Disease co-occurrence matrix summarizing relative risk scores and correlations. Each row and column denote diseases, ordered by hierarchical clustering of risk scores. The color is defined by relative risk scores while the size is determined by ϕ value, indicating the robustness of the association (see Methods). The diagonal tiles are colored by the UK Biobank's disease hierarchy to visualize if diseases from the same category cluster together. Associations for the 62 diseases that have at least one relative risk ratio higher than four ($log_2RR \geq 2$) or lower than minus four ($log_2RR \leq -2$) are plotted.

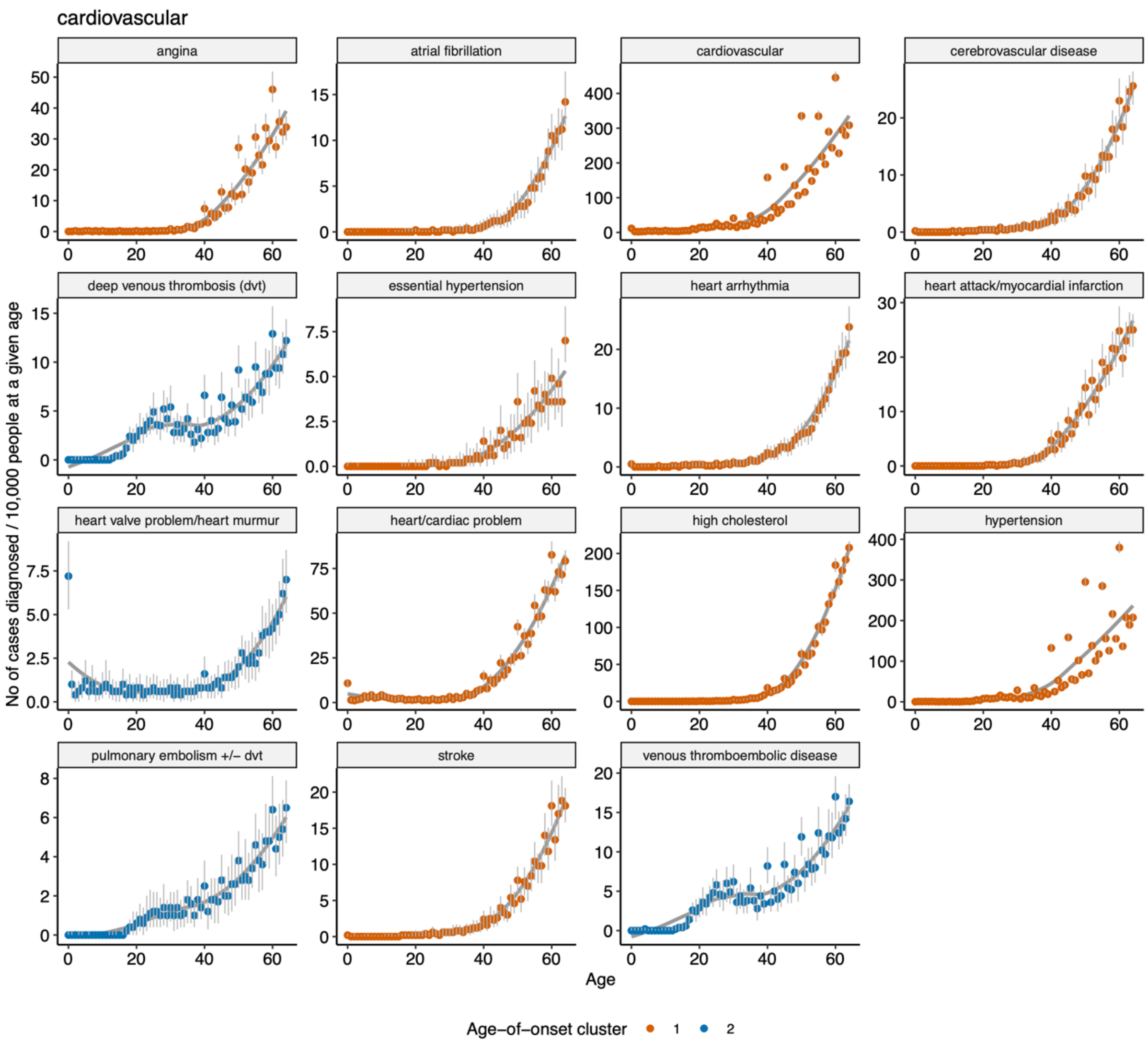


Figure S10: Age-of-onset distributions for the cardiovascular diseases. The y-axis shows how many people in 10,000 are diagnosed with that disease at a certain age (x-axis). The plots are also normalized by the number of people that are older than a given age so that it is unaffected by the distribution of ages in the UKBB (Figure S2b). We ran permutations to define confidence intervals for the disease onset rates. We thus down-sampled the UKBB population using 50,000 participants for 100 times and calculated the median (points, colored by the age-of-onset cluster in Figure 1) and 95% range of all points (gray error lines). A best-fit curve (calculated using loess regression between the medians and age-of-onset) is also displayed.

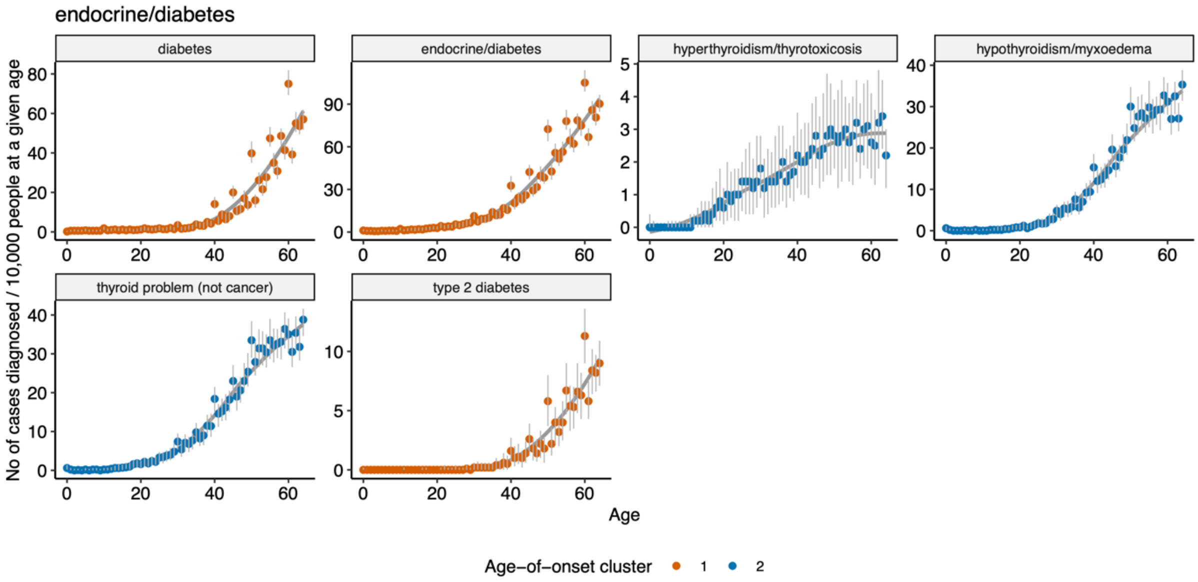


Figure S11: Same as Figure S10, but for endocrine / diabetes diseases.

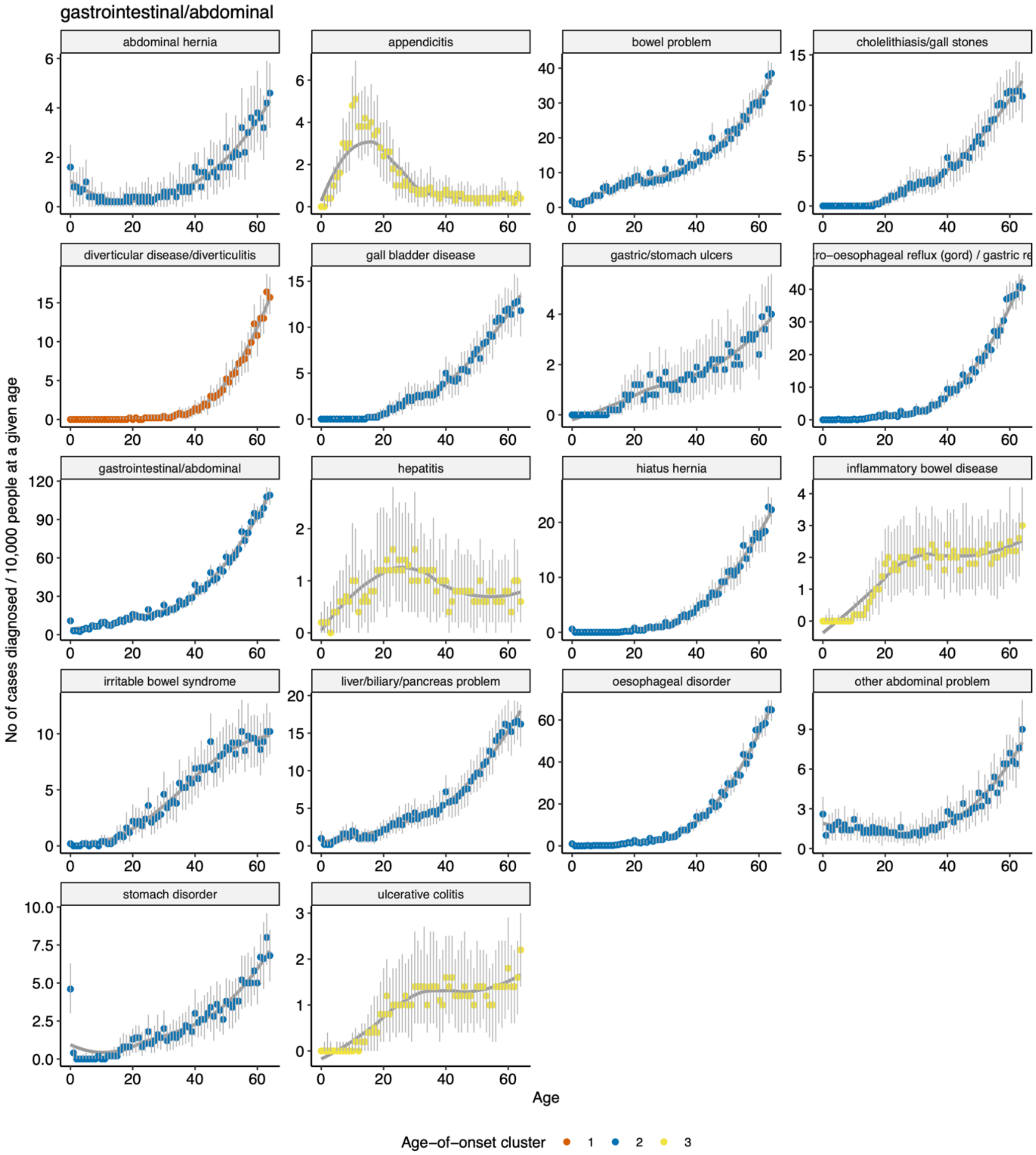


248

249 Figure S12: Same as Figure S10, but for gastrointestinal / abdominal diseases.

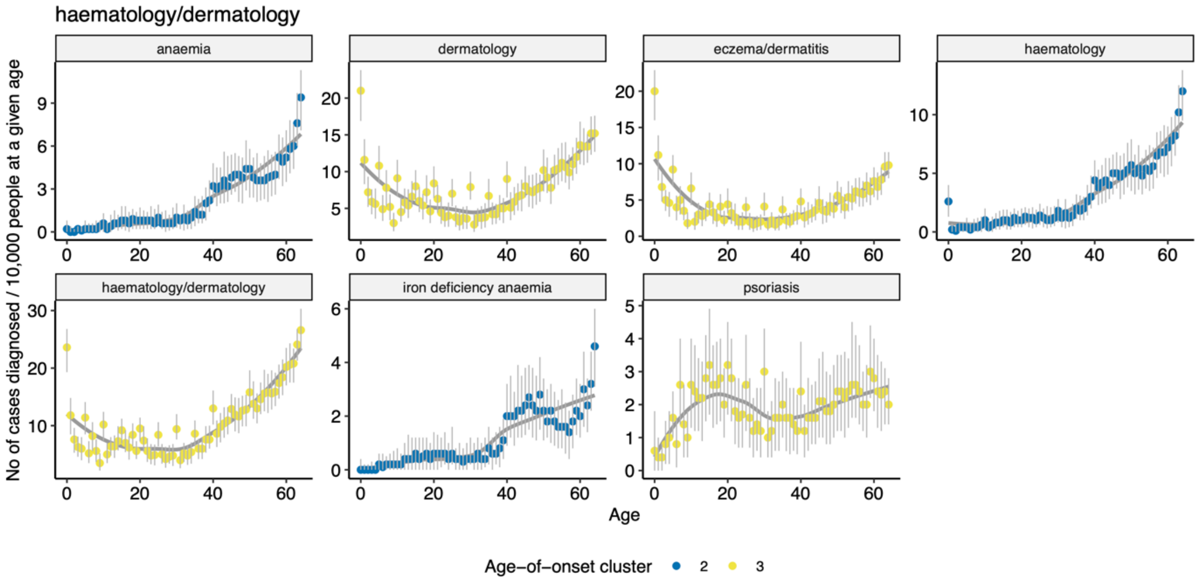


Figure S13: Same as Figure S10, but for haematology / dermatology diseases.

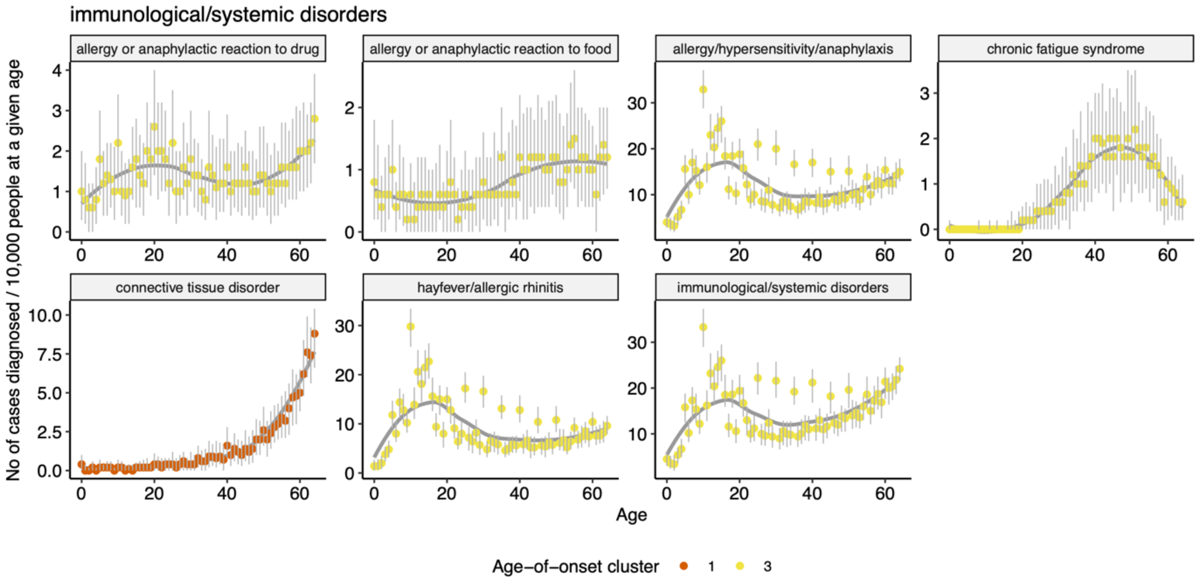


Figure S14: Same as Figure S10, but for immunological / systemic disorders.

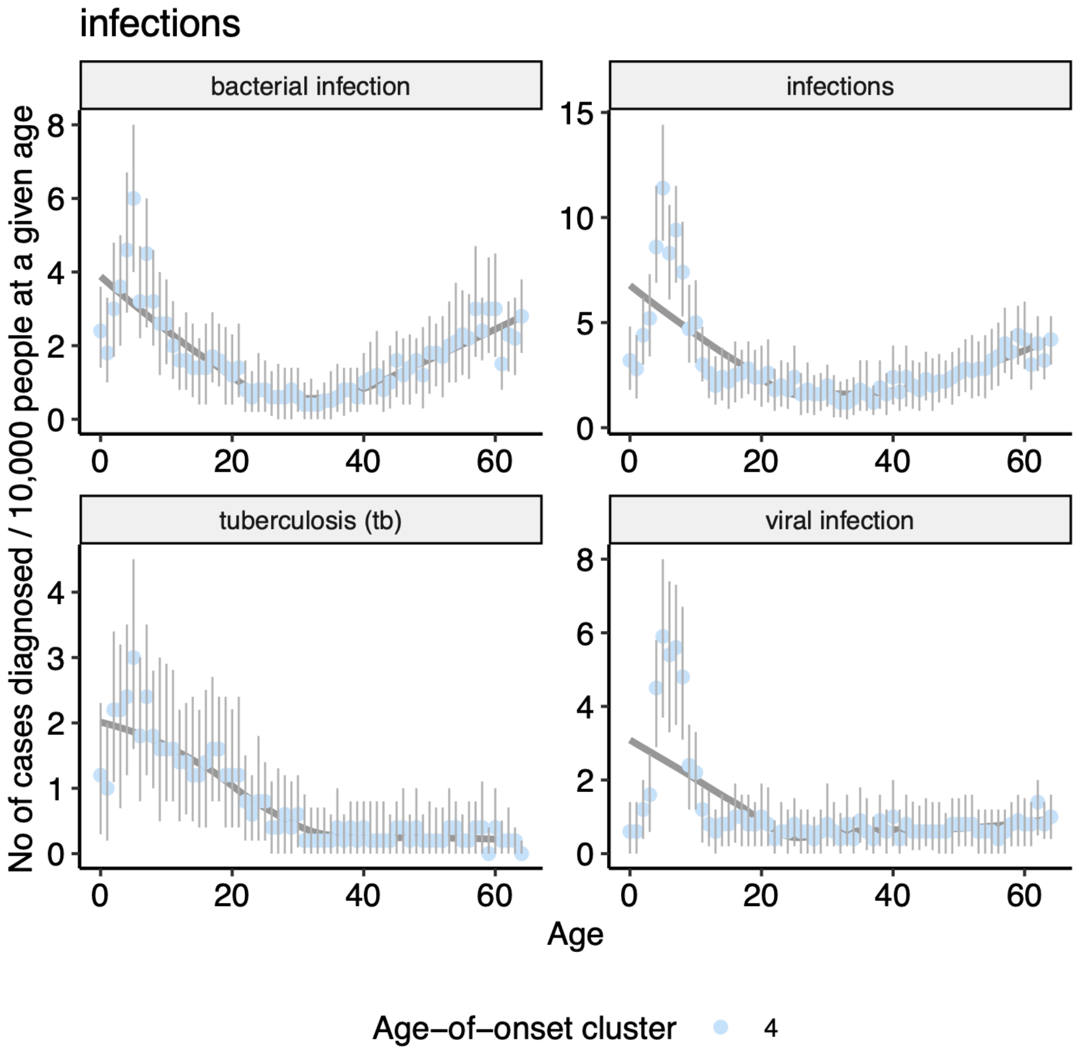


Figure S15: Same as Figure S10, but for infections.

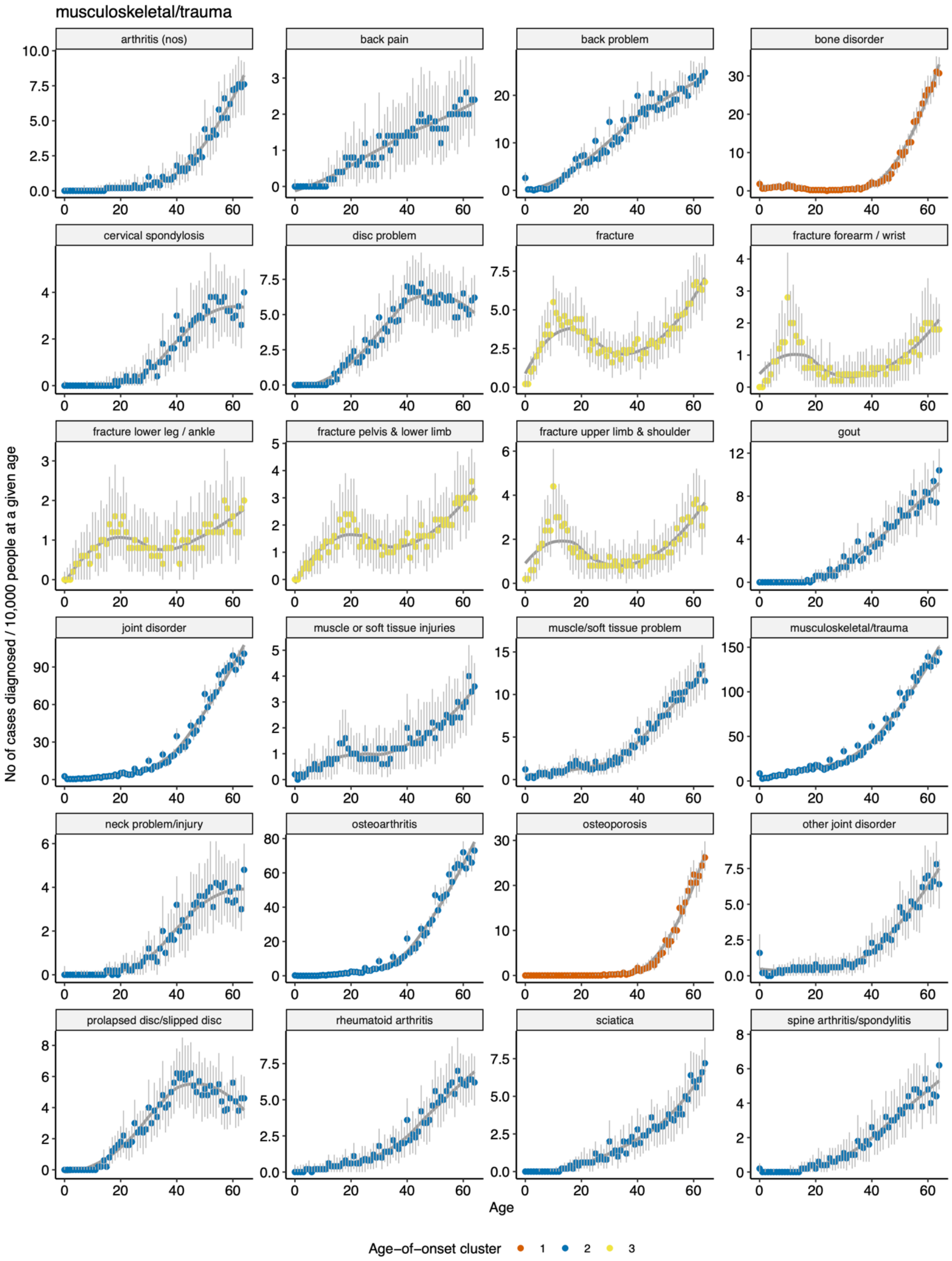


Figure S16: Same as Figure S10, but for musculoskeletal / trauma diseases.

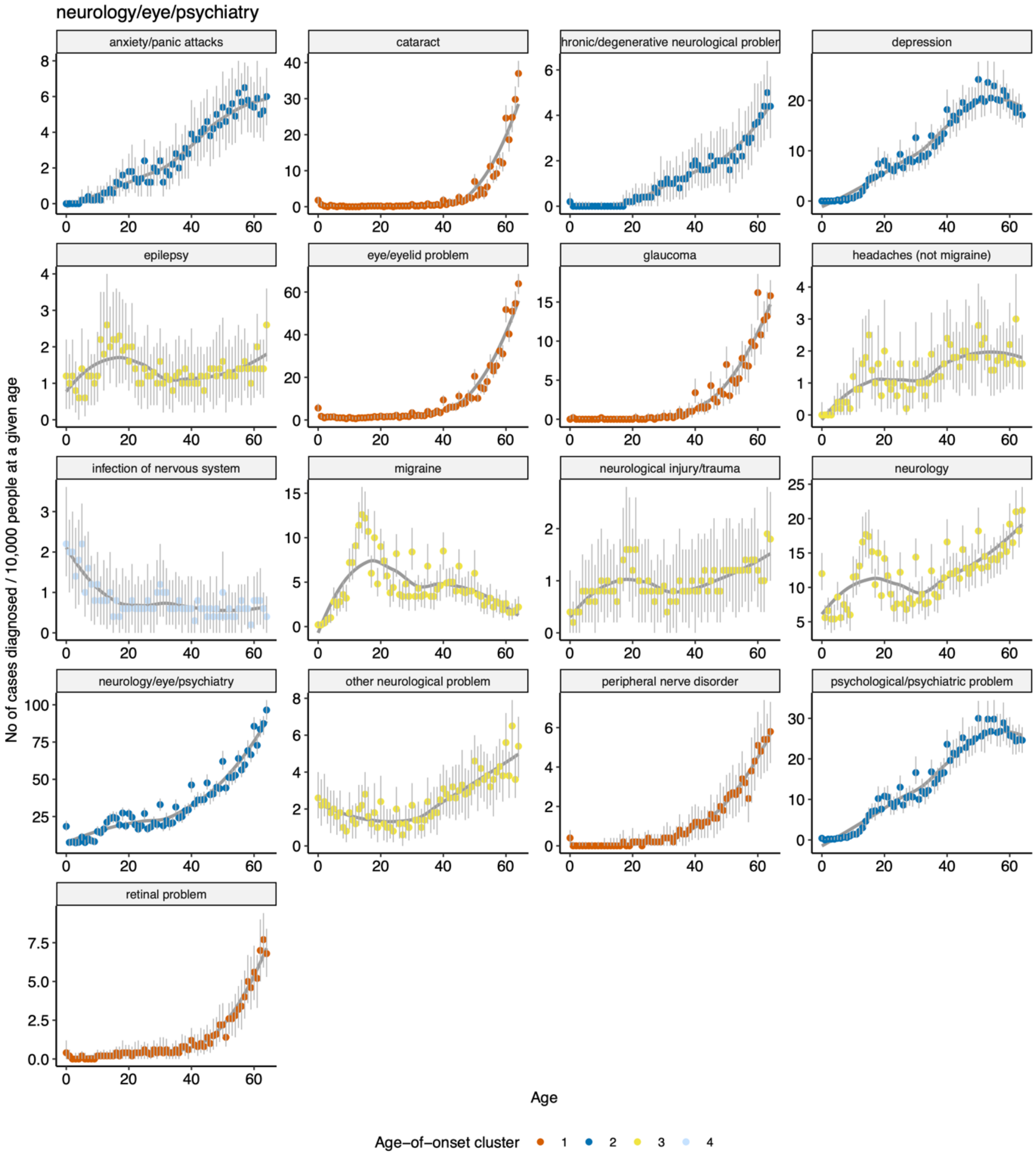


Figure S17: Same as Figure S10, but for neurology / eye / psychiatry diseases.

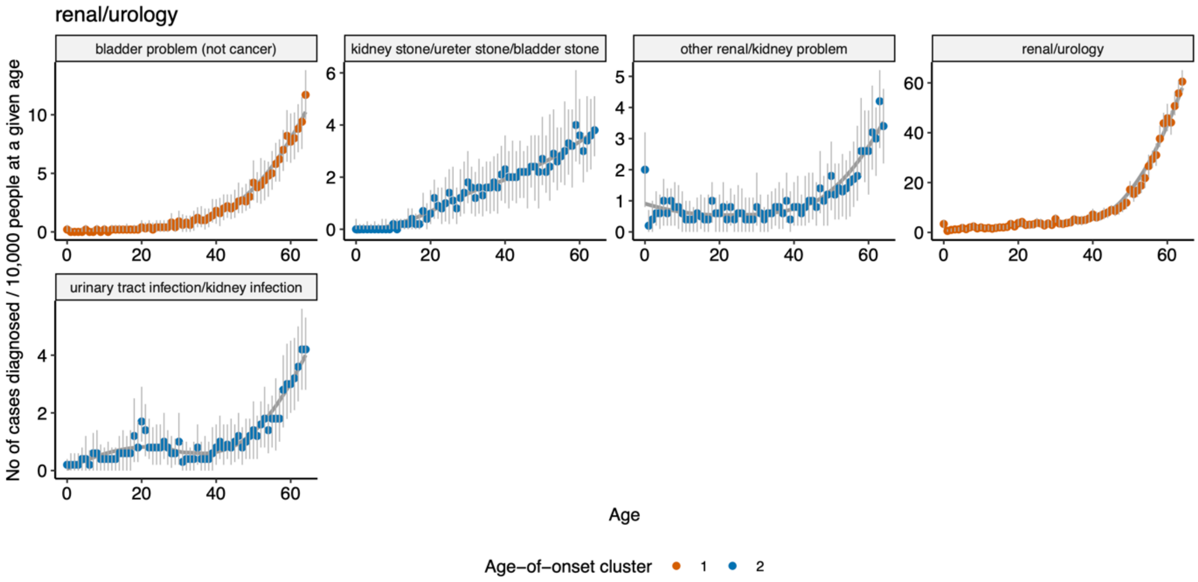


260
261 Figure S18: Same as Figure S10, but for renal / urology diseases.

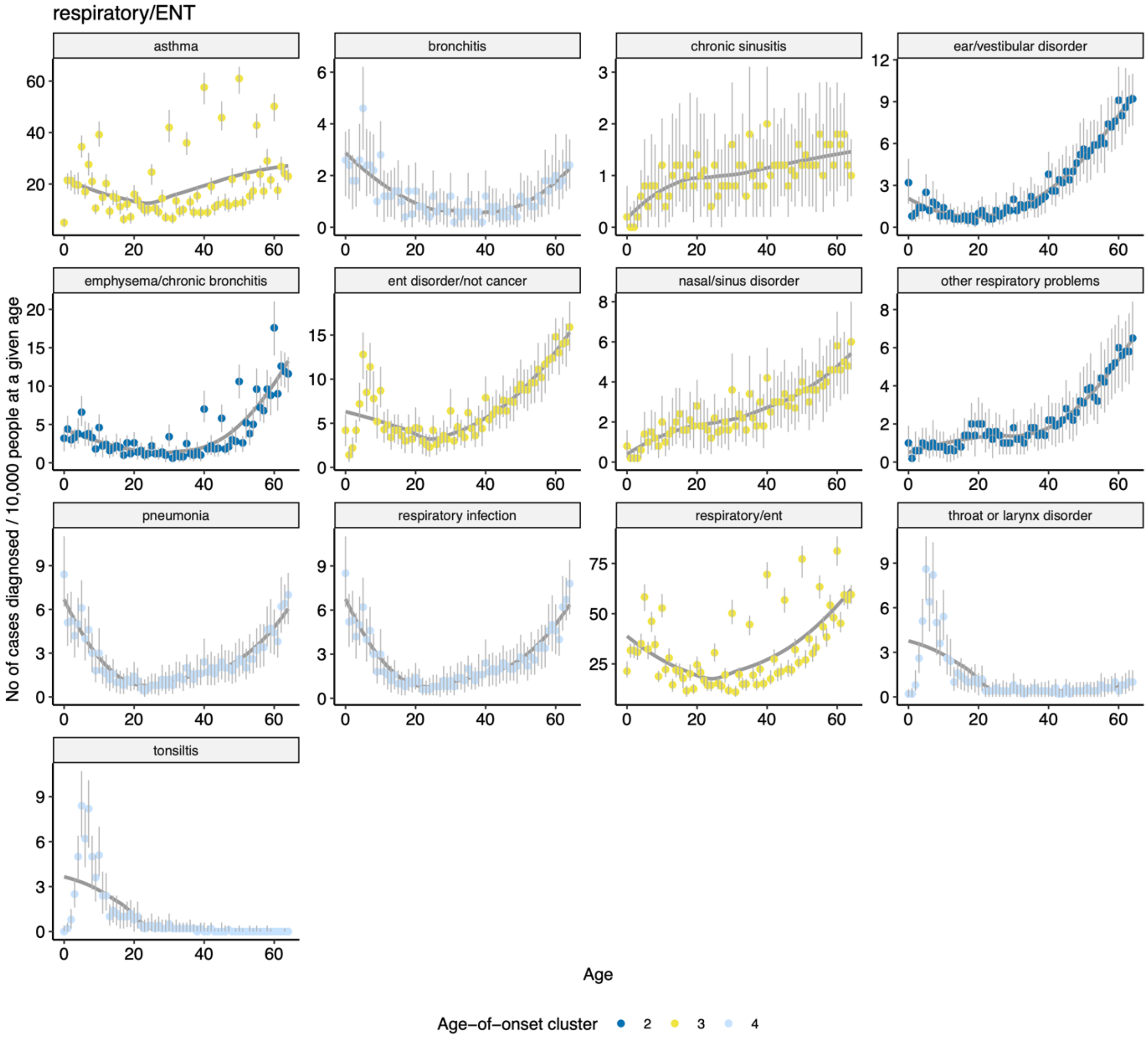


262
263 Figure S19: Same as Figure S10, but for respiratory / ENT diseases.
264
265

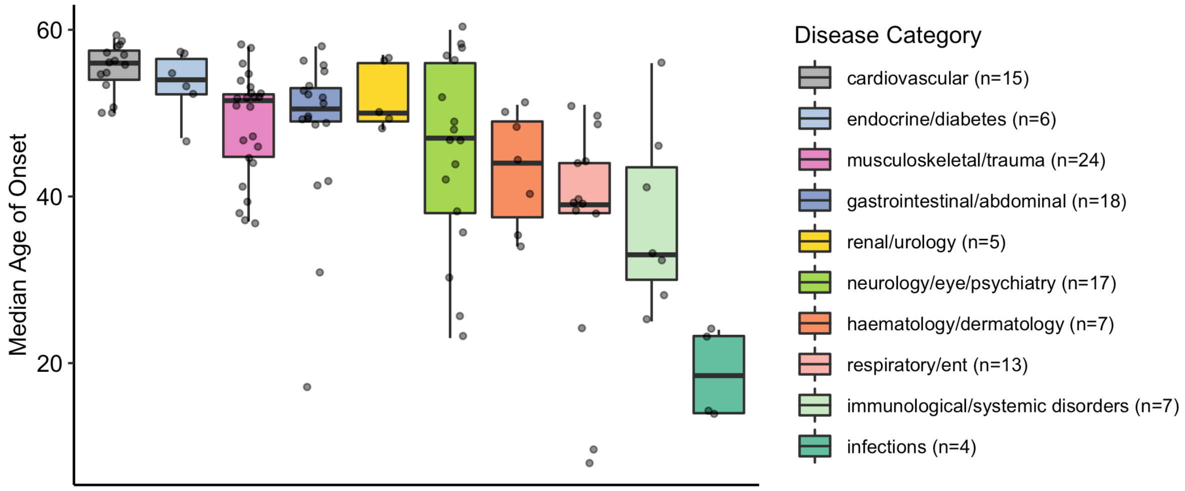


Figure S20: Distribution of median age-of-onset (y-axis) across categories (x-axis). Points show diseases, grouped by the categories (individual boxplots). Categories are ordered by the median value of the median age-of-onset.

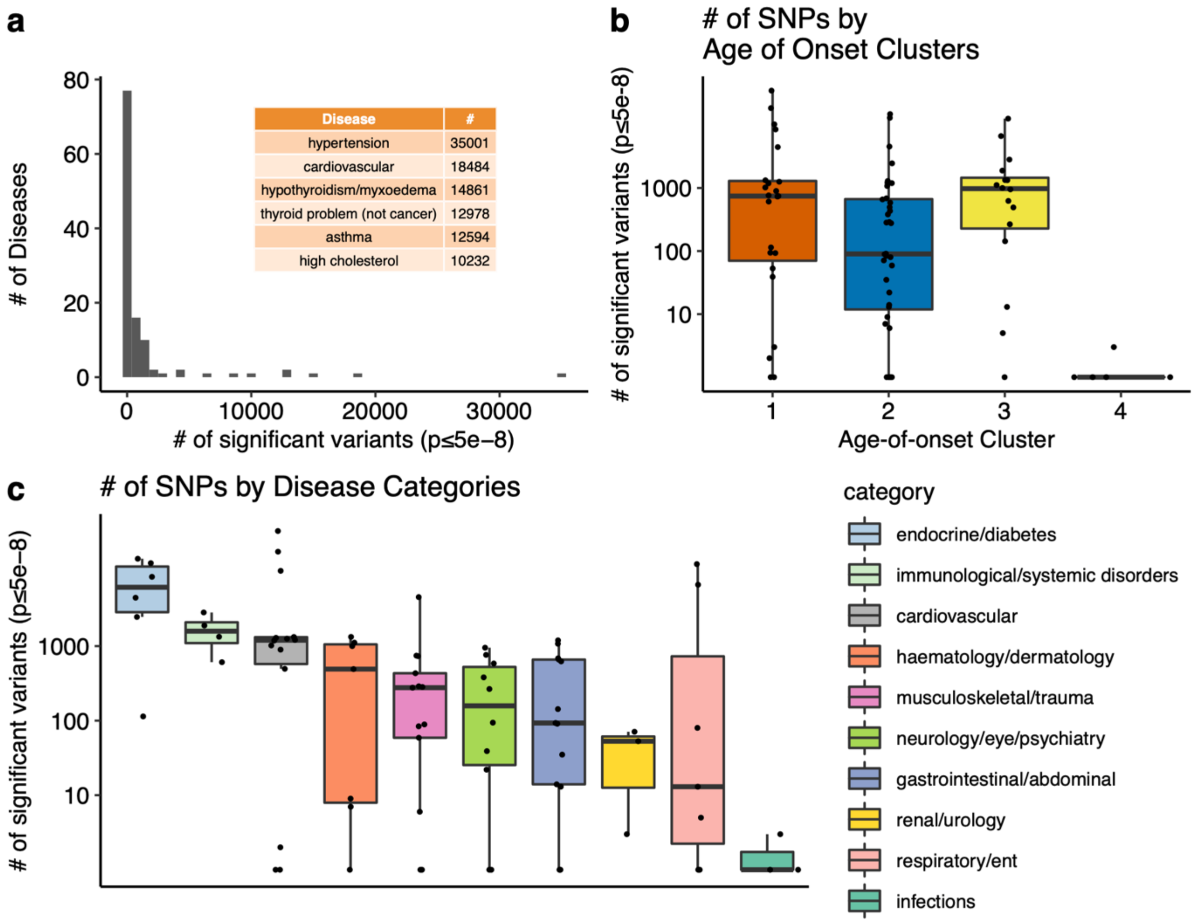


Figure S21: a) Number of diseases for different number of significant variants (p≤5e-8). Diseases with the highest number of associations (N≥10,000) are given as an inset table. b) Comparison of the number of significant associations (y-axis, on a log scale) across age-of-onset clusters (x-axis) (ANOVA after excluding cluster 4, p = 0.06). Since the y-axis is on a log scale, diseases with zero significant associations are not shown on the graph. c) The same as b) but for disease categories. Categories are ordered by the median number of significant SNPs.

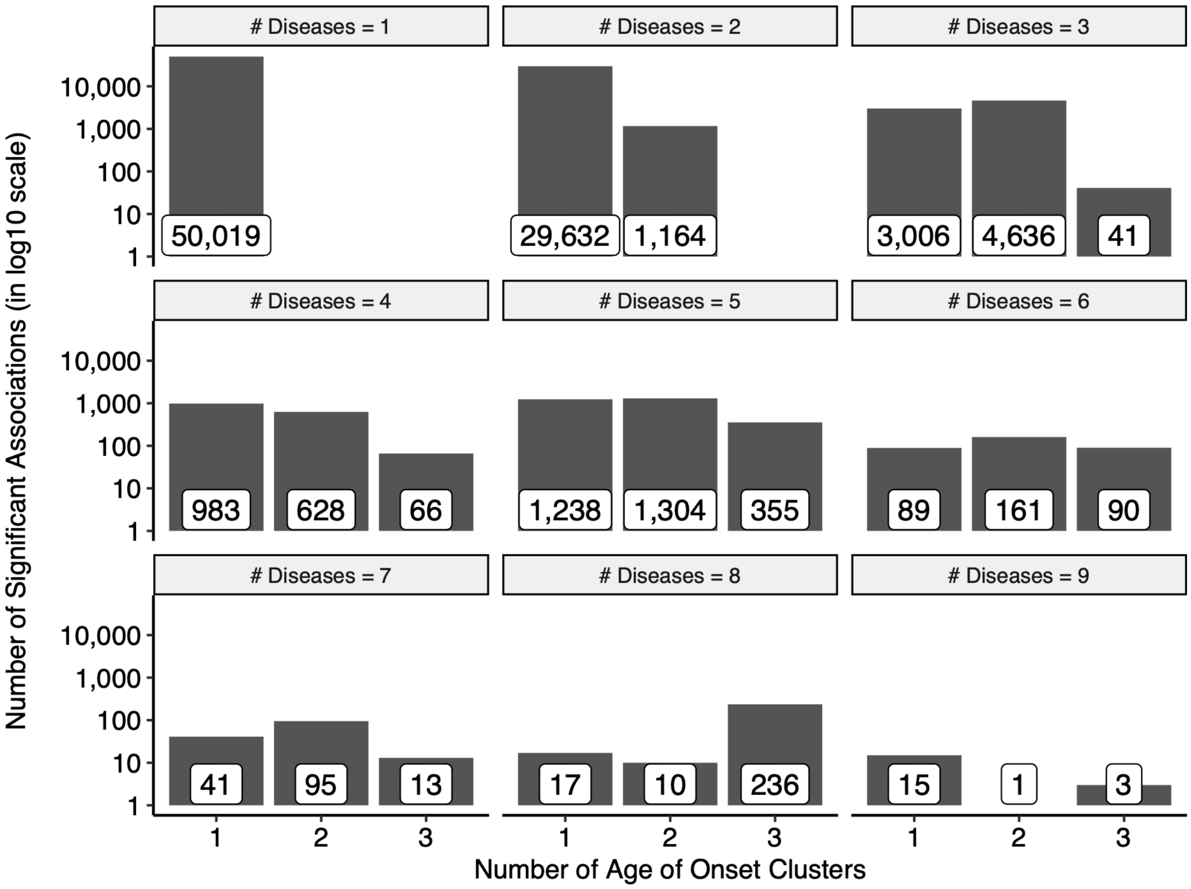


Figure S22: Distributions of the number of significant associations (y-axis) according to the number of diseases associated with a given SNP and the number of age-of-onset clusters (x-axis). For example, the upper left plot indicates that 50,019 polymorphisms are significantly associated with one disease in one age-of-onset cluster, while the lower right plot shows that there are 15, 1, and 3 significant SNPs associated with 9 diseases in one, two, or three age-of-onset clusters, respectively.

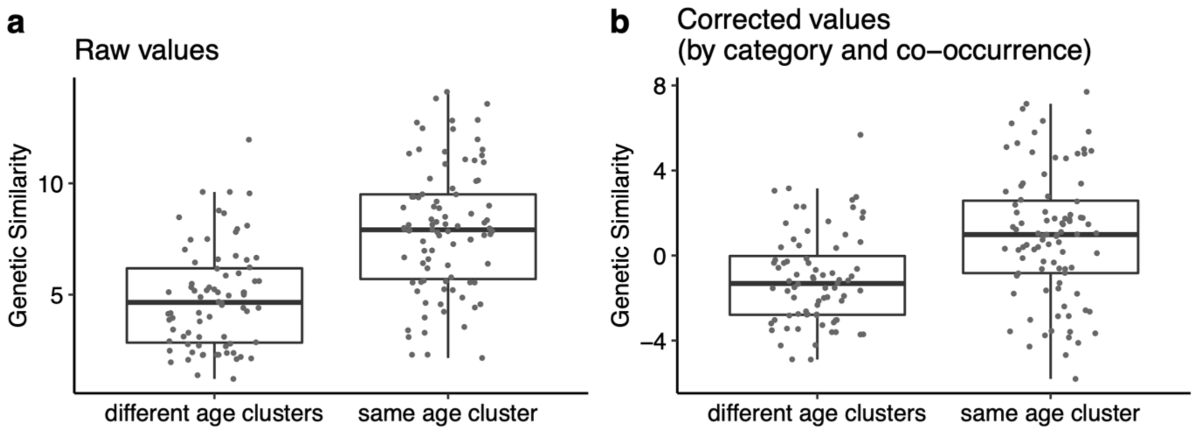


Figure S23: a) The difference between genetic similarity within and across age-of-onset clusters. Y-axis shows the genetic similarity (see Methods). b) The same as a) but the y-axis is corrected for disease category and co-occurrence using a linear model. This panel is the same as Figure 2b and given here only for an easier comparison.

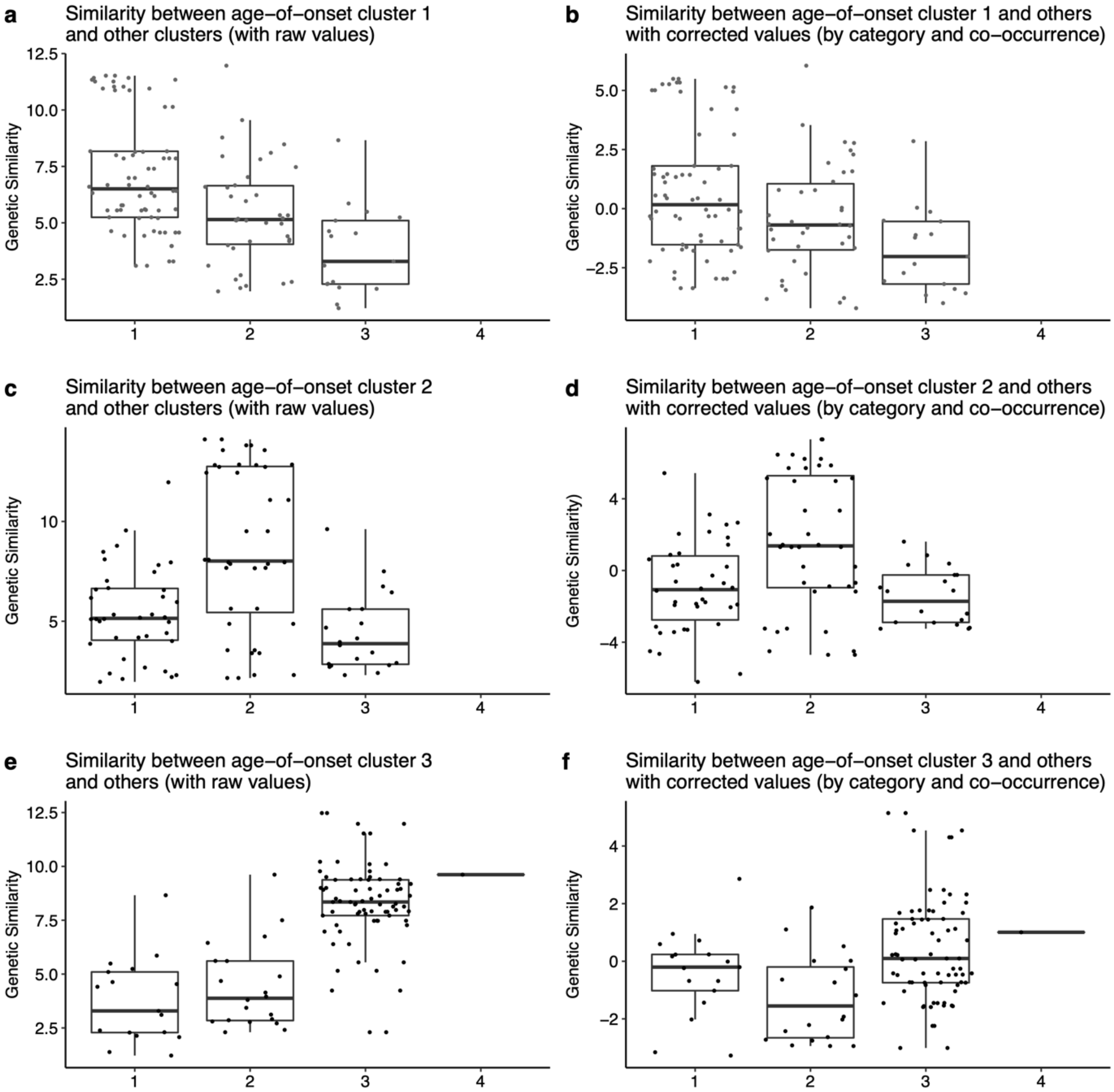


Figure S24: Genetic similarities between cluster 1 (a, b), 2 (c, d), 3 (e, f) and other age-of-onset clusters. The y-axis shows the genetic similarity on a log2 scale as the raw values (a, c, e) or as values corrected for disease category and co-occurrence using a linear model (b, d, f) (see Methods for details).

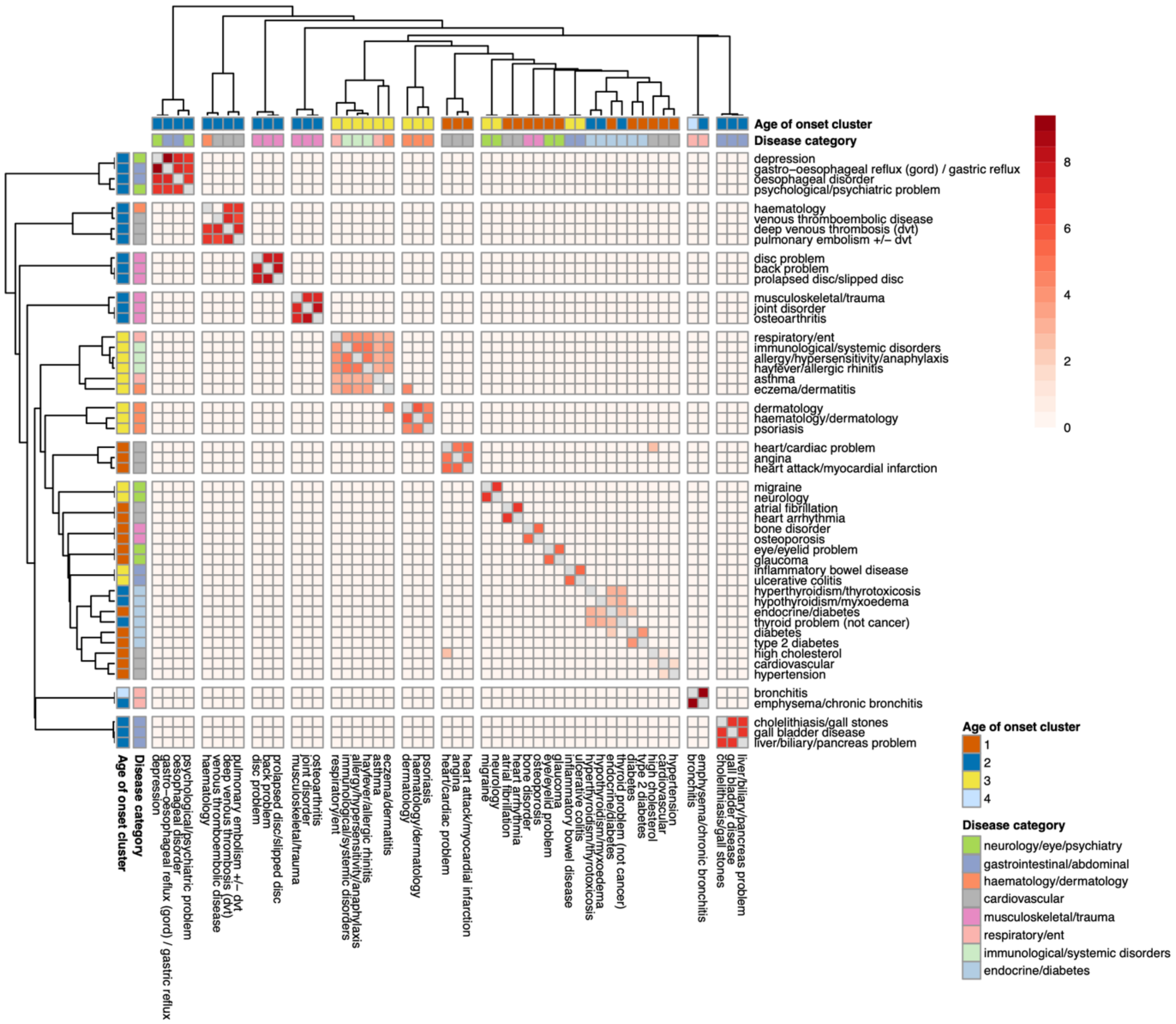


Figure S25: Significant genetic similarities (p≤0.01) calculated using independent LD blocks. Diseases (n=50) with at least one significant genetic similarity are displayed. The color shows the genetic similarity score, darker red means a higher score. Annotation columns show the age-of-onset clusters and disease categories. The diseases are clustered by the hierarchical clustering of genetic similarity scores.

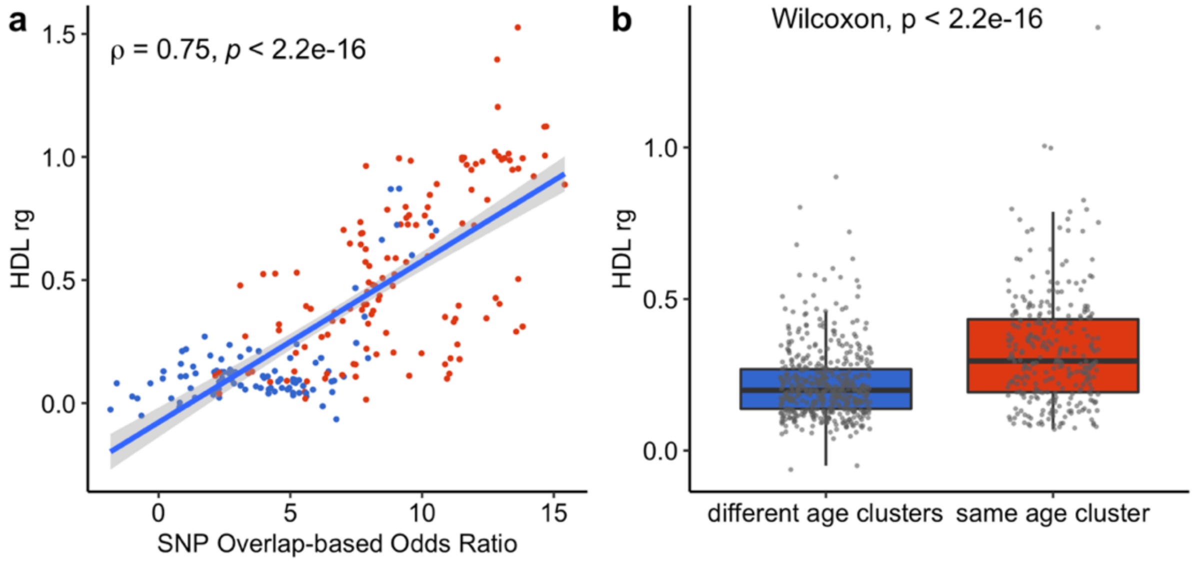


Figure S26: a) The correlation between the genetic similarity scores calculated using the SNP overlap-based odds ratio (x-axis) and HDL (y-axis). Blue points show the similarities calculated between diseases in different age of onset clusters and red points show the similarities calculated between diseases in the same age of onset cluster.

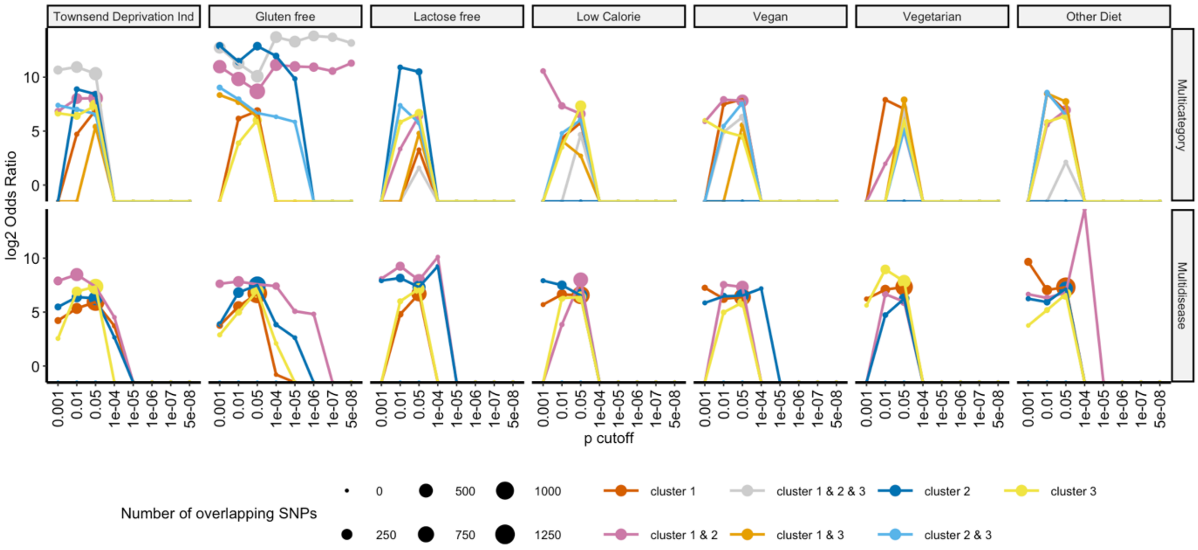


Figure S27: Overlap between SNPs associated with multiple disease or disease categories in specific age-of-onset clusters (color-coded) and Townsend deprivation index or specific diet regimes. The y-axis shows the log 2 transformed Odds Ratio and the x-axis shows the p-value cutoff to consider SNPs associated with Townsend deprivation index (*i.e.*, a material deprivation index incorporating unemployment, car and home ownership, and household overcrowding) or specific diet regimes. The size of the points shows the number of overlapping SNPs.

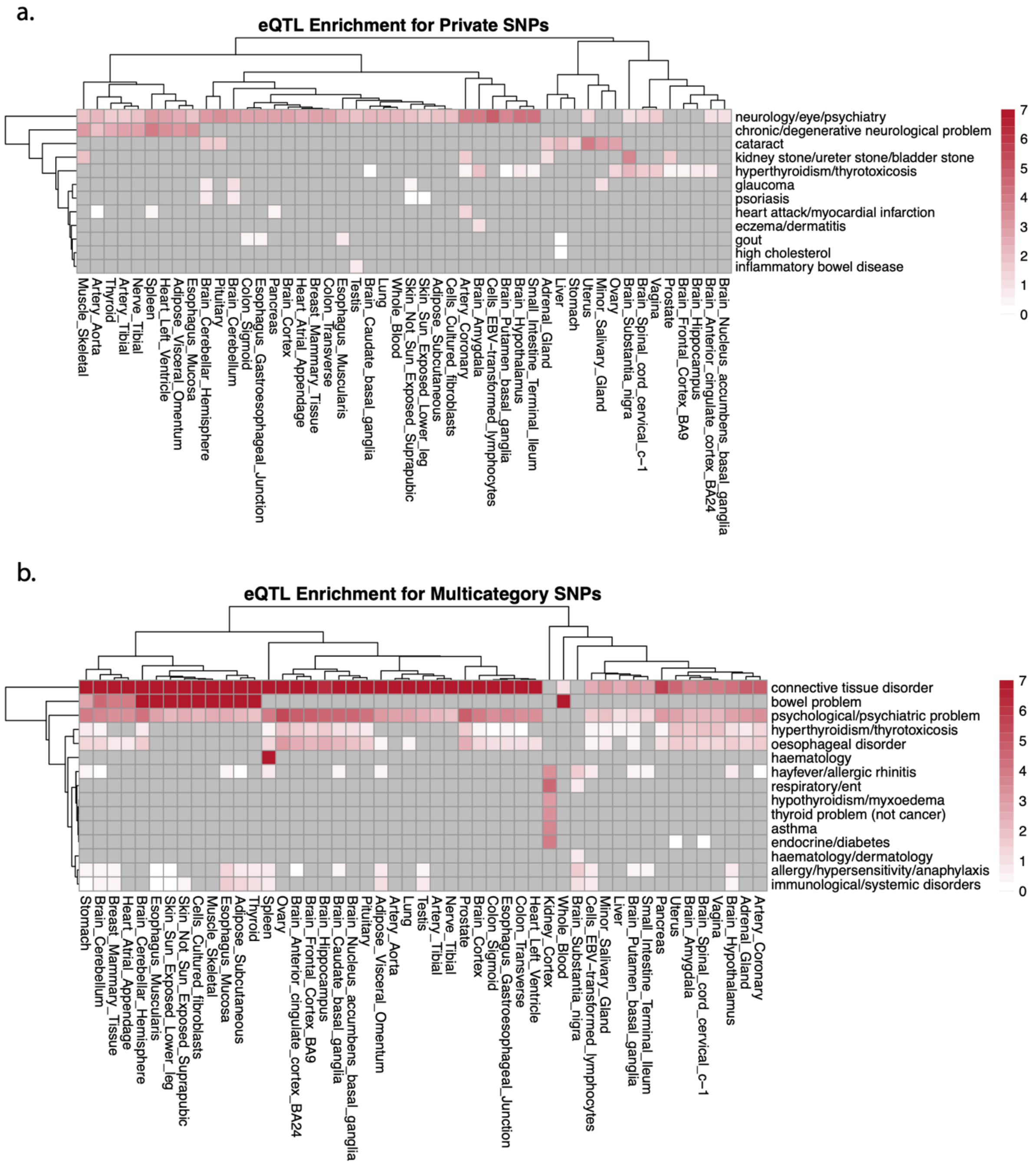


Figure S28: Heatmap showing the enrichment of disease associated (y-axis) SNPs among eQTLs in each tissue (x-axis). Color shows the log2 odds ratio and only the significant enrichment results (as determined by Fisher's exact test, FDR corrected p-value < 0.1). a) Enrichment for only the disease-specific (private) SNPs, b) Enrichment for Multicategory SNPs (*i.e.*, SNPs associated with multiple diseases spanning multiple disease categories).

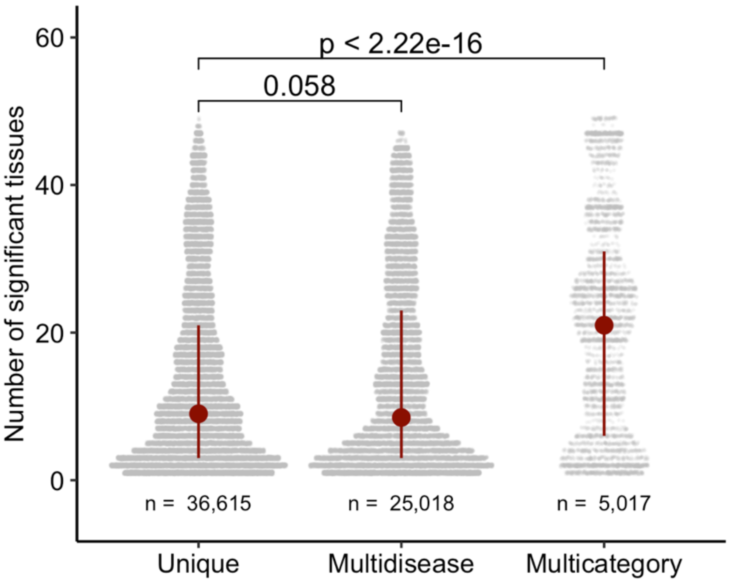


Figure S29: The distribution of the number of significant eQTL-tissue associations for s unique, multidisease, or multicategory SNPs are associated with (based on GTEx v8 data). P-values are calculated using Wilcoxon test. Dark red segments show the range between 1st and 3rd quartiles and the points show the median.

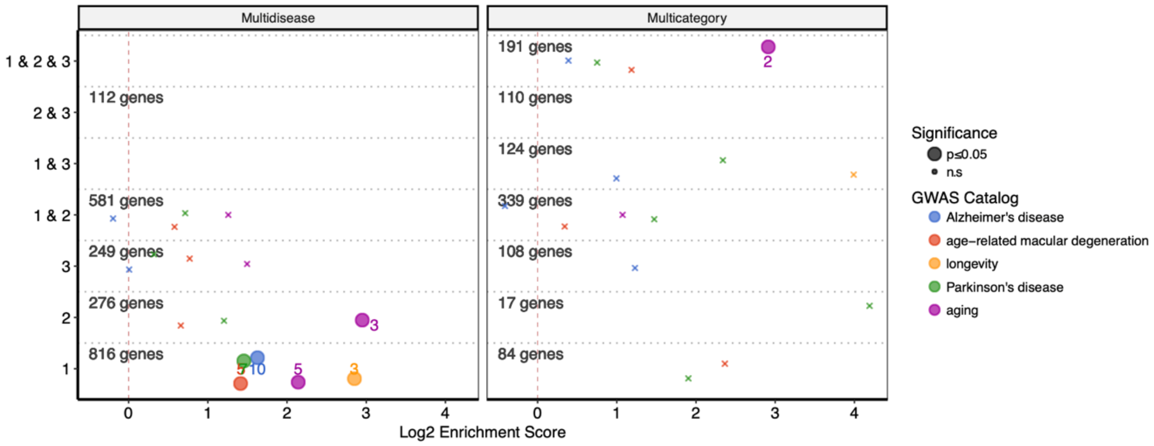


Figure S30: Overlap between genes associated with selected few aging-related traits and genes associated with diseases in different clusters. The x-axis shows log2 enrichment score, and the y-axis shows the age-of-onset clusters. The numbers of genes in each cluster (for both multidisease and multicategory genes) are given. The size of the points shows the statistical significance (large points show marginal p-value≤0.05, small 'x' indicates non-significant overlaps) and the color shows different aging-related GWAS Catalog traits. The colored numbers near the points show the numbers of overlapping genes.

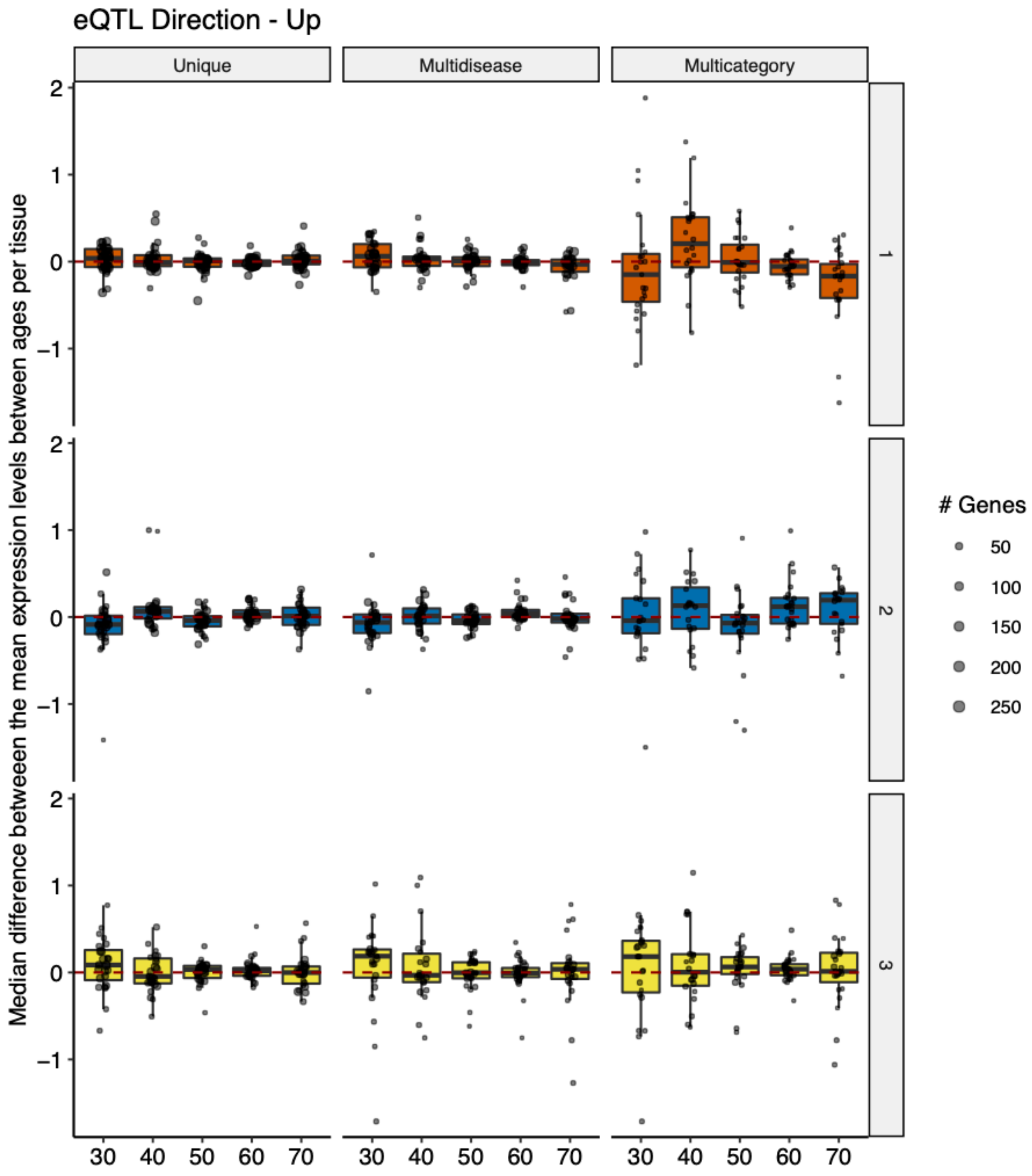


Figure S31: Age-related expression changes of the genes with significant eQTLs associated with unique, multidisease, or multicategory diseases in different age-of-onset clusters. In order to match age-related expression changes with the variants, tissue-specific eQTL data was used. More specifically, disease-associated SNPs were first filtered to only include those with a positive association with the gene expression (*i.e.* eQTLs associated with increased gene expression). Using GTEx expression data, mean gene expression values for each age group is calculated. Taking the difference between age groups, we calculated the expression difference at each break point (30, 40,...,70, x-axis). Differences for multiple genes for each tissue are summarized by taking the median expression difference (y-axis). Each point represents a tissue and the size indicate the number of genes averaged in that particular tissue.

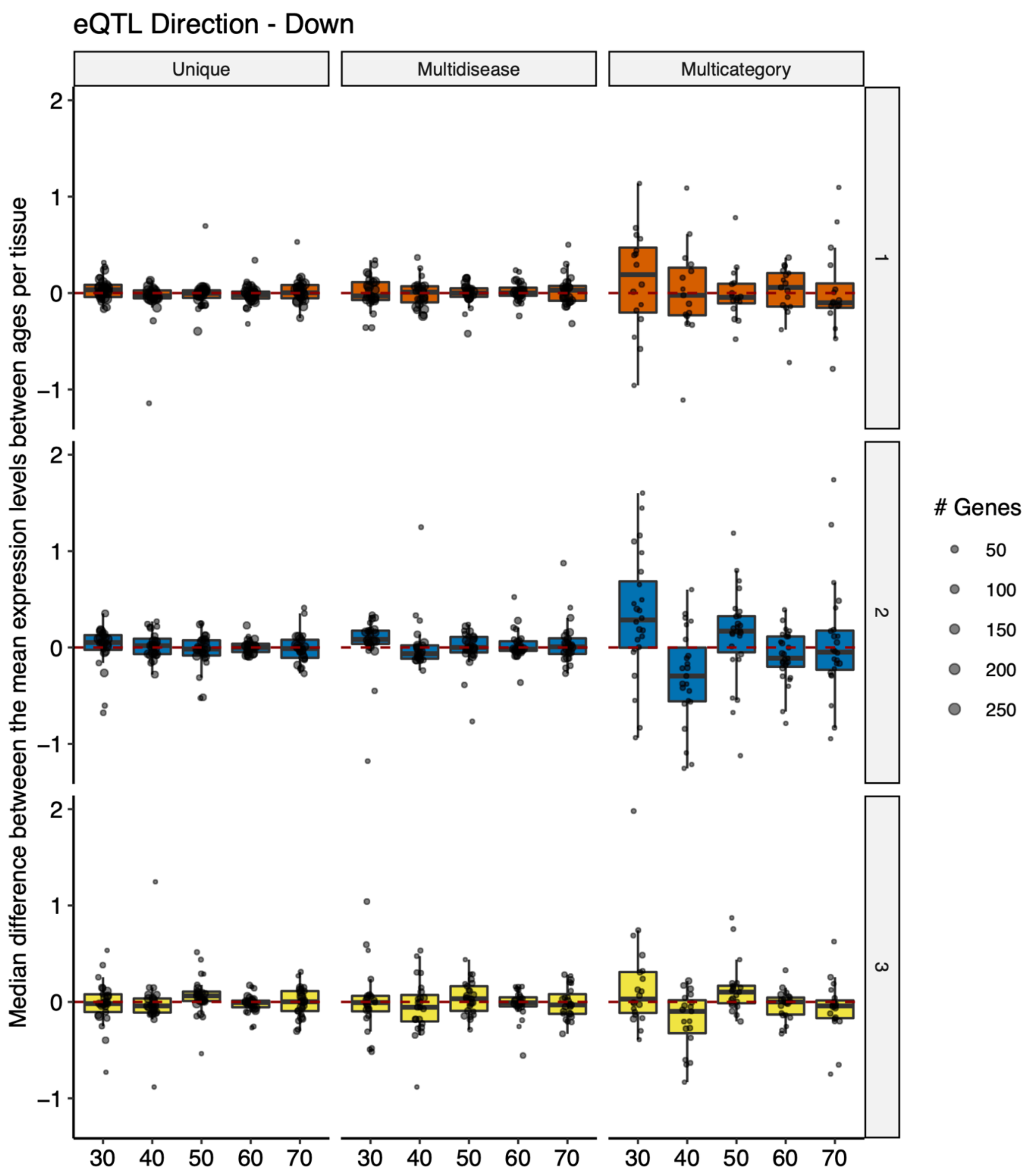


Figure S32: Age-related expression changes of the genes with significant eQTLs associated with unique, multidisease, or multicategory diseases in different age-of-onset clusters. In order to match age-related expression changes with the variants, tissue-specific eQTL data was used. More specifically, disease-associated SNPs were first filtered to only include those with a negative association with the gene expression (*i.e.* eQTLs associated with decreased gene expression). Using GTEx expression data, mean gene expression values for each age group is calculated. Taking the difference between age groups, we calculated the expression difference at each break point (30, 40,...,70, x-axis). Differences for multiple genes for each tissue are summarized by taking the median expression difference (y-axis). Each point represents a tissue and the size indicate the number of genes averaged in that particular tissue.

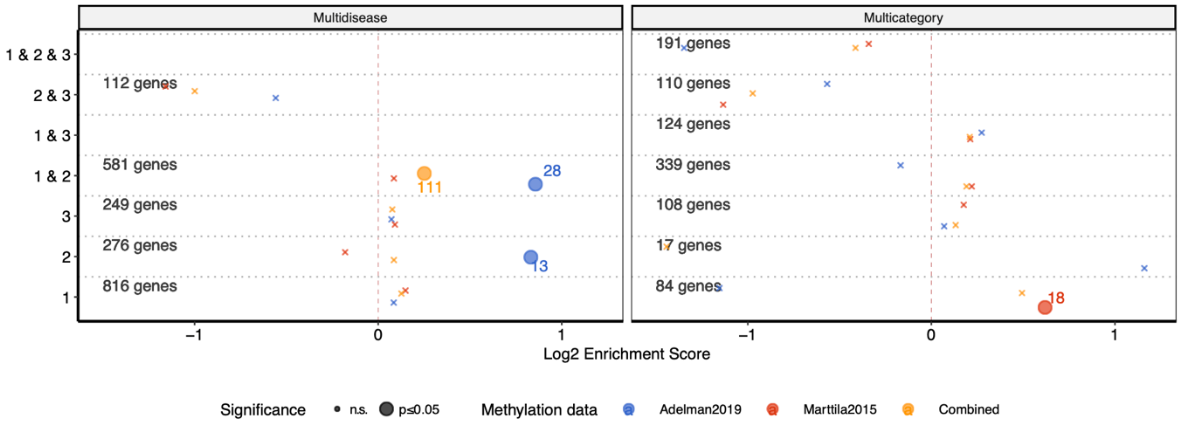


Figure S33: Overlap between differentially methylated genes during ageing and genes associated with diseases in different clusters. The x-axis shows log2 enrichment score, and the y-axis shows the age-of-onset clusters. The numbers of genes in each cluster (for both multidisease and multicategory genes) are given. The size of the points shows the statistical significance (large points show marginal p-value≤0.05, small ‘x’ indicate non-significant overlaps) and the color shows different methylation datasets. The colored numbers near the points show the numbers of overlapping genes.

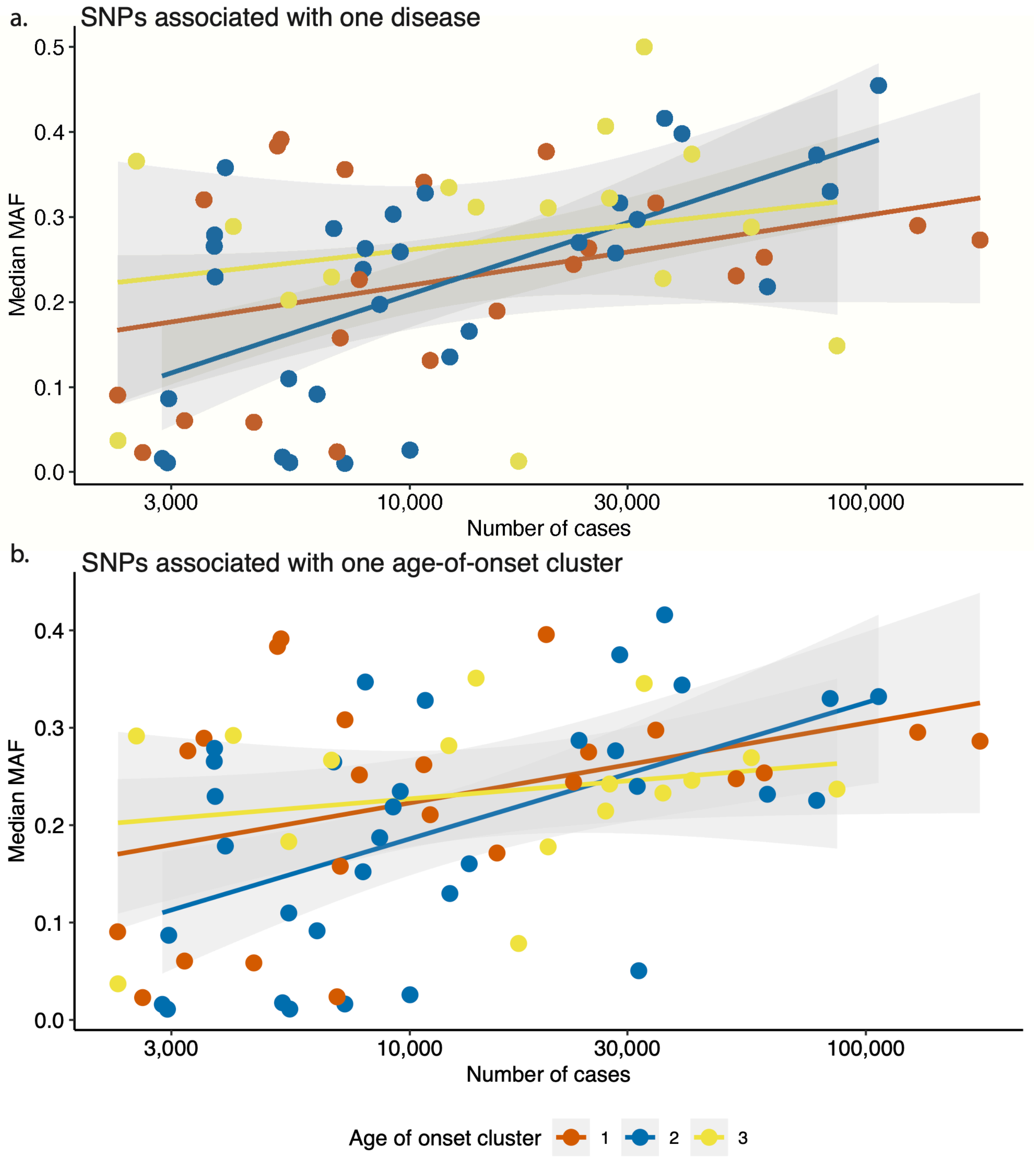


Figure S34: Association between median MAF (y-axis) and the number of cases (x-axis, log10 scale) for diseases (points) in different age-of-onset clusters (shown with different colors) for a) SNPs associated with only one disease, b) SNPs associated with any number of diseases within one cluster. Linear regression lines and standard errors of the lines are shown for each age-of-onset cluster separately.

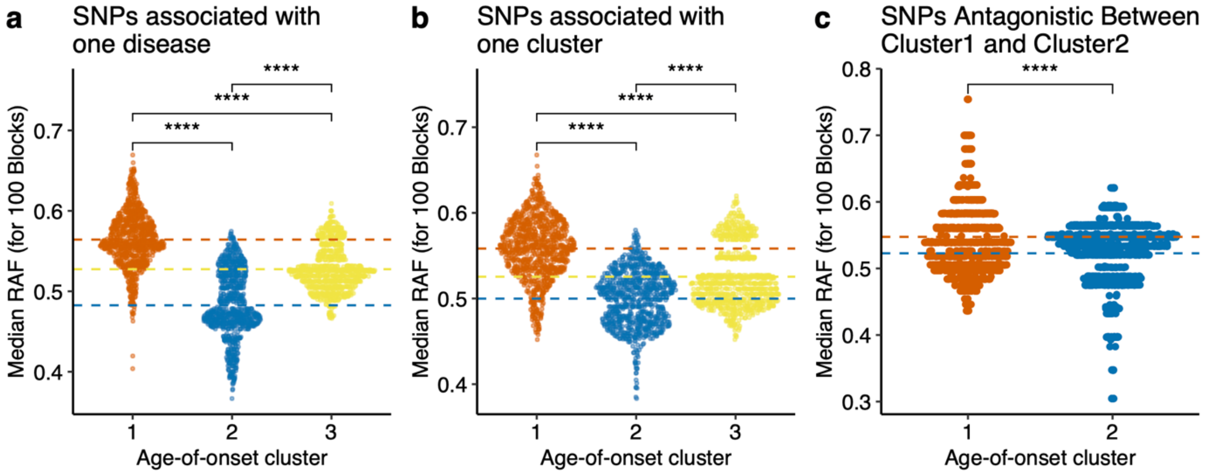


Figure S35: The distribution of Median Risk Allele Frequencies (RAF, y-axis) for 100 randomly sampled LD blocks, for 1,000 times, using variants a) associated with one disease, b) associated with one cluster, c) with antagonistic association between cluster 1 and cluster 2. *ns: p>0.05, *:p≤0.05, **: p≤0.01, ***: p≤0.001, ****: p≤0.00001*

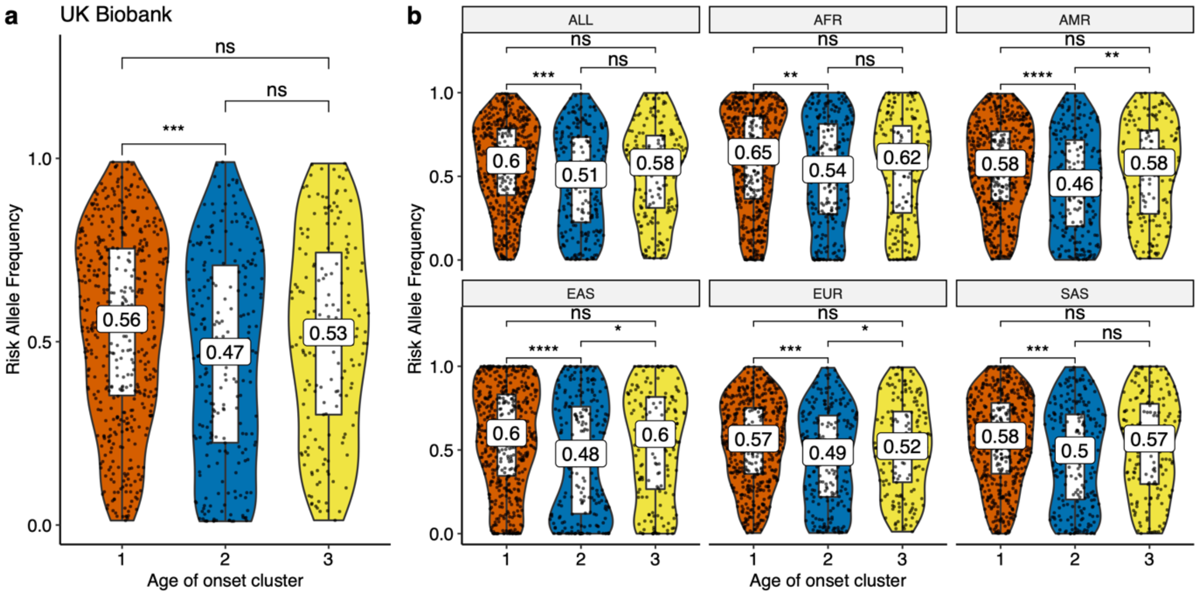


Figure S36: a) Risk allele frequency distributions (y-axis) of different age-of-onset clusters (x-axis) in UK Biobank for SNPs associated with one disease. This plot is the same as Figure 4a, and included here for an easier comparison. b) The same as panel a but for different 1000 Genomes super-populations (ALL: complete 1000 Genomes cohort, AFR: African, AMR: Ad Mixed American, EAS: East Asian, EUR: European, SAS: South Asian). *ns: p>0.05, *:p≤0.05, **: p≤0.01, ***: p≤0.001, ****: p≤0.00001*

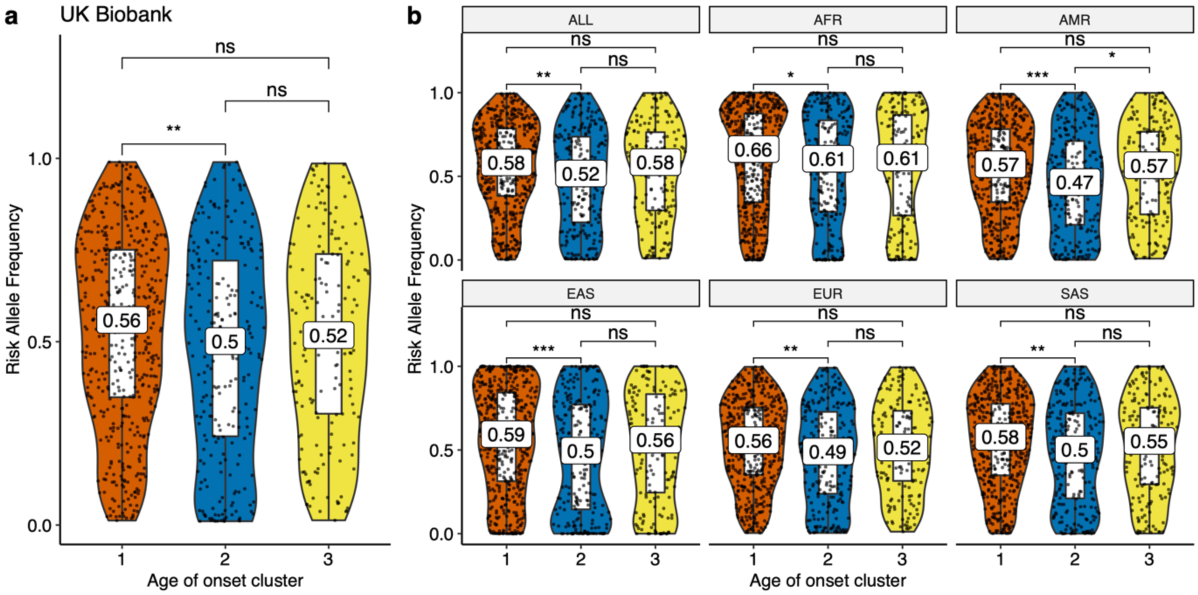


Figure S37: a) Risk allele frequency distributions (y-axis) of different age-of-onset clusters (x-axis) in UK Biobank for SNPs associated with one cluster, excluding antagonistic associations. This plot is the same as Figure 4b, and included here for an easier comparison. b) The same as a) but for different 1000 Genomes super-populations (ALL: complete 1000 Genomes cohort, AFR: African, AMR: Ad Mixed American, EAS: East Asian, EUR: European, SAS: South Asian). *ns: p>0.05, *:p≤0.05, **: p≤0.01, ***: p≤0.001, ****: p≤0.00001*

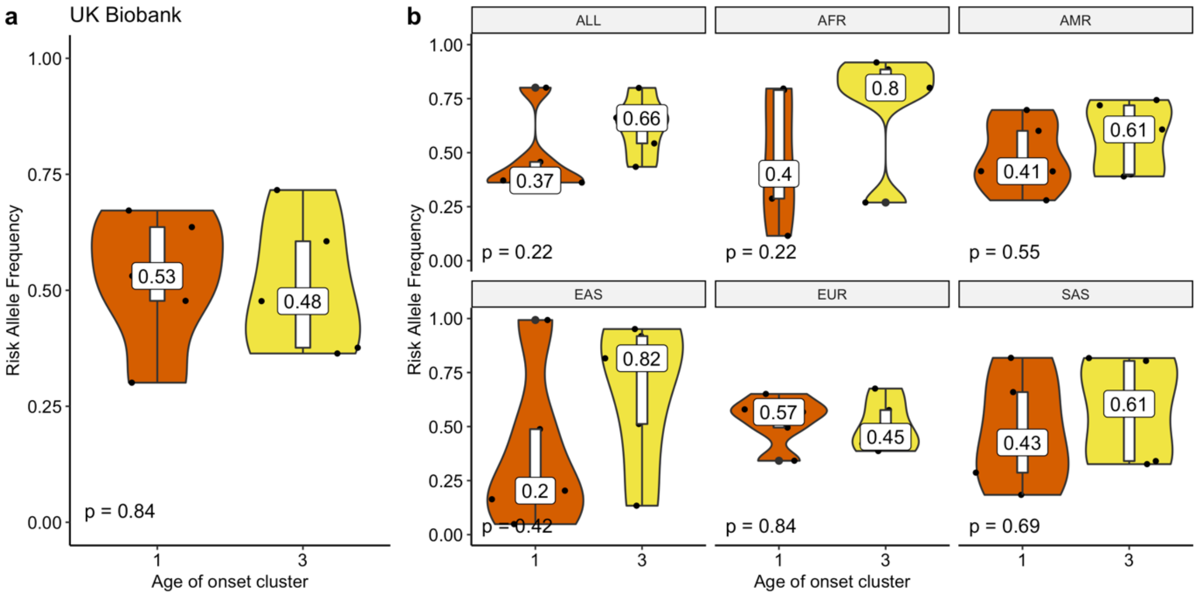


Figure S38: Risk allele frequency distributions (y-axis) for the age-of-onset cluster 1 and 3 in the UKBB (x-axis) for a) SNPs that have antagonistic association with cluster 1 and 3 (excluding agonists between cluster 1 and 3). b) The same as panel a but for different 1000 Genomes super-populations (ALL: complete 1000 Genomes cohort, AFR: African, AMR: Ad Mixed American, EAS: East Asian, EUR: European, SAS: South Asian).

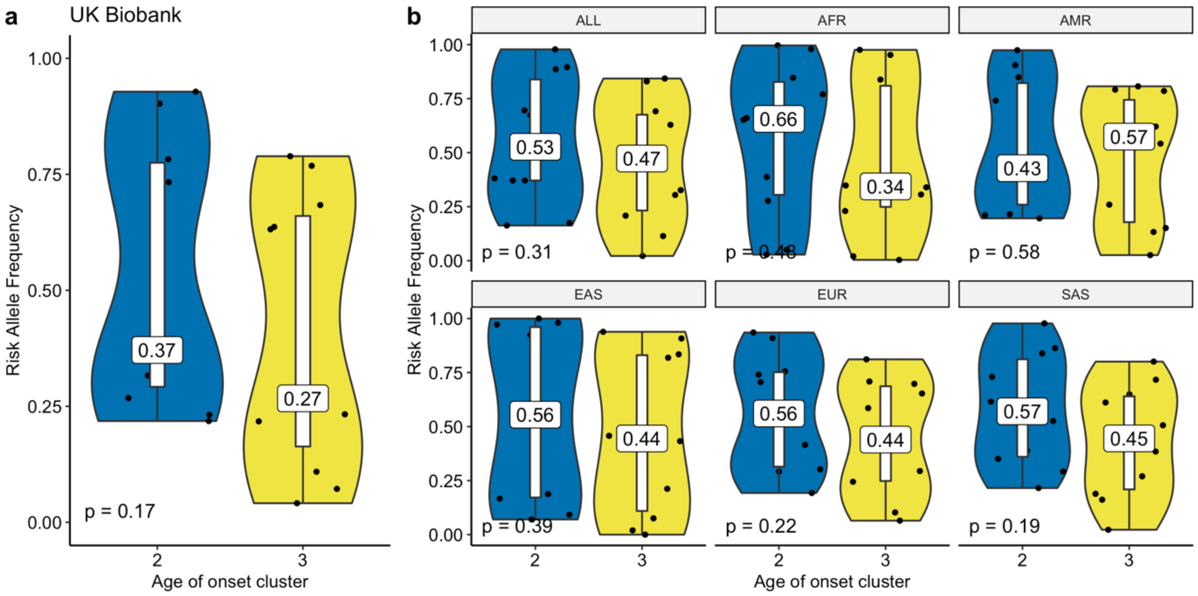


Figure S39: Risk allele frequency distributions (y-axis) for the age-of-onset cluster 2 and 3 in the UKBB (x-axis) for a) SNPs that have antagonistic association with cluster 2 and 3 (excluding agonists between cluster 2 and 3). b) The same as panel a but for different 1000 Genomes super-populations (ALL: complete 1000 Genomes cohort, AFR: African, AMR: Ad Mixed American, EAS: East Asian, EUR: European, SAS: South Asian).

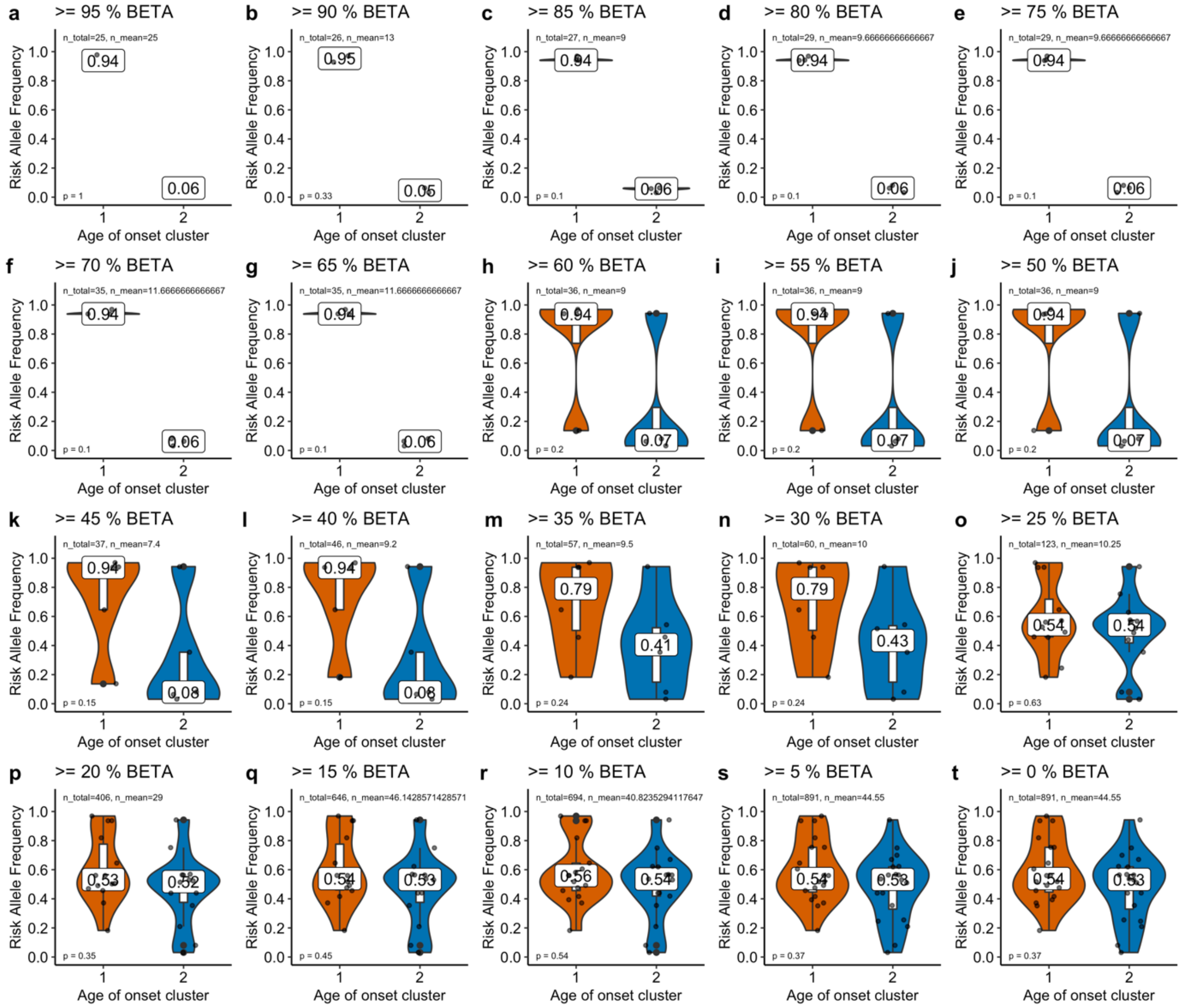


Figure S40: Risk allele frequencies in UK Biobank for the loci showing antagonistic associations between cluster 1 and cluster 2 filtered by different effect size cutoffs. The title of each plot shows the cutoff, where e.g. >=95 % BETA means only the SNPs with a BETA (effect size) value higher than 95% of all other antagonistic SNPs are used. >=0 % BETA means no filtering.

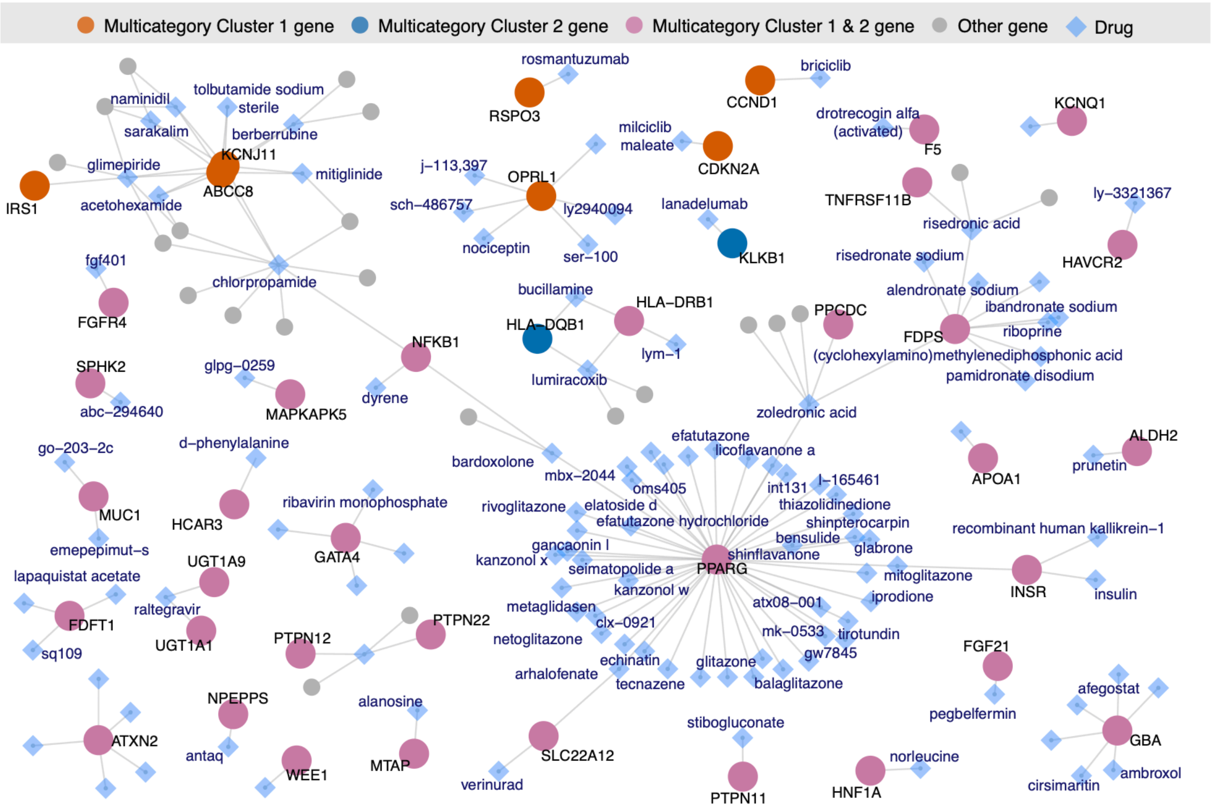


Figure S41: 'Drug-target gene' interaction network for the drugs that specifically target multicategory cluster 1, cluster 2 or cluster '1 & 2' genes as determined by Fisher's exact test. Blue diamonds show the drugs with significant association or targeting only one gene in these gene groups. Diamonds without written names are only represented with the ChEMBL IDs in the datasets and did not have names. Drug labels written in bold are drugs approved for different conditions. Circles represent the genes targeted by the significant hits, colored by their age-of-onset cluster. Gray circles show the genes targeted by these drugs but are not among the gene set of interest.

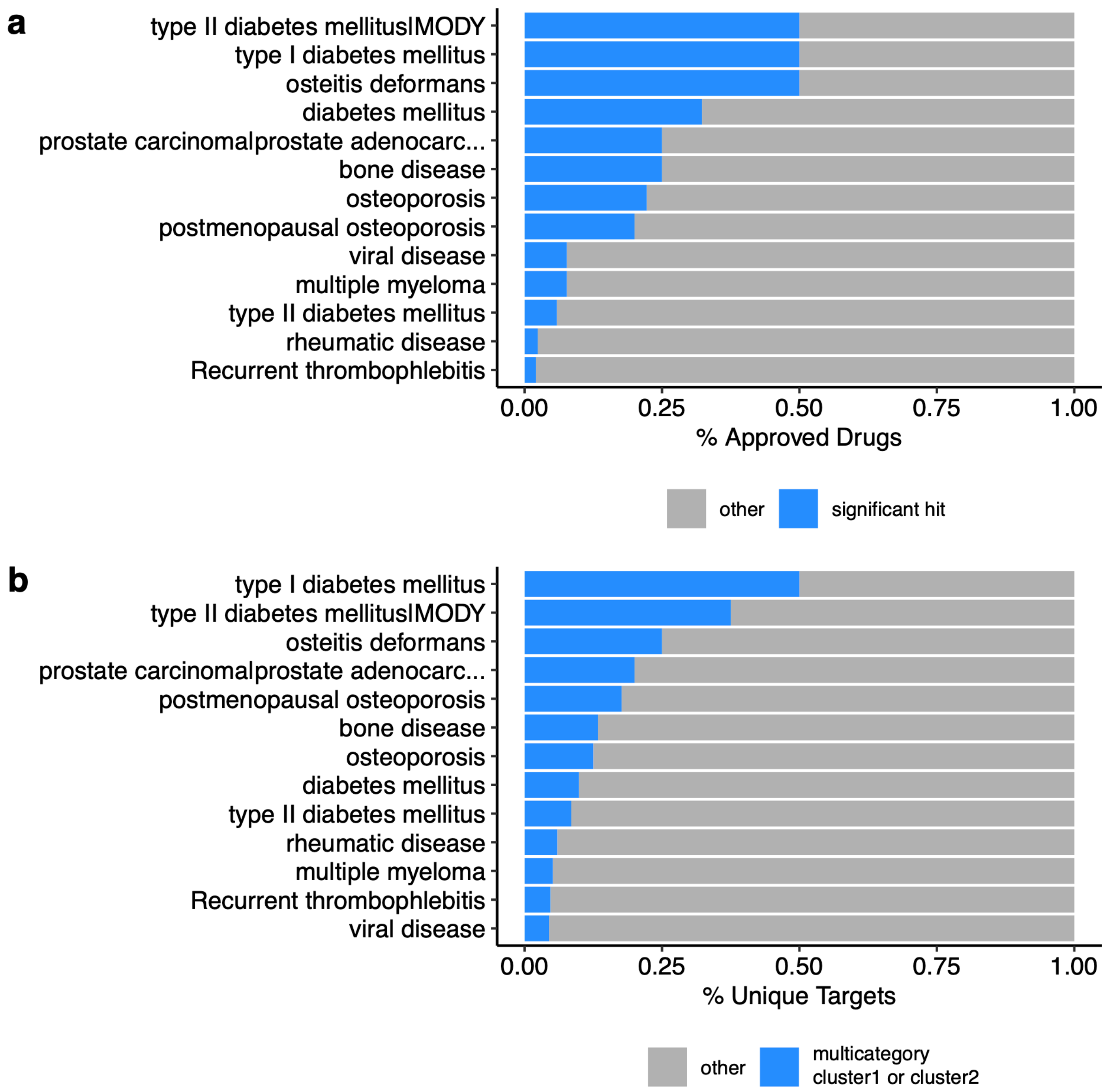


Figure S42: Distribution of a) the drugs and b) their targets approved for 13 conditions treated with the significant hits for drug repurposing. a) X-axis shows the proportion of the significant hits in the drug repurposing study approved for the treatment of the conditions listed on y-axis. b) The same as a) but showing the proportion of unique targets of the approved drugs (x-axis) for the conditions listed on the y-axis.

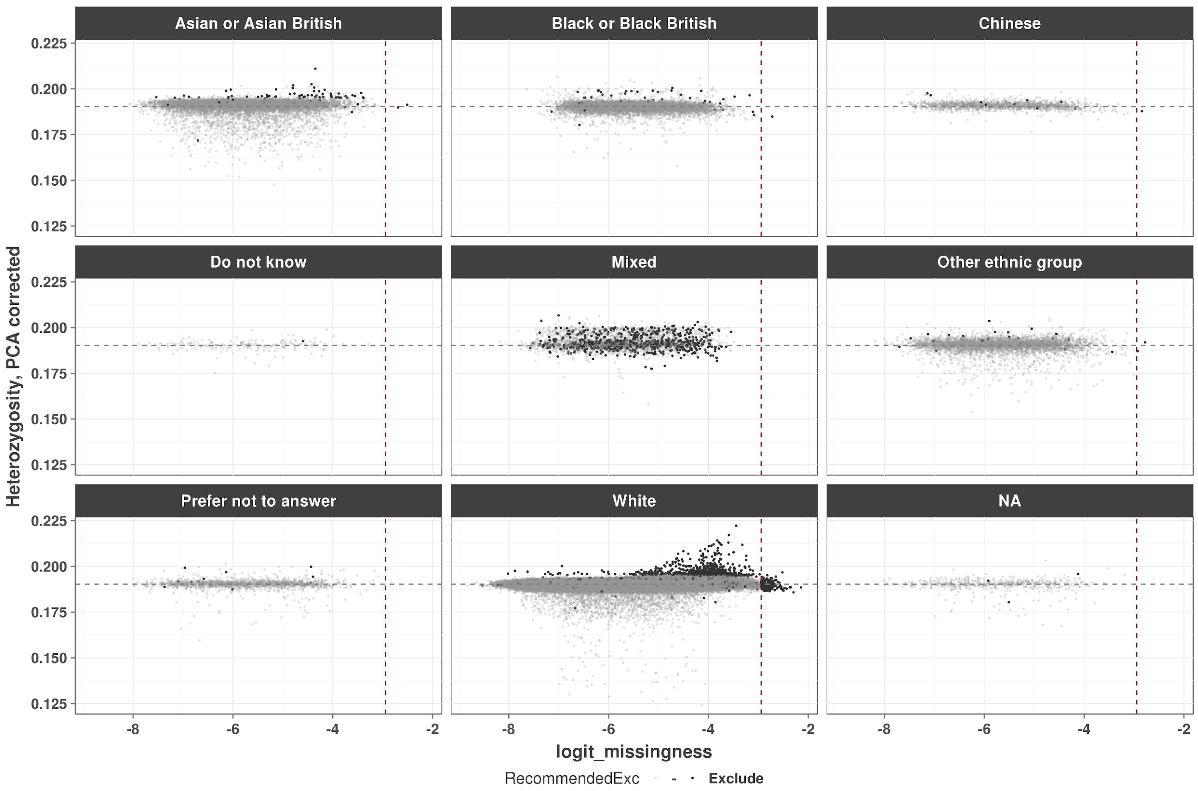


Figure S43: Scatter plot between logit(missingness) and PCA corrected heterozygosity measures. Each panel shows a self-declared ethnic background. Vertical red lines show the missing rate of 0.05, and horizontal grey lines show the average heterozygosity in UK Biobank.

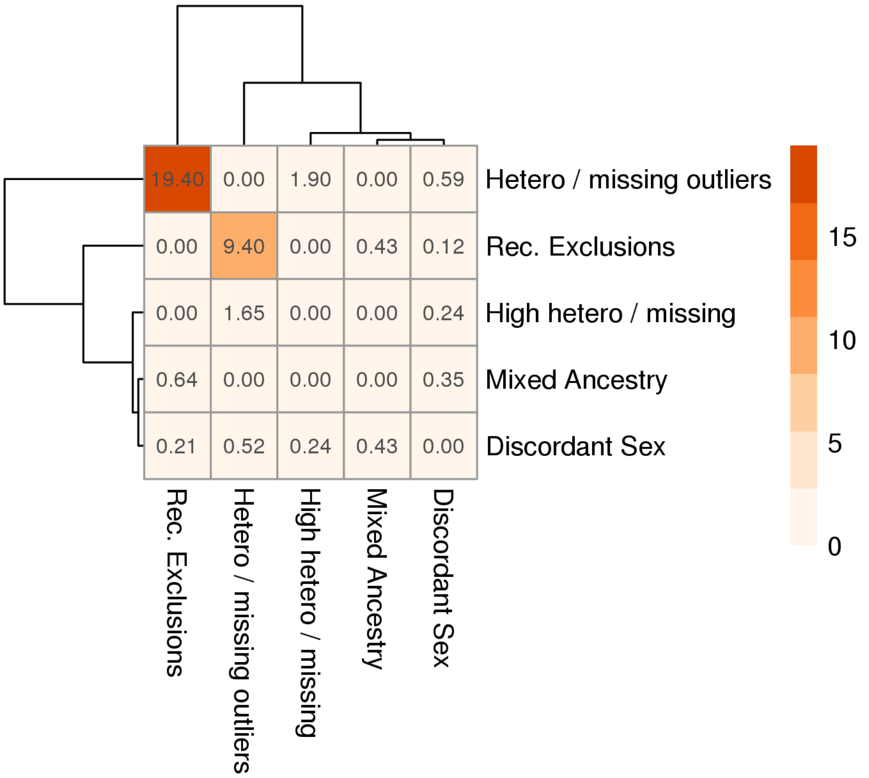


Figure S44: Heatmap showing the percent overlap between exclusions based on different criteria. Values show the percent of the column in the row, e.g. 19.4% of "Rec. Exclusions" are in "Hetero / missing outliers" i) "Hetero / missing outliers": '22027-0.0' (Outliers for heterozygosity or missing rate), ii) "Rec. Exclusions": field '22010-0.0' (Recommended genomic analysis exclusions), iii) "High hetero / missing": '22018-0.0', High heterozygosity rate (after correcting for ancestry) or high missing rate, iv) "Mixed Ancestry": '22018-0.0', Participant self-declared as having a mixed ancestral background, and v) "Discordant Sex": as described in the sample QC methods.

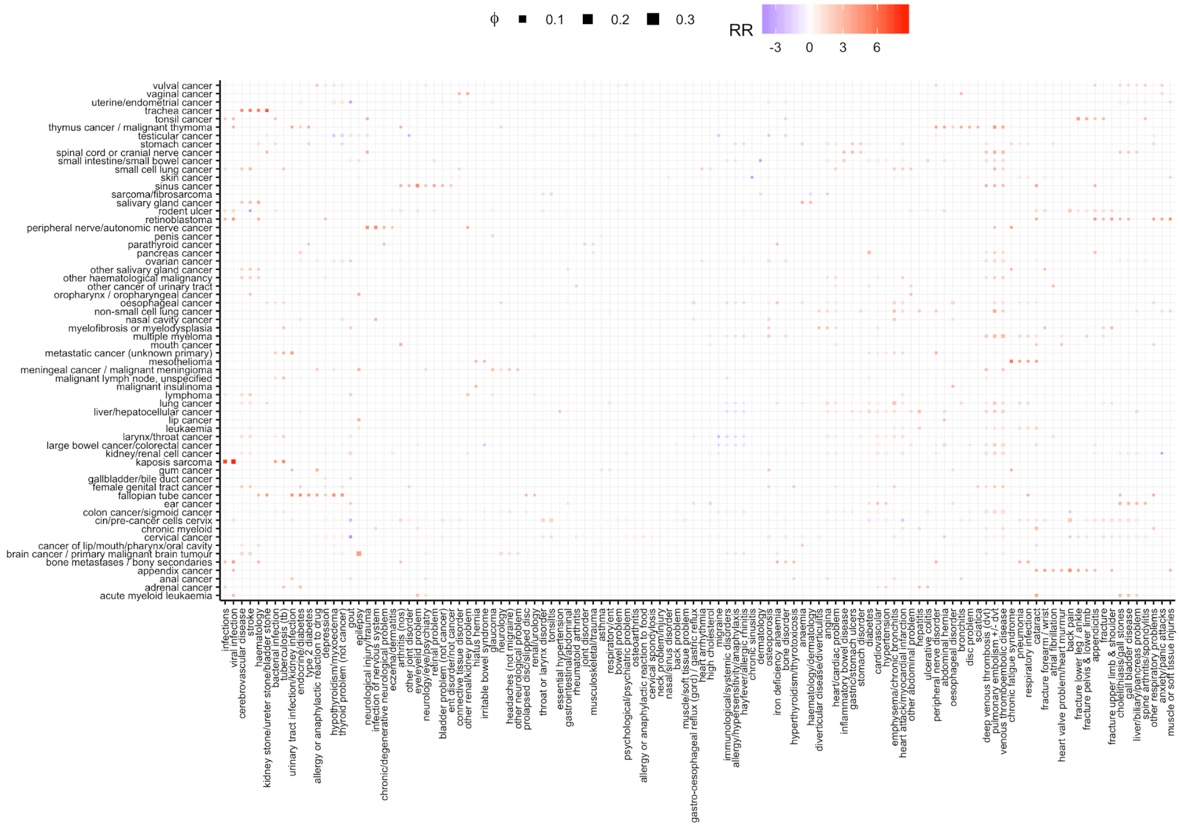


Figure S45: Cancer - disease co-occurrence matrix summarizing relative risk scores and correlations. Each row shows a cancer type and column shows a disease. The color is defined by relative risk scores while the size is determined by ϕ value (in the same scale as Figure S9 for better comparison), indicating the robustness. Associations for the 114 diseases and 62 cancers that have at least one relative risk ratio higher than four ($log_2RR \geq 2$) or lower than minus four ($log_2RR \leq -2$) are plotted.

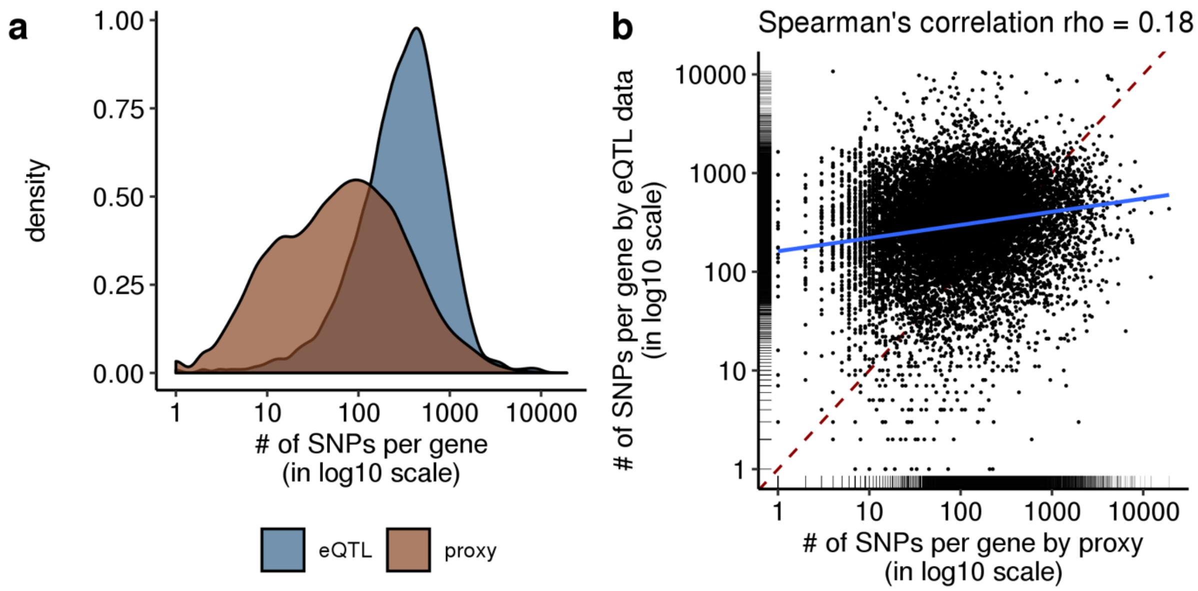


Figure S46: a) Density plots showing the number of SNPs per gene, based on eQTL data (blue) and proximity (brown). b) Scatter plot between the number of SNPs per gene mapped using genomic proximity (x-axis) or eQTL data (y-axis). Each dot represents a gene and the blue line shows the linear model. Dashed red line shows one-to-one relationship. The rug-plots on the axes show the marginal distribution of genes.
